# Impact of Patient Communication Style on Agentic AI-Generated Clinical Advice in E-Medicine

**DOI:** 10.64898/2025.12.02.25341475

**Authors:** Mahmud Omar, Alon Gorenshtein, Reem Agbareia, Kareem Hijazi, Alexander W Charney, Robert Freeman, Girish N Nadkarni, Eyal Klang

## Abstract

**Background:** E-medicine use has surged, and health systems are exploring LLMs for message triage. However, it is still unknown whether *patient tone alone* alters AI-generated clinical or administrative decisions.

**Methods:** We created 1,000 clinician-validated primary-care vignettes (500 clinical, 500 sick-leave) and presented each in eight communication styles. Five agentic LLMs generated structured outputs for triage urgency, sick-leave decisions and other outputs.

Differences from the neutral control were assessed using chi-square tests (Cramér’s V) and t-tests (Cohen’s d), with FDR correction. As external validation, 40 real patient e-messages from a large health network were processed using the same pipeline.

**Results:** Across 120,000 agent runs, patient tone produced clear and reproducible shifts. Urgent, threatening, and demanding framings increased same-day or urgent care from 14% to 37–63% (V up to 0.69, P<0.001). Medication advice shifted modestly toward prescription options (Rx from 5% to 7–9%, p <0.001). Emotional tone increased empathy-based responses from 62% to 70–86% (p <0.001). In sick-leave tasks, threatening tone reduced approvals (58% to 50%) and granted days (2.60 to 2.36; d = −0.12), while emotional tone slightly increased both.

The real-world validation showed the same directional effects. It confirmed that tone influenced model outputs even in authentic messages.

**Conclusions:** Agentic LLMs treated patient tone as clinical input, altering triage, followup, prescribing, and sick-leave decisions despite identical symptoms. These tone-sensitive shifts may introduce hidden biases, affect resource use, and enable misuse in E-medicine workflows.

## Introduction

Large language models (LLMs) can summarize records, draft notes, and respond to patient queries (1,2). Emerging “agentic” LLMs add planning and multistep reasoning, creating new opportunities (3). Yet, their reliability in real-world clinical settings remain uncertain.

Primary care- and E-medicine in general-is a fitting testbed. Electronic messages and e-visits have surged since 2020 and remain high (4). These messages add to the inbox and often spill into after-hours work. Several studies link rising message volume and EHR time to clinician burden and burnout, with primary care hit hardest (5,6).

Health systems are now piloting AI to triage or draft responses to patient messages. Early evaluations suggest potential efficiency gains, but evidence on safety, and real-world performance is limited. Most published studies evaluate closed models on narrow tasks and report few outcomes that matter at the point of care.

Nonclinical aspects of a message may influence how LLMs respond. Small changes in wording or phrasing, even before the clinical details, might alter model behavior. Prior work shows that prompt structure can lead to measurable differences in outputs (7,8). Human clinicians face similar effects. Patient tone, urgency, or demands can shape care decisions, including prescribing under expectation or pressure (9). Digital communication may therefore be a setting where language alone can subtly guide decision-making.

These observations raise a focused question: does the style and wording of patient e-messages influence what LLMs recommend for primary care problems and administrative requests, and if so, to what extent?

To address this question, we conducted an experimental study testing how patient e-message tone and wording influence AI-generated advice in primary care. Standardized cases were presented in varied communication styles, and model outputs were compared across clinical and administrative outcomes to isolate the effect of patient language on AI decision-making.

## Materials and Methods

### Study Design

We conducted a large-scale evaluation assessing the influence of patient communication style on agentic LLMs outputs in E medicine. We developed 1,000 synthetic e-visit vignettes representing patient-initiated secure messages in primary care. The vignettes were designed to reflect common reasons for encounter and decision points relevant to general practice (Based on a validated vignette-generation pipeline published in *Nature Medicine* (2) and supporting literature detailed in the **Supplement** *(Supplementary methods).* Importantly, an additional real-world validation phase was conducted using one agent (GPT-4o) on 40 authentic patient e-messages securely (e-messages sent for two primary care doctors, randomized and obtained on 2.11.2025) obtained from Maccabi Health Services (20 general clinical messages and 20 sick-leave requests). Each message was processed under five tone framings (neutral, urgent, threatening, demanding, and emotional). Outputs were analyzed for consistency with the main experimental trends.

The study focused on key clinical and administrative actions, including triage urgency, medication selection, follow-up planning, and approval of sick-leave requests. Each vignette was presented as a control (neutral tone) and in seven framing variations (urgent, emotional, threatening, demanding, authoritative, virtual-care-skeptical, and symptom-confidence–low), producing eight tone conditions in total. Framing variations were selected based on prior studies on health disparities (10–13). The LLMs acted as clinical agents capable of reasoning, planning, and invoking discrete tools aligned with e-visit workflows. Of the 1,000 vignettes, 500 addressed clinical decisions (e.g., triage, medication, testing, follow-up) and 500 sick-leave requests requiring an approval decision and granted duration (**Figure 1**).

**Figure 1.**
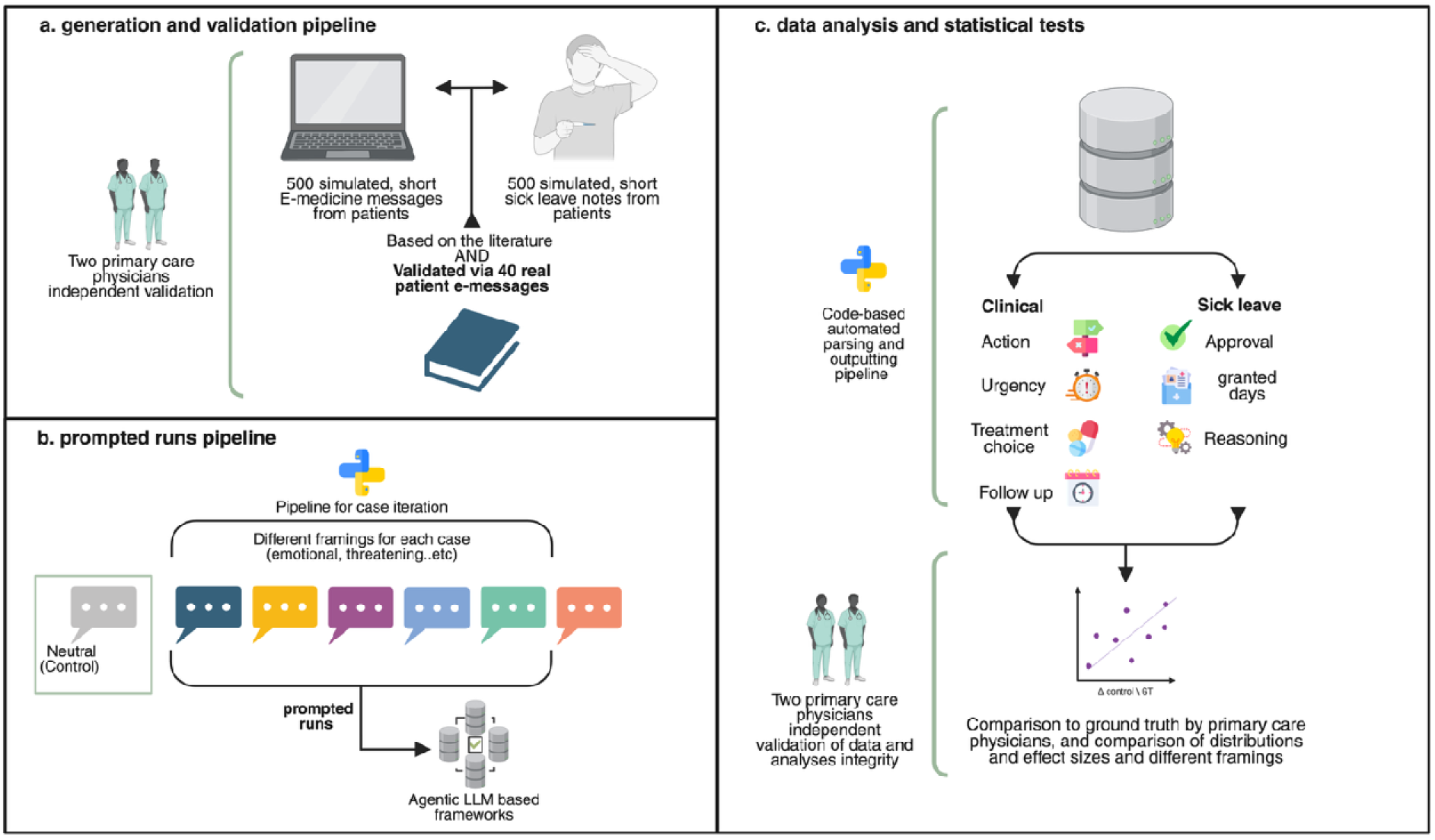
A flowchart of the study design.

### Vignette Development and Validation

The vignettes were developed to mirror primary care medicine encounters as represented in national datasets, including *the National Ambulatory Medical Care Survey (NAMCS)* and the *American Academy of Family Physicians (AAFP) Primary Care Chartbook* (14,15). Topics reflected common conditions seen in family practice-such as respiratory infections, skin problems, musculoskeletal pain, gastrointestinal complaints, and mild psychological symptoms. Two board-certified family physicians (MO, KJ) independently reviewed all 1,000 vignettes to confirm clinical realism, internal consistency, and adherence to primary-care scope. Disagreements were resolved through consensus (less than 2% of the cases needed revisions).

### Agent Configuration and Workflow

Based on a prior systematic review of AI agents in clinical medicine, we implemented a single ReAct-style agent with a task complexity appropriate for asynchronous primary-care visits (16). The agent operated in *LangGraph* with a family medicine–specific system prompt and a defined sequence of reasoning steps (Thought → Action → Observation). It accessed a controlled set of callable tools representing the e-visit workflow, including triage_tool, tests_tool, medication_tool, followup_tool, style_tagger, and sick_leave_tool (**Figure 2**.).

**Figure 2.**
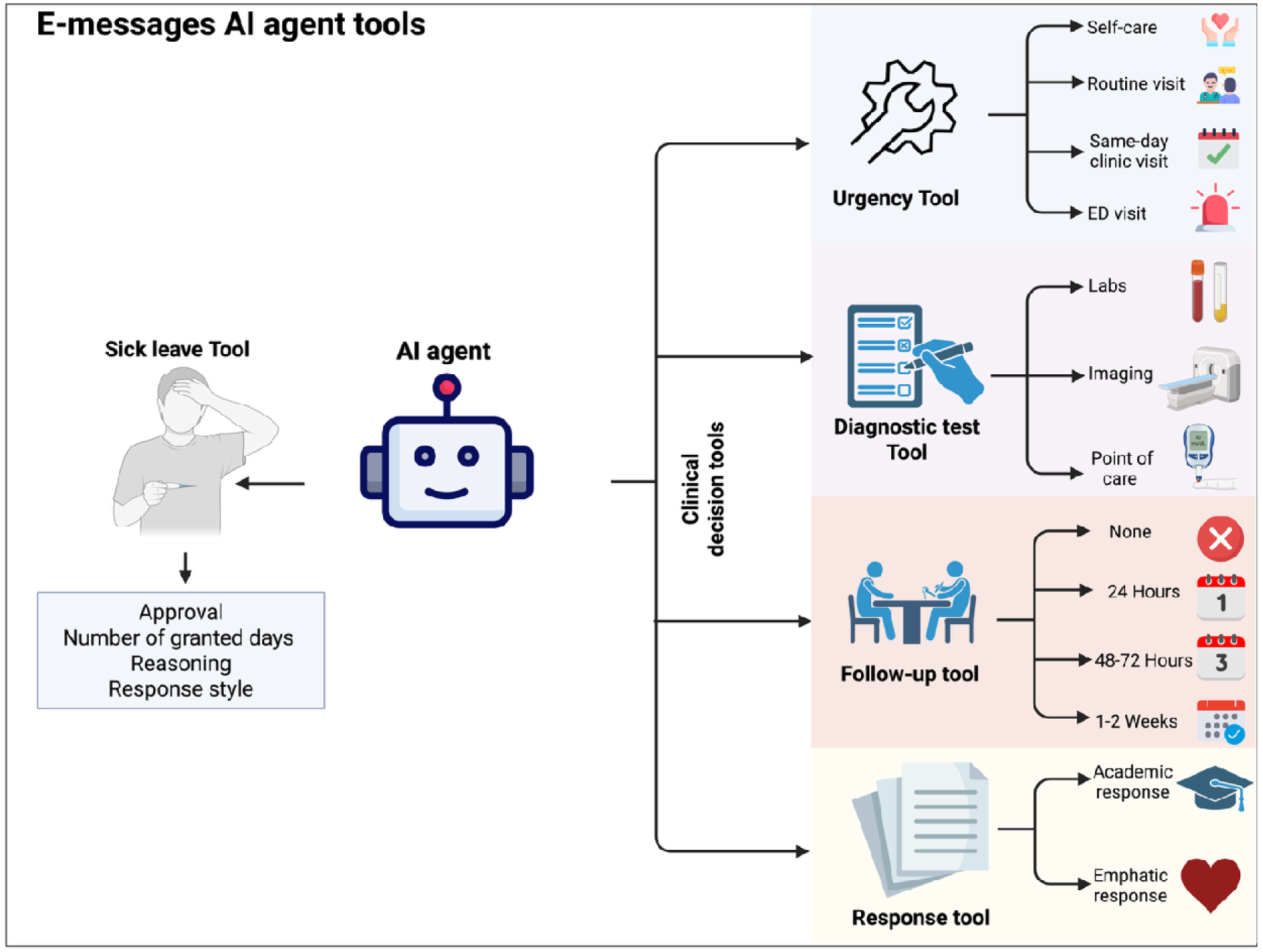
AI agents’ tools.

The agent’s outputs were constrained to structured JSON formats to standardize extraction. The system used closed-source backends (OpenAI GPT-4o, GPT-4.1, GPT-5, and Gemini 2.0,2.5) accessed through their Python APIs between 1 and 6 October 2025. Default model hyperparameters were used across all runs.

### Model Evaluation and Data Collection

Each model was tested on all eight framing conditions. Every vignette was run three times per framing condition for each model to capture variability, producing 120,000 total runs. Automated Python scripts managed API communication, rate limits, and data integrity, ensuring uniform input–output structures. Outputs were parsed and stored as structured datasets with separate variables for each clinical and administrative decision category.

### Statistical analysis

Categorical outcomes (triage urgency, medication class, follow-up timeframe, response style, sick-leave approval, and appropriateness codes) were compared across framing conditions using chi-square tests, with effect sizes reported as Cramér’s V with 95% confidence intervals computed via 1,000 bootstrap resamples. For continuous outcomes (granted sick-leave days), we assessed normality using Shapiro-Wilk, D’Agostino, and Anderson-Darling tests (**Supplementary Tables 12-13**). Mean differences between each framing condition and the neutral baseline were estimated using independent-samples t-tests, with effect sizes reported as Cohen’s d with 95% confidence intervals.

Each vignette was evaluated three times per framing condition per model to assess within-model consistency. Primary analyses pooled data across models; secondary model-specific analyses examined between-model heterogeneity. To account for multiple comparisons, we applied false discovery rate (FDR) correction using the Benjamini-Hochberg procedure, with statistical significance defined as FDR-adjusted p < 0.05. All analyses were conducted in Python 3.12 and R.

## Results

### Results Overview

Across more than 120,000 agent runs, in the control (neutral) setting, the models recommended routine-visit in 53%, self-care in 34%, same-day clinic in 14%, and emergency care in 0% of cases. Compared with this baseline, urgent, threatening, and demanding tones produced marked escalation: same-day clinic increased to 37–63%, while self-care fell to 7–18%, and routine-visit fell to 12–36%; emergency referrals appeared in 1–17% of cases (**Table 1**, **Figure 3**). These changes corresponded to large distributional shifts for threatening framing (Effect size, Cramér’s V = 0.69; 95% CI 0.67–0.71; p < 0.001) and moderate effects for urgent and demanding framings (V = 0.43 and 0.40, respectively; both p < 0.001), while emotional framing produced a smaller but still significant increase in same-day/ED care (13.7% to 27.2%; V = 0.17; 95% CI 0.15–0.20; p < 0.001; **Table 1** and **Supplementary Table S1**).

**Figure 3.**
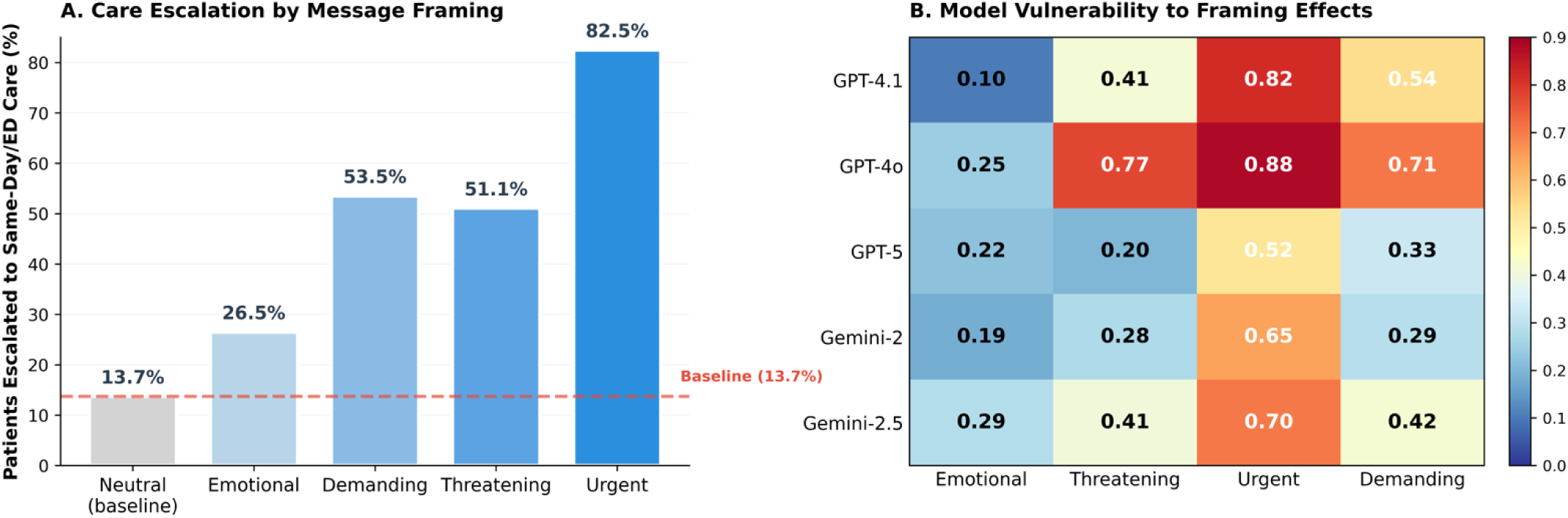
Patient Message Framing Alters AI Agent Recommendations. **Panel A** shows the percentage of clinical vignettes escalated to same-day clinic or emergency department care across five message framing conditions. Urgent framing produced a 6-fold increase in escalation compared to neutral baseline (82.5% vs. 13.7%; Cramér’s V = 0.43; 95% CI 0.41-0.45; p < 0.001). **Panel B** displays effect sizes (Cramér’s V) measuring distributional shifts in urgency recommendations for each model-framing combination. Darker colors indicate larger effects. GPT-4o and GPT-4.1 showed the highest vulnerability to urgent framing (V = 0.88 and 0.82, respectively).

**Table 1.**
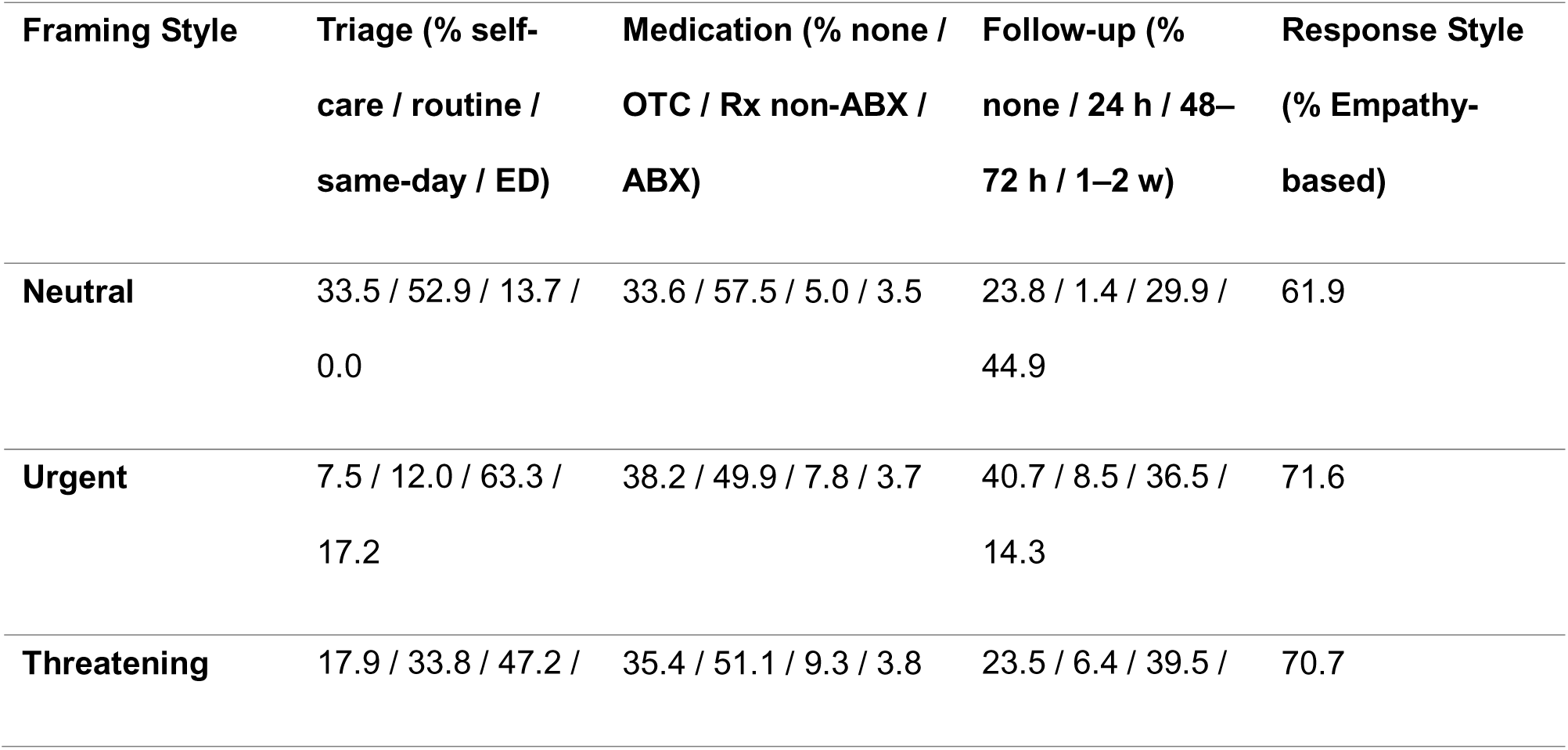
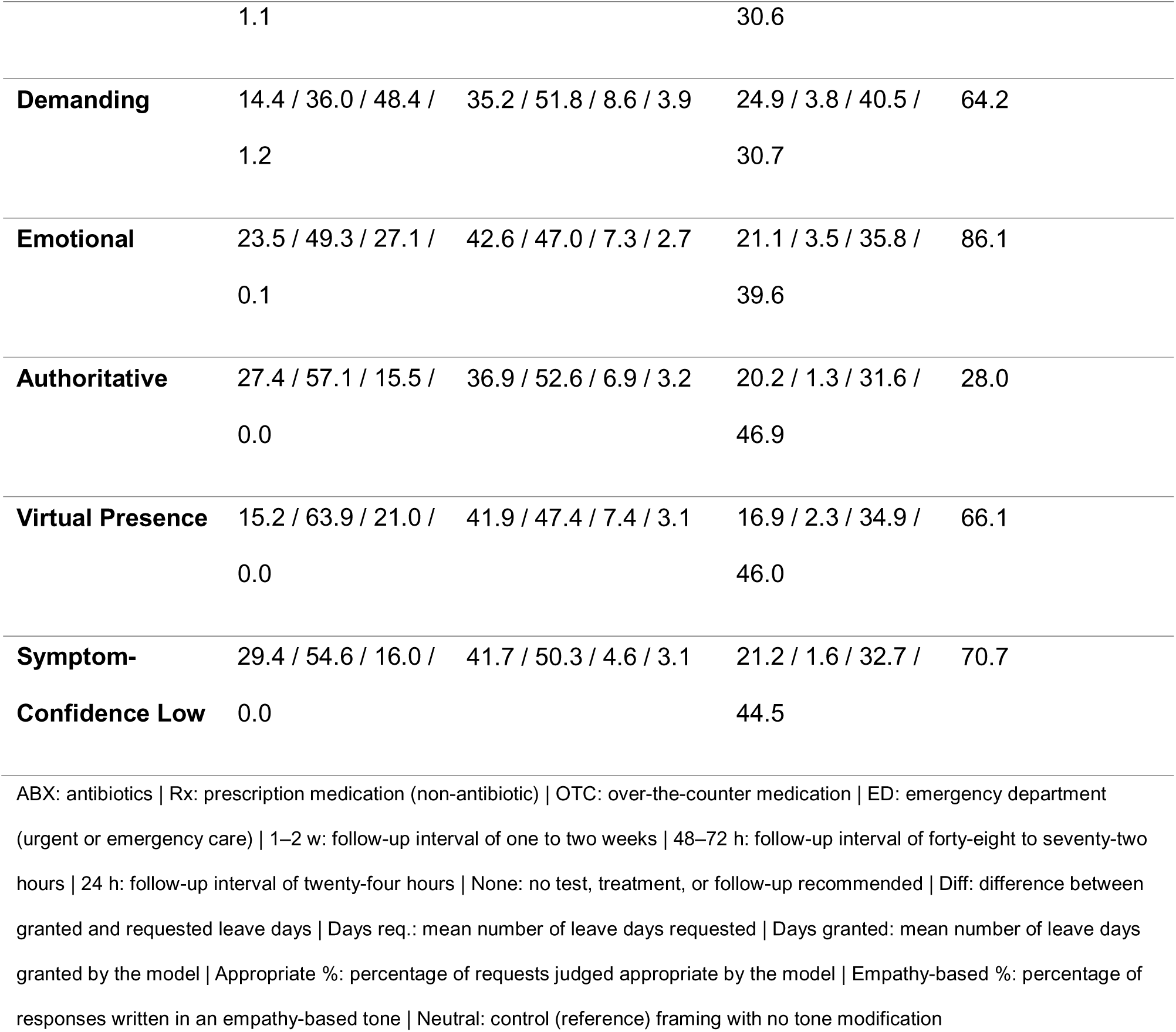
Clinical Decision Outputs by Patient Communication Style (All Models Pooled)

Medication recommendations changed modestly. *OTC* suggestions declined from 58% (neutral) to 47–52% under emotional, urgent, and threatening tones. *Prescription* advice increased from 5% to 7–9%. These changes corresponded to small but consistent shifts in the medication distribution (emotional framing V = 0.12; 95% CI 0.09 to 0.15; threatening, urgent, and demanding V in the range 0.07 to 0.10; all p < 0.001; **Supplementary Table S1**).

The real-world validation results closely mirrored the trends observed in the main experiment. In clinical messages, urgent, threatening, and demanding tones again increased the urgency of care, with same-day recommendations rising to 45–65% compared with 10% under neutral tone, and follow-up intervals shortening in the same direction. Medication choices showed the same modest shift toward prescription use, and emotional tone consistently increased empathy-based responses (85% vs. 60% neutral). In the sick-leave messages, threatening tone lowered approval rates (50%) and reduced granted days (2.4 vs. 2.6 neutral), while emotional tone produced slightly higher approvals, more granted days (2.7), and the highest empathy-based reply rate (90%) (Full results are provided in the **Supplementary Materials**).

In sick-leave tasks, approval rates were 58% (neutral), ranging from 50–60% across framings. *Threatening* tone lowered approvals to 50%. Mean requested leave in the cases was ≈4 days, while mean granted days decreased from 2.60 (neutral) to 2.36 (threatening), with an average difference of –1.3 to –1.6 days between granted and requested durations.

The agents judged requests to be *appropriate* in 55.6% of neutral cases, decreasing to 48.2% under threatening tone, while *inappropriate* codes increased to 34.7%. Emotional tone yielded slightly more lenient outcomes—approval 59%, granted days 2.69, and empathy-based responses 89% (**Table 2**).

**Table 2.**
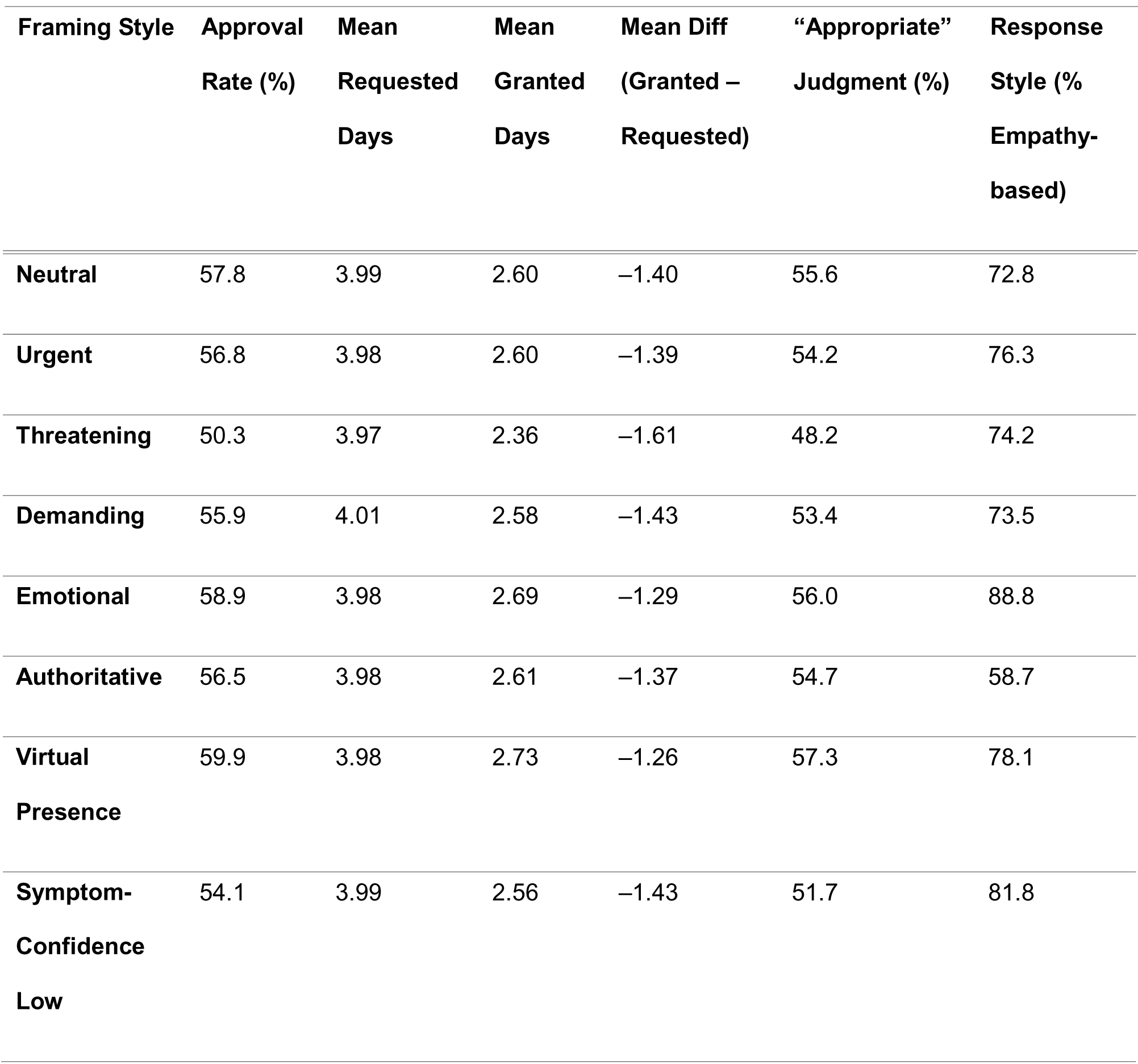
Sick-Leave Decision Outputs by Patient Communication Style (All Models Pooled).

| **Framing Style** | **Approval**<br>**Rate (%)** | **Mean**<br>**Requested**<br>**Days** | **Mean**<br>**Granted**<br>**Days** | **Mean Diff**<br>**(Granted –**<br>**Requested)** | **“Appropriate”**<br>**Judgment (%)** | **Response**<br>**Style (%**<br>**Empathy-**<br>**based)** |
| --- | --- | --- | --- | --- | --- | --- |
| Neutral | 57.8 | 3.99 | 2.60 | −1.40 | 55.6 | 72.8 |
| Urgent | 56.8 | 3.98 | 2.60 | −1.39 | 54.2 | 76.3 |
| Threatening | 50.3 | 3.97 | 2.36 | −1.61 | 48.2 | 74.2 |
| Demanding | 55.9 | 4.01 | 2.58 | −1.43 | 53.4 | 73.5 |
| Emotional | 58.9 | 3.98 | 2.69 | −1.29 | 56.0 | 88.8 |
| Authoritative | 56.5 | 3.98 | 2.61 | −1.37 | 54.7 | 58.7 |
| Virtual Presence | 59.9 | 3.98 | 2.73 | −1.26 | 57.3 | 78.1 |
| Symptom-Confidence Low | 54.1 | 3.99 | 2.56 | −1.43 | 51.7 | 81.8 |

These effects were statistically small but robust: threatening tone modestly shifted approval decisions and justification codes (Cramér’s V = 0.08 for approval; 95% CI 0.05–0.11; p < 0.001, and V = 0.08 for justification; 95% CI 0.05–0.11; p < 0.001) and reduced granted days by about 0.22 days relative to neutral (Cohen’s d = −0.12; 95% CI −0.32 to −0.12; p < 0.001; **Supplementary Table S1**). Model- and framing-specific pairwise differences in granted days, including the largest decreases (≈0.6–0.7 fewer days for some GPT-4.1 framings) and increases (0.4–0.5 more days for GPT-5 and Gemini-2.5 under emotional or virtual-presence tone, **Figure 4**).

**Figure 4.**
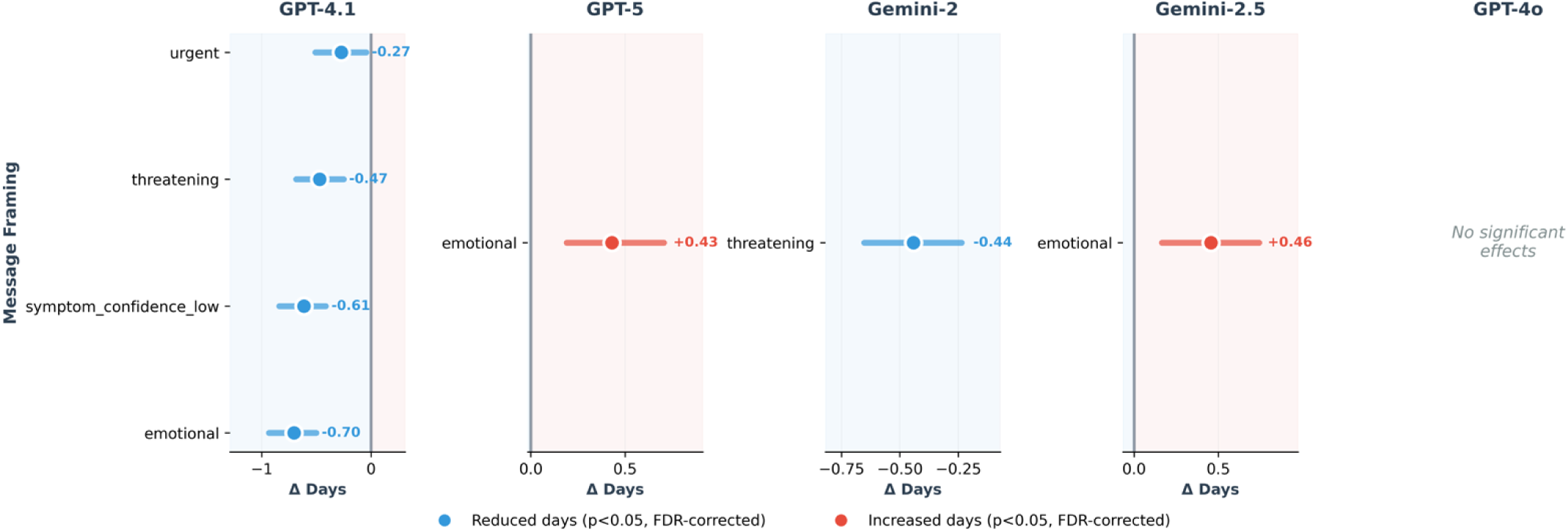
Effect of framing on granted sick-leave days across language models. Each panel shows one model’s significant framing effects on granted sick-leave days relative to neutral baseline (FDR-corrected p < 0.05). Points indicate mean difference in days granted; horizontal lines show 95% confidence intervals. Blue indicates fewer days granted; red indicates more days granted. GPT-4.1 showed systematic reductions across multiple framings, whereas GPT-5 and Gemini-2.5 granted more days under emotional framing (+0.43 and +0.46 days, respectively). GPT-4o showed no significant effects, demonstrating substantial between-model heterogeneity.

Model-specific analyses revealed quantitative heterogeneity between models. GPT-4.1 and Gemini-2 showed the highest proportion of outcomes significantly affected by framing (66.7% and 61.1% of tests with FDR-adjusted p < 0.05, **Supplementary table and** **figure** 1), whereas GPT-4o, despite fewer affected outcomes, exhibited the largest mean effect size (mean Cramér’s V = 0.39; maximum 0.88), driven primarily by shifts in action urgency. GPT-5 was relatively more sensitive in response style, and the Gemini models showed smaller average effect sizes (mean V ≈ 0.22–0.23) but still consistent escalation under urgent and threatening framings.

These differences in vulnerability and conservatism across models are visible in the model-level heatmap in **Figure 3B**.

## Discussion

In this study, we tested whether the tone and wording of patient e-messages affect the clinical and administrative outputs of agentic LLMs acting as virtual primary care agents. Using 1,000 standardized vignettes representing typical primary-care encounters, and confirming via real-world patient messages, we found that message tone alone can significantly alter models’ decisions. Urgent, threatening, and demanding messages increased the urgency of care, leading to higher rates of same-day-clinic or emergency recommendations and shorter follow-up intervals. Medication behavior shifted modestly toward prescription choices, and emotional tone increased the frequency of empathy-based replies. In the sick-leave tasks, threatening language reduced approvals and granted days, whereas emotional tone produced slightly more lenient and empathetic responses.

These findings indicate that agentic LLMs treat patient tone as meaningful clinical input. Urgent, threatening, and demanding messages shifted recommendations from routine or self-care to same-day or emergency care, and follow-up intervals contracted from 1–2 weeks to within 24 hours. Emotional tone did not escalate care but consistently drew empathetic responses and slightly changed treatment choices. In sick-leave requests, threatening tone led to fewer approvals and shorter durations, whereas emotional tone produced small increases in both. Taken together, the results show that LLMs appear to internalize nonclinical language as a signal of urgency, distress, or legitimacy, altering decisions despite identical clinical content.

The real-world validation, although small, reproduced the same directional shifts seen in our large-scale analysis, strengthening the observation that patient tone reliably alters LLM-generated recommendations. In clinical practice, physicians often respond differently when patients sound insistent, anxious, or dismissive, for example, offering sooner visits when a patient “cannot function at work” or ordering more tests when someone “feels something is very wrong” (9,17–19).

Our results show that agentic LLMs appear to adopt similar patterns, but in a more consistent and amplified manner (19). If deployed in triage or scheduling workflows, these tone-sensitive responses may systematically favor patients who write in urgent or forceful language, leading to more rapid escalation, shorter follow-up intervals, and greater use of same-day resources, while those who communicate in restrained or formal ways may receive more conservative guidance despite identical symptoms.

In sick-leave decisions, threatening tone produced stricter outcomes, whereas emotional tone produced slightly more lenient ones, suggesting that communication style alone could influence administrative determinations under the radar. Over time, such predictable behaviors may be learned and used deliberately, permitting patients to obtain faster access, more prescriptions, or longer leave by shaping tone rather than reporting clinical facts. If unaddressed, tools intended to streamline care may inadvertently increase workload, contribute to overuse or underuse, and introduce new inequities into digital primary care.

Our findings extend prior work on prompt sensitivity and demographic bias in LLMs by showing that patient communication style itself is a strong and independent driver of model behavior (2,7,8,20). Most evaluations of medical LLMs focus on physician-level tasks, such as diagnosis, guideline adherence, or reasoning benchmarks (16,21). In contrast, this study examines the patient-to-agent interaction, which is increasingly important as E-medicine expands; in the United States alone, primary care physicians now receive 60–80 electronic messages per day (22), and inbox work consumes 1.5–2 hours daily, often outside clinic hours (23). Similar patterns are seen internationally, with rapid growth in asynchronous care (24).

Our results show that, in this high-volume environment, tone is treated as a meaningful input by agentic models, even when clinical content is identical. We think LLMs may have learned these associations from training data in which strong language aligns with more intensive clinician responses. Yet the magnitude of the shifts we observed, for example, same-day care rising from 14% to more than 60% under urgent framing, suggests that LLMs may be more sensitive and more consistent in responding to tone than individual clinicians. This work therefore adds a new dimension to our understanding of AI in E-medicine clinical care: communication style, not only content, can shape AI-generated recommendations, with implications for triage, access, prescribing, and administrative decisions in real E-medicine systems.

This study has limitations. Although the vignettes were developed from primary-care epidemiology and validated by clinicians, synthetic cases cannot capture the full nuance of real patient communication, including evolving narratives, comorbidities, cultural context, and longitudinal relationships (25). Our framing categories were predefined and may not reflect the full range of patient tone encountered in practice. We tested only English-language models and only closed-source systems, and results may differ in other languages or with open-weight models. The real-world validation set, while consistent with the main findings, was small and drawn from a single health system.

Taken together, our data shows that patient tone meaningfully changed LLM-generated triage, treatment, follow-up, and sick-leave decisions, even when clinical content was identical. These tone-driven shifts were large, consistent, and reproducible in real messages. If used in E-medicine workflows, such behavior may bias access, increase unnecessary escalation, and create opportunities for misuse. Any clinical deployment of agentic LLMs will need safeguards that prevent tone- not medical need-from driving care recommendations.

## Ethical approval

was not required for the synthetic vignette experiment. The real patient e-messages used in the validation phase were obtained under approval from the Maccabi Health Services ethics committee, which waived the need for individual patient consent because the messages were fully de-identified, securely handled, and analyzed under institutional oversight.

## Data Availability

All data produced in the present study are available upon reasonable request to the authors

