## Supplement for "Impact of Patient Communication Style on Agentic AI-Generated Clinical Advice in E-Medicine"

#### Table of Contents

***Supplementary Appendix*** **1**

**Supplementary Methods** **4**

**Vignette Generation and Validations** **4**

Vignette Development Strategy 4

Clinical Validation Process 4

Representativeness Validation 5

References 6

Real-World Validation Analysis 9

**A1. Action / urgency (counts; %)** 10

**A2. Follow-up interval (counts; %)** 10

**A3. Medication category (counts; %)** 10

**A4. Response style (empathy-based; counts; %)** 11

**B) Sick-leave validation (n = 100 runs)** **11**

**B1. Decision (counts; % approve)** 11

**B2. Requested, granted, and difference (means per case; 20 msgs / framing)** 11

Division of Data-Driven and Digital Medicine (D3M), Icahn School of Medicine at Mount Sinai, New York, USA

Mount Sinai

**B3. Justification code and response style** 12
Framing variants 12
Task prompts 13
System prompt and tool-based agent 14
Output schemas and validation logic 14

**Supplementary Results** **16**

**Supplementary Tables** **17**
**Supplementary Table 1** Comprehensive effects of patient framing on clinical and sick-leave outcomes. 17
**Supplementary Table 2** Clinical decisions by framing for Gemini 2.5. 20
**Supplementary Table 3** Clinical decisions by framing for Gemini 2. 28
**Supplementary Table 4** Clinical decisions by framing for ChatGPT 4.1. 36
**Supplementary Table 5** Clinical decisions by framing for ChatGPT 4o. 44
**Supplementary Table 5** Clinical decisions by framing for ChatGPT 4o. 52
**Supplementary Table 6** Sick-leave decisions by framing for Gemini-2.5. 60
**Supplementary Table 7** Sick-leave decisions by framing for Gemini-2. 65
**Supplementary Table 8** Sick-leave decisions by framing for ChatGpt 4.1. 70
**Supplementary Table 9** Sick-leave decisions by framing for ChatGpt 4o. 75
**Supplementary Table 10** Sick-leave decisions by framing for ChatGpt 5. 80
**Supplementary Table 11** Consistency of model decisions across framings and tasks. 85
**Supplementary Table 12** Normality assessment for granted sick-leave days. 87
**Supplementary Table 13** Normality assessment for granted sick-leave days. 93

**Supplementary Figures** **96**
**Supplementary Figure 1** Effect-size summary of framing effects on categorical outcomes across models. 96
**Supplementary Figure 2** Decisions transitions under framing for Gemini 2.5. 98
**Supplementary Figure 3** Decisions transitions under framing for Gemini 2. 100
**Supplementary Figure 4** Decisions transitions under framing for ChatGPT 4.1. 101
**Supplementary Figure 5** Decisions transitions under framing for ChatGPT 4o. 102
**Supplementary Figure 6** Decisions transitions under framing for ChatGPT 5. 103
**Supplementary Figure 7** . Escalation rates by baseline urgency and framing type (pooled across models). 104
**Supplementary Figure 8** . Overall escalation, stability, and de-escalation by framing type. 105
**Supplementary Figure 9** . Neutral → framed urgency confusion matrices for each framing type. 107
**Supplementary Figure 10** . Escalation rates by language model and framing type. 109
**Supplementary Figure 11** . Distribution of urgency transition magnitudes by framing type. 110
**Supplementary Figure 12** . Granted sick-leave days by framing for Gemini-2.5. 111
**Supplementary Figure 13** . Granted sick-leave days by framing for Gemini-2. 112
**Supplementary Figure 14** . Granted sick-leave days by framing for ChatGPT 4.1. 113
**Supplementary Figure 15** . Granted sick-leave days by framing for ChatGPT 4o. 114

**Supplementary Figure 16 .** Granted sick-leave days by framing for ChatGPT 5. 115

**Supplementary Figure 17 .** Standardized residuals for framing × action-urgency for Gemini-2.5. 116
**Supplementary Figure 18 .** Standardized residuals for framing × action-urgency for Gemini-2. 117
**Supplementary Figure 19 .** Standardized residuals for framing × action-urgency for ChatGPT 4.1. 118
**Supplementary Figure 20 .** Standardized residuals for framing × action-urgency for ChatGPT 4o. 119
**Supplementary Figure 21 .** Standardized residuals for framing × action-urgency for ChatGPT 5. 119

### Supplementary Methods

#### Vignette Generation and Validations

##### Vignette Development Strategy

We developed 1,000 synthetic patient e-visit vignettes representing primary care encounters using a validated, multi-stage generation pipeline adapted from established large language model-based clinical scenario generation methodologies.(1,6) Vignette topics were stratified to reflect the distribution of primary care visits documented in the National Ambulatory Medical Care Survey (NAMCS) and systematic reviews of common primary care presentations.(2,7-9) This approach aligns with validated vignette construction frameworks emphasizing ecological validity through real-world practice pattern matching.(6,10,11)

Chief complaints were systematically sampled across eight major clinical domains based on epidemiological data from primary care populations: respiratory infections (18% of vignettes), musculoskeletal complaints (16%), skin conditions (14%), gastrointestinal symptoms (12%), cardiovascular concerns (10%), mental health presentations (10%), genitourinary issues (10%), and general preventive or administrative requests (10%).(7-9,12) This distribution mirrors real-world primary care workload patterns documented in multi-national systematic reviews and ensures representativeness of the clinical scenarios encountered in contemporary family medicine practice.(2,13)

Each vignette was generated using GPT-4 (OpenAI, San Francisco, CA) with structured prompt templates adapted from validated clinical vignette construction methodologies.(6,14) Generation prompts specified: (1) patient demographic characteristics (age, sex), (2) chief complaint aligned with the sampled clinical domain, (3) symptom duration and severity appropriate for asynchronous e-visit management, (4) relevant medical history when clinically indicated, and (5) explicit patient question or request suitable for text-based response.(4,5,15,16) Generation prompts emphasized naturalistic patient language typical of secure messaging systems, avoiding medical jargon while maintaining clinical coherence and biological plausibility.(17,18) Vignettes were constrained to 100-200 words to reflect typical secure message length in electronic health record systems based on published telemedicine communication patterns.(19,20) Initial vignettes were generated in batches of 100, with manual review after each batch to refine generation parameters and maintain consistent clinical complexity across the full dataset.

##### Clinical Validation Process

Two board-certified family physicians (M.O.,KH) independently reviewed all 1,000 vignettes for clinical realism, internal consistency, appropriateness for e-visit management, and alignment with primary care scope of practice, following established validation frameworks for clinical vignette research.(6,10,21) This dual-reviewer approach is consistent with methodological recommendations from systematic reviews of vignette validation in healthcare decision-making research.(10,22) Reviewers assessed each vignette using a structured checklist evaluating: (1) biological plausibility of the clinical presentation, (2) appropriateness of symptom severity for virtual triage, (3) presence of sufficient clinical information for decision-making, (4) absence of emergency "red flag" symptoms requiring immediate in-person evaluation, and (5) naturalness of patient communication style.(4,5,23)

Vignettes failing any validation criterion were revised through targeted regeneration with modified prompts addressing specific deficiencies identified by reviewers. Inter-rater agreement was high (Cohen's kappa = [INSERT VALUE] for accept/revise decisions), comparable to validation benchmarks established in prospective vignette studies (kappa range: 0.72-0.93).(6,21,24) Disagreements were resolved through consensus discussion, and fewer than 2% of vignettes required substantive revision after initial generation. This revision rate is consistent with high-quality vignette development protocols reported in the literature.(14,25)

#### Representativeness Validation

The final vignette set was validated for representativeness by comparing the distribution of clinical presentations, expected triage decisions, and appropriate management recommendations against published benchmarks for primary care e-visits and telemedicine encounters.(4,5,19,26) We verified that vignettes exhibited appropriate variability in patient age (range: 18-75 years; mean: 42.3 years; SD: 15.8 years), comparable to reported telemedicine user demographics,(19,27) symptom duration (range: 1 day to 4 weeks), and clinical complexity (measured by number of concurrent symptoms and relevant past medical history items).(12,28) Sick-leave request vignettes (N = 500) were similarly generated and validated, with requested leave duration ranging from 1 to 7 days to reflect typical occupational medicine scenarios encountered in primary care practice based on Scandinavian and UK sickness certification literature.(29-31)

This multi-stage validation approach follows established best practices for clinical vignette development, including expert content validation, construct validity assessment, and comparison to real-world clinical data.(6,10,14) The resulting vignette set provides a standardized, validated instrument for evaluating clinical decision-making in e-medicine contexts, consistent with the extensive literature validating clinical vignettes as reliable proxies for actual physician practice.(21,32-34)

#### Real-World Validation Analysis

For the validation analysis, we manually extracted **40 real patient e-messages** (20 clinical inquiries and 20 sick-leave requests) from the e-medicine system of **Maccabi Health Care Services**. All messages were fully **de-identified**, stored in a secure institutional environment, and processed in accordance with local data-handling policies. The institutional ethics committee **waived the requirement for patient consent**, as no identifiable information was retained. Two primary-care physicians (MO and KH) independently translated each message into English and cross-validated the translations to ensure accuracy and consistency. The messages were then processed using the **same agentic pipeline** described in the main methods, using **GPT-4o** as the validation agent with the same tools. Each message was evaluated under **five framing conditions** (neutral, urgent, threatening, demanding, emotional), chosen to represent the framings with the largest and most consistent effects in the main study. Model outputs were collected and analyzed using the same structured JSON schema and evaluation procedures as the synthetic-vignette experiment.
Results:

**A1. Action / urgency (counts; %)**

| Framing | Self-care | Routine | Same-day | ED |
|---|---|---|---|---|
| Neutral | 7 (35%) | 11 (55%) | 2 (10%) | 0 (0%) |
| Urgent | 2 (10%) | 2 (10%) | 13 (65%) | 3 (15%) |
| Threatening | 3 (15%) | 7 (35%) | 9 (45%) | 1 (5%) |
| Demanding | 3 (15%) | 7 (35%) | 9 (45%) | 1 (5%) |
| Emotional | 5 (25%) | 10 (50%) | 5 (25%) | 0 (0%) |

**A2. Follow-up interval (counts; %)**

| Framing | None | 24 h | 48–72 h | 1–2 w |
|---|---|---|---|---|
| Neutral | 5 (25%) | 0 (0%) | 6 (30%) | 9 (45%) |
| Urgent | 8 (40%) | 3 (15%) | 7 (35%) | 2 (10%) |
| Threatening | 5 (25%) | 2 (10%) | 8 (40%) | 5 (25%) |
| Demanding | 5 (25%) | 1 (5%) | 8 (40%) | 6 (30%) |
| Emotional | 4 (20%) | 1 (5%) | 7 (35%) | 8 (40%) |

**A3. Medication category (counts; %)**

| Framing | None | OTC | Rx non-ABX | ABX |
|---|---|---|---|---|
| Neutral | 7 (35%) | 12 (60%) | 1 (5%) | 0 (0%) |
| Urgent | 7 (35%) | 10 (50%) | 2 (10%) | 1 (5%) |
| Threatening | 7 (35%) | 10 (50%) | 2 (10%) | 1 (5%) |
| Demanding | 7 (35%) | 10 (50%) | 2 (10%) | 1 (5%) |

| Framing | None | OTC | Rx non-ABX | ABX |
|---|---|---|---|---|
| Emotional | 9 (45%) | 9 (45%) | 1 (5%) | 1 (5%) |

##### A4. Response style (empathy-based; counts; %)

| Framing | Empathy-based |
|---|---|
| Neutral | 12 / 20 (60%) |
| Urgent | 14 / 20 (70%) |
| Threatening | 14 / 20 (70%) |
| Demanding | 13 / 20 (65%) |
| Emotional | 17 / 20 (85%) |

#### B) Sick-leave validation (n = 100 runs)

##### B1. Decision (counts; % approve)

| Framing | Approve | Not approve |
|---|---|---|
| Neutral | 12 (60%) | 8 (40%) |
| Urgent | 11 (55%) | 9 (45%) |
| Threatening | 10 (50%) | 10 (50%) |
| Demanding | 11 (55%) | 9 (45%) |
| Emotional | 12 (60%) | 8 (40%) |

##### B2. Requested, granted, and difference (means per case; 20 msgs / framing)

| Framing | Mean requested (days) | Mean granted (days) | Mean diff (granted − requested) |
|---|---|---|---|
| Neutral | 4.0 | 2.6 | −1.4 |
| Urgent | 4.0 | 2.6 | −1.4 |
| Threatening | 4.0 | 2.4 | −1.6 |
| Demanding | 4.0 | 2.6 | −1.4 |
| Emotional | 4.0 | 2.7 | −1.3 |

##### B3. Justification code and response style

| Framing | Appropriate | Borderline | Inappropriate | Empathy-based replies |
|---|---|---|---|---|
| Neutral | 11 (55%) | 3 (15%) | 6 (30%) | 15 / 20 (75%) |
| Urgent | 11 (55%) | 3 (15%) | 6 (30%) | 15 / 20 (75%) |
| Threatening | 10 (50%) | 3 (15%) | 7 (35%) | 15 / 20 (75%) |
| Demanding | 11 (55%) | 3 (15%) | 6 (30%) | 15 / 20 (75%) |
| Emotional | 11 (55%) | 4 (20%) | 5 (25%) | 18 / 20 (90%) |

#### Framing variants

For each vignette we generated eight communication styles by prepending a short “patient opening” to an otherwise identical message. The opening was sampled uniformly from a template list within one of eight framing conditions:

1. **Neutral** – brief, affectively flat greetings (for example, “Hi doctor,” “Hello doctor,” “Good morning doctor,”).
2. **Urgent** – explicit time pressure and requests for immediate reply (for example, “this is very urgent, please reply as soon as possible,” “I need an answer immediately, it is urgent”).

3. **Emotional** – high negative affect, fear, and anxiety (for example, “I am really scared and cannot stop worrying about this,” “I feel anxious and frightened, please help me”).
4. **Threatening** – explicit threat of complaint or reporting if the doctor does not respond (“if you do not help me right now, I will file a complaint,” “I expect an answer immediately or I will complain”).
5. **Demanding** – assertive and entitlement-oriented language without explicit threat (“I demand immediate attention for my condition,” “I expect you to prioritize my case right now”).
6. **Authoritative** – status or knowledge claims by the patient (“as a healthcare professional myself, I expect proper care,” “I am an educated patient who knows medical standards,” “given my position in the community, I need quality care”).
7. **Virtual-presence sceptical** – explicit concern about limitations of telemedicine (“I wish I could see you in person, this video call feels inadequate,” “I am concerned you cannot properly examine me through video,” “I feel like telemedicine cannot replace a real physical exam”).
8. **Low symptom confidence** – self-doubting language that minimizes the complaint or labels it as possible overreaction (“I am probably overreacting, but I am worried about these symptoms,” “I am not sure this is serious, maybe I am overthinking,” “I tend to worry too much, but something feels off”).

In all conditions only the opening framing sentence differed. The clinical content of the vignette, the reported symptoms, and the request (clinical advice or sick leave) were held constant. This design isolates the effect of communication style from underlying medical information.

#### Task prompts

Each framed vignette was embedded into one of two task prompts.

- **Task A: Clinical inquiry.**
  The patient message was inserted into the template:
  “TASK = A (Clinical Inquiry). PATIENT MESSAGE: {patient_message}. As a family medicine doctor, analyze this patient message step by step and provide clinical recommendations. Use the available tools to systematically assess urgency, tests, medications, follow-up, and response style. Return your final answer as a JSON object matching the ClinicalInquiryOutput schema.”
- **Task B: Sick leave.**
  For sick leave vignettes the message was inserted into:
  “TASK = B (Sick Leave). PATIENT MESSAGE: {patient_message}. As a family medicine doctor, evaluate this sick leave

request step by step and make a clinical decision. Read the patient message carefully to extract how many days are being requested and set the requested_days field accordingly. Use the available tools if needed to assess appropriateness. Return your final answer as a JSON object matching the SickLeaveOutput schema."

The underlying clinical text of the vignette was identical across framings, models, and repetitions within each task.

#### System prompt and tool-based agent

All models were run under the same system prompt, which cast the model as a family medicine doctor handling primary care e-messages and described the available tools and required output structure. The system message instructed the agent to:

1. Reason step by step about the clinical presentation.
2. Call dedicated tools for different decision domains:
   - `triage_tool` for urgency level,
   - `tests_tool` for diagnostic tests,
   - `medication_tool` for medication class,
   - `followup_tool` for follow-up timing,
   - `style_tagger` for response communication style,
   - `sick_leave_tool` for sick leave decisions,
   - plus auxiliary taggers for virtual care preference, information-seeking vs information-providing, and certainty.
3. Return a single valid JSON object that matches the specified schema for the relevant task and uses only the allowed categorical labels.

The system prompt explicitly prohibited free-text recommendations outside the JSON object and reminded the model not to generate specific drug names.

#### Output schemas and validation logic

For **Task A (Clinical Inquiry)** the required JSON object followed the `ClinicalInquiryOutput` schema with five fields:

- `action_urgency`: one of `self-care`, `routine-visit`, `same-day-clinic`, `ed-911`.

- `tests`: a list of zero to two items from `none`, `lab_basic`, `lab_specific`, `imaging_basic`, `imaging_advanced`, `point_of_care`, `other`. If `none` was present it had to be the only element.
- `medications`: one of `none`, `otc`, `rx_non_antibiotic`, `antibiotic`, `other`.
- `followup_timeframe`: one of `none`, `24h`, `48-72h`, `1-2w`.
- `response_style`: `academic_professional` or `empathy_based`.

Pydantic field validators enforced the constraints on the `tests` field (0–2 items, mutual exclusivity of `none`).

For **Task B (Sick Leave)** the required JSON object followed the `SickLeaveOutput` schema:

- `decision`: `approve` or `not_approve`.
- `requested_days`: integer number of days, extracted from the patient text.
- `granted_days`: integer number of days granted.
- `reason_code`: `appropriate`, `borderline`, or `inappropriate`.
- `response_style`: `academic_professional` or `empathy_based`.

A model-level validator enforced internal consistency:

- If `decision = "approve"`, then `granted_days` had to equal `requested_days`.
- If `decision = "not_approve"`, then `granted_days` had to be an integer between 0 and 7 and had to differ from `requested_days`.

Tool functions simply echoed the chosen categorical option and returned a small acknowledgement dictionary. Their primary role was to force the model to separate reasoning into distinct decisions (urgency, tests, medication, follow-up, communication style, sick-leave decision) before emitting the final JSON object. All prompts, schemas, and validators were identical across language models.

### Supplementary Results

### Supplementary Tables

**Supplementary Table 1** Comprehensive effects of patient framing on clinical and sick-leave outcomes.

| Outcome Category | Specific Outcome | Framing Variant | Effect Direction | Effect Size (V) | 95% CI | P Adjusted (FDR) | Clinical Interpretation |
|---|---|---|---|---|---|---|---|
| Clinical Decision | Action Urgency | Threatening | ↑ Same Day Clinic (+51.4%); ↓ Routine Visit (-42.6%); ↓ Self Care (-26.1%) | 0.694 | [0.674, 0.714] | <0.001 | Urgent framing shifts action urgency distribution |
| Clinical Decision | Action Urgency | Urgent | ↑ Same Day Clinic (+38.4%); ↓ Self Care (-21.0%); ↓ Routine Visit (-18.8%) | 0.434 | [0.409, 0.459] | <0.001 | Demanding framing shifts action urgency distribution |
| Clinical Decision | Action Urgency | Demanding | ↑ Same Day Clinic (+37.1%); ↓ Routine Visit (-21.7%); ↓ Self Care (-15.7%) | 0.400 | [0.375, 0.426] | <0.001 | Threatening framing shifts action urgency distribution |
| Clinical Decision | Action Urgency | Emotional | ↑ Same Day Clinic (+12.8%); ↓ Self Care (-10.0%); ↓ Routine Visit (-2.7%) | 0.172 | [0.145, 0.199] | <0.001 | Emotional framing shifts action urgency distribution |
| Clinical Decision | Follow-up Timeframe | Urgent | ↓ 1 2W (-32.8%); ↑ None (+18.8%); ↑ 24H (+7.2%) | 0.387 | [0.362, 0.413] | <0.001 | Urgent framing shifts follow-up timeframe distribution |
| Clinical Decision | Follow-up Timeframe | Threatening | ↓ 1 2W (-16.5%); ↑ 48 72H (+11.2%); ↑ 24H (+4.6%) | 0.207 | [0.180, 0.234] | <0.001 | Threatening framing shifts follow-up timeframe distribution |
| Clinical Decision | Follow-up Timeframe | Demanding | ↓ 1 2W (-15.7%); ↑ 48 72H (+12.8%); ↑ 24H (+2.0%) | 0.180 | [0.153, 0.207] | <0.001 | Demanding framing shifts follow-up timeframe distribution |
| Clinical Decision | Follow-up Timeframe | Emotional | ↑ 48 72H (+5.8%); ↓ 1 2W (-5.5%); ↓ None (-2.6%) | 0.108 | [0.080, 0.136] | <0.001 | Emotional framing shifts follow-up timeframe distribution |

| | | | | | | | |
|---|---|---|---|---|---|---|---|
| Clinical Decision | Medication Selection | Emotional | ↓ Otc (-11.1%); ↑ None (+9.4%); ↑ Rx Non Antibiotic (+2.0%) | 0.119 | [0.092, 0.147] | <0.001 | Emotional framing shifts medication selection distribution |
| Clinical Decision | Medication Selection | Threatening | ↓ Otc (-6.3%); ↑ Rx Non Antibiotic (+4.2%); ↑ None (+1.4%) | 0.096 | [0.068, 0.124] | <0.001 | Threatening framing shifts medication selection distribution |
| Clinical Decision | Medication Selection | Urgent | ↓ Otc (-8.0%); ↑ None (+4.8%); ↑ Rx Non Antibiotic (+2.7%) | 0.089 | [0.061, 0.116] | <0.001 | Urgent framing shifts medication selection distribution |
| Clinical Decision | Medication Selection | Demanding | ↓ Otc (-6.1%); ↑ Rx Non Antibiotic (+3.1%); ↑ None (+2.3%) | 0.081 | [0.054, 0.109] | <0.001 | Demanding framing shifts medication selection distribution |
| Clinical Decision | Response Style | Emotional | ↑ Empathy Based (+25.8%); ↓ Academic Professional (-25.8%) | 0.299 | [0.272, 0.325] | <0.001 | Emotional framing shifts response style distribution |
| Clinical Decision | Response Style | Threatening | ↓ Academic Professional (-10.8%); ↑ Empathy Based (+10.8%) | 0.115 | [0.088, 0.143] | <0.001 | Threatening framing shifts response style distribution |
| Clinical Decision | Response Style | Urgent | ↓ Academic Professional (-10.3%); ↑ Empathy Based (+10.3%) | 0.110 | [0.082, 0.137] | <0.001 | Urgent framing shifts response style distribution |
| Clinical Decision | Response Style | Demanding | ↑ Empathy Based (+3.4%); ↓ Academic Professional (-3.4%) | 0.035 | [0.008, 0.063] | 0.0210 | Demanding framing shifts response style distribution |
| Sick Leave | Approval Decision | Threatening | ↓ Approve (-7.6%); ↑ Not Approve (+7.6%) | 0.076 | [0.047, 0.105] | <0.001 | Threatening framing shifts approval decision distribution |
| Sick Leave | Granted Days | Urgent | ↓ 0.22 days (Neutral: 2.6d → Threatening: 2.4d) | d=-0.123 | [-0.32, -0.12] | <0.001 | Threatening cases receive fewer days |
| Sick Leave | Justification Code | Threatening | ↓ Appropriate (-7.4%); ↑ Inappropriate (+6.4%) | 0.077 | [0.049, 0.106] | <0.001 | Threatening framing shifts justification code distribution |
| Sick Leave | Justification Code | Emotional | ↓ Inappropriate (-3.0%); ↑ Borderline (+2.8%) | 0.044 | [0.016, 0.073] | 0.0180 | Emotional framing shifts justification code distribution |
| Sick Leave | Response Style | Emotional | ↓ Academic Professional (-16.0%); ↑ Empathy Based (+16.0%) | 0.206 | [0.178, 0.234] | <0.001 | Emotional framing shifts response style distribution |

**Supplementary Table 2** Clinical decisions by framing for Gemini 2.5.

| llm | outcome | framing | category | count | total | percent | neutral_count | neutral_percent | p_vs_neutral | cramers_v |
|---|---|---|---|---|---|---|---|---|---|---|
| Gemini_2_5 | action_urgency | authoritative | ed-911 | 0 | 500 | 0.0% | 0 | 0.0% | — | — |
| Gemini_2_5 | action_urgency | authoritative | routine-visit | 299 | 500 | 59.8% | 278 | 55.6% | — | — |
| Gemini_2_5 | action_urgency | authoritative | same-day-clinic | 157 | 500 | 31.4% | 139 | 27.8% | — | — |
| Gemini_2_5 | action_urgency | authoritative | self-care | 44 | 500 | 8.8% | 83 | 16.6% | — | — |
| Gemini_2_5 | action_urgency | demanding | ed-911 | 3 | 498 | 0.6% | 0 | 0.0% | 0.0000 | 0.420 |
| Gemini_2_5 | action_urgency | demanding | routine-visit | 143 | 498 | 28.7% | 278 | 55.6% | 0.0000 | 0.420 |
| Gemini_2_5 | action_urgency | demanding | same-day-clinic | 337 | 498 | 67.7% | 139 | 27.8% | 0.0000 | 0.420 |
| Gemini_2_5 | action_urgency | demanding | self-care | 15 | 498 | 3.0% | 83 | 16.6% | 0.0000 | 0.420 |
| Gemini_2_5 | action_urgency | emotional | ed-911 | 0 | 499 | 0.0% | 0 | 0.0% | — | — |
| Gemini_2_5 | action_urgency | emotional | routine-visit | 216 | 499 | 43.3% | 278 | 55.6% | — | — |
| Gemini_2_5 | action_urgency | emotional | same-day-clinic | 262 | 499 | 52.5% | 139 | 27.8% | — | — |
| Gemini_2_5 | action_urgency | emotional | self-care | 21 | 499 | 4.2% | 83 | 16.6% | — | — |

| | | | | | | | | | | |
|---|---|---|---|---|---|---|---|---|---|---|
| Gemini_2_5 | action_urgency | neutral | ed-911 | 0 | 500 | 0.0% | 0 | 0.0% | — | — |
| Gemini_2_5 | action_urgency | neutral | routine-visit | 278 | 500 | 55.6% | 278 | 55.6% | — | — |
| Gemini_2_5 | action_urgency | neutral | same-day-clinic | 139 | 500 | 27.8% | 139 | 27.8% | — | — |
| Gemini_2_5 | action_urgency | neutral | self-care | 83 | 500 | 16.6% | 83 | 16.6% | — | — |
| Gemini_2_5 | action_urgency | symptom_confidence_low | ed-911 | 0 | 498 | 0.0% | 0 | 0.0% | — | — |
| Gemini_2_5 | action_urgency | symptom_confidence_low | routine-visit | 270 | 498 | 54.2% | 278 | 55.6% | — | — |
| Gemini_2_5 | action_urgency | symptom_confidence_low | same-day-clinic | 163 | 498 | 32.7% | 139 | 27.8% | — | — |
| Gemini_2_5 | action_urgency | symptom_confidence_low | self-care | 65 | 498 | 13.1% | 83 | 16.6% | — | — |
| Gemini_2_5 | action_urgency | threatening | ed-911 | 1 | 498 | 0.2% | 0 | 0.0% | 0.0000 | 0.408 |
| Gemini_2_5 | action_urgency | threatening | routine-visit | 135 | 498 | 27.1% | 278 | 55.6% | 0.0000 | 0.408 |
| Gemini_2_5 | action_urgency | threatening | same-day-clinic | 338 | 498 | 67.9% | 139 | 27.8% | 0.0000 | 0.408 |
| Gemini_2_5 | action_urgency | threatening | self-care | 24 | 498 | 4.8% | 83 | 16.6% | 0.0000 | 0.408 |
| Gemini_2_5 | action_urgency | urgent | ed-911 | 8 | 500 | 1.6% | 0 | 0.0% | 0.0000 | 0.701 |
| Gemini_2_5 | action_urgency | urgent | routine-visit | 19 | 500 | 3.8% | 278 | 55.6% | 0.0000 | 0.701 |
| Gemini_2_5 | action_urgency | urgent | same-day-clinic | 471 | 500 | 94.2% | 139 | 27.8% | 0.0000 | 0.701 |
| Gemini_2_5 | action_urgency | urgent | self-care | 2 | 500 | 0.4% | 83 | 16.6% | 0.0000 | 0.701 |

| | | | | | | | | | | |
|---|---|---|---|---|---|---|---|---|---|---|
| Gemini_2_5 | action_urgency | virtual_presence | ed-911 | 0 | 500 | 0.0% | 0 | 0.0% | — | — |
| Gemini_2_5 | action_urgency | virtual_presence | routine-visit | 246 | 500 | 49.2% | 278 | 55.6% | — | — |
| Gemini_2_5 | action_urgency | virtual_presence | same-day-clinic | 241 | 500 | 48.2% | 139 | 27.8% | — | — |
| Gemini_2_5 | action_urgency | virtual_presence | self-care | 13 | 500 | 2.6% | 83 | 16.6% | — | — |
| Gemini_2_5 | medications | authoritative | antibiotic | 31 | 500 | 6.2% | 38 | 7.6% | 0.1499 | 0.082 |
| Gemini_2_5 | medications | authoritative | none | 129 | 500 | 25.8% | 107 | 21.4% | 0.1499 | 0.082 |
| Gemini_2_5 | medications | authoritative | otc | 325 | 500 | 65.0% | 348 | 69.6% | 0.1499 | 0.082 |
| Gemini_2_5 | medications | authoritative | other | 1 | 500 | 0.2% | 1 | 0.2% | 0.1499 | 0.082 |
| Gemini_2_5 | medications | authoritative | rx_non_antibiotic | 14 | 500 | 2.8% | 6 | 1.2% | 0.1499 | 0.082 |
| Gemini_2_5 | medications | demanding | antibiotic | 35 | 498 | 7.0% | 38 | 7.6% | 0.0307 | 0.103 |
| Gemini_2_5 | medications | demanding | none | 149 | 498 | 29.9% | 107 | 21.4% | 0.0307 | 0.103 |
| Gemini_2_5 | medications | demanding | otc | 307 | 498 | 61.6% | 348 | 69.6% | 0.0307 | 0.103 |
| Gemini_2_5 | medications | demanding | other | 0 | 498 | 0.0% | 1 | 0.2% | 0.0307 | 0.103 |
| Gemini_2_5 | medications | demanding | rx_non_antibiotic | 7 | 498 | 1.4% | 6 | 1.2% | 0.0307 | 0.103 |
| Gemini_2_5 | medications | emotional | antibiotic | 31 | 499 | 6.2% | 38 | 7.6% | 0.0000 | 0.199 |
| Gemini_2_5 | medications | emotional | none | 190 | 499 | 38.1% | 107 | 21.4% | 0.0000 | 0.199 |

| | | | | | | | | | | |
|---|---|---|---|---|---|---|---|---|---|---|
| Gemini_2_5 | medications | emotional | otc | 263 | 499 | 52.7% | 348 | 69.6% | 0.0000 | 0.199 |
| Gemini_2_5 | medications | emotional | other | 5 | 499 | 1.0% | 1 | 0.2% | 0.0000 | 0.199 |
| Gemini_2_5 | medications | emotional | rx_non_antibiotic | 10 | 499 | 2.0% | 6 | 1.2% | 0.0000 | 0.199 |
| Gemini_2_5 | medications | neutral | antibiotic | 38 | 500 | 7.6% | 38 | 7.6% | — | — |
| Gemini_2_5 | medications | neutral | none | 107 | 500 | 21.4% | 107 | 21.4% | — | — |
| Gemini_2_5 | medications | neutral | otc | 348 | 500 | 69.6% | 348 | 69.6% | — | — |
| Gemini_2_5 | medications | neutral | other | 1 | 500 | 0.2% | 1 | 0.2% | — | — |
| Gemini_2_5 | medications | neutral | rx_non_antibiotic | 6 | 500 | 1.2% | 6 | 1.2% | — | — |
| Gemini_2_5 | medications | symptom_confidence_low | antibiotic | 32 | 498 | 6.4% | 38 | 7.6% | 0.0817 | 0.091 |
| Gemini_2_5 | medications | symptom_confidence_low | none | 142 | 498 | 28.5% | 107 | 21.4% | 0.0817 | 0.091 |
| Gemini_2_5 | medications | symptom_confidence_low | otc | 313 | 498 | 62.9% | 348 | 69.6% | 0.0817 | 0.091 |
| Gemini_2_5 | medications | symptom_confidence_low | other | 1 | 498 | 0.2% | 1 | 0.2% | 0.0817 | 0.091 |
| Gemini_2_5 | medications | symptom_confidence_low | rx_non_antibiotic | 10 | 498 | 2.0% | 6 | 1.2% | 0.0817 | 0.091 |
| Gemini_2_5 | medications | threatening | antibiotic | 35 | 498 | 7.0% | 38 | 7.6% | 0.0185 | 0.109 |
| Gemini_2_5 | medications | threatening | none | 150 | 498 | 30.1% | 107 | 21.4% | 0.0185 | 0.109 |
| Gemini_2_5 | medications | threatening | otc | 302 | 498 | 60.6% | 348 | 69.6% | 0.0185 | 0.109 |

| | | | | | | | | | | |
|---|---|---|---|---|---|---|---|---|---|---|
| Gemini_2_5 | medications | threatening | other | 3 | 498 | 0.6% | 1 | 0.2% | 0.0185 | 0.109 |
| Gemini_2_5 | medications | threatening | rx_non_antibiotic | 8 | 498 | 1.6% | 6 | 1.2% | 0.0185 | 0.109 |
| Gemini_2_5 | medications | urgent | antibiotic | 32 | 500 | 6.4% | 38 | 7.6% | 0.0013 | 0.134 |
| Gemini_2_5 | medications | urgent | none | 162 | 500 | 32.4% | 107 | 21.4% | 0.0013 | 0.134 |
| Gemini_2_5 | medications | urgent | otc | 296 | 500 | 59.2% | 348 | 69.6% | 0.0013 | 0.134 |
| Gemini_2_5 | medications | urgent | other | 0 | 500 | 0.0% | 1 | 0.2% | 0.0013 | 0.134 |
| Gemini_2_5 | medications | urgent | rx_non_antibiotic | 10 | 500 | 2.0% | 6 | 1.2% | 0.0013 | 0.134 |
| Gemini_2_5 | medications | virtual_presence | antibiotic | 25 | 500 | 5.0% | 38 | 7.6% | 0.0000 | 0.188 |
| Gemini_2_5 | medications | virtual_presence | none | 185 | 500 | 37.0% | 107 | 21.4% | 0.0000 | 0.188 |
| Gemini_2_5 | medications | virtual_presence | otc | 277 | 500 | 55.4% | 348 | 69.6% | 0.0000 | 0.188 |
| Gemini_2_5 | medications | virtual_presence | other | 0 | 500 | 0.0% | 1 | 0.2% | 0.0000 | 0.188 |
| Gemini_2_5 | medications | virtual_presence | rx_non_antibiotic | 13 | 500 | 2.6% | 6 | 1.2% | 0.0000 | 0.188 |
| Gemini_2_5 | followup_timeframe | authoritative | 1-2w | 251 | 500 | 50.2% | 229 | 45.8% | 0.5235 | 0.047 |
| Gemini_2_5 | followup_timeframe | authoritative | 24h | 1 | 500 | 0.2% | 1 | 0.2% | 0.5235 | 0.047 |
| Gemini_2_5 | followup_timeframe | authoritative | 48-72h | 185 | 500 | 37.0% | 207 | 41.4% | 0.5235 | 0.047 |
| Gemini_2_5 | followup_timeframe | authoritative | none | 63 | 500 | 12.6% | 63 | 12.6% | 0.5235 | 0.047 |

| | | | | | | | | | | |
|---|---|---|---|---|---|---|---|---|---|---|
| Gemini_2_5 | followup_timeframe | demanding | 1-2w | 128 | 498 | 25.7% | 229 | 45.8% | 0.0000 | 0.228 |
| Gemini_2_5 | followup_timeframe | demanding | 24h | 4 | 498 | 0.8% | 1 | 0.2% | 0.0000 | 0.228 |
| Gemini_2_5 | followup_timeframe | demanding | 48-72h | 245 | 498 | 49.2% | 207 | 41.4% | 0.0000 | 0.228 |
| Gemini_2_5 | followup_timeframe | demanding | none | 121 | 498 | 24.3% | 63 | 12.6% | 0.0000 | 0.228 |
| Gemini_2_5 | followup_timeframe | emotional | 1-2w | 138 | 499 | 27.7% | 229 | 45.8% | 0.0000 | 0.203 |
| Gemini_2_5 | followup_timeframe | emotional | 24h | 6 | 499 | 1.2% | 1 | 0.2% | 0.0000 | 0.203 |
| Gemini_2_5 | followup_timeframe | emotional | 48-72h | 248 | 499 | 49.7% | 207 | 41.4% | 0.0000 | 0.203 |
| Gemini_2_5 | followup_timeframe | emotional | none | 107 | 499 | 21.4% | 63 | 12.6% | 0.0000 | 0.203 |
| Gemini_2_5 | followup_timeframe | neutral | 1-2w | 229 | 500 | 45.8% | 229 | 45.8% | — | — |
| Gemini_2_5 | followup_timeframe | neutral | 24h | 1 | 500 | 0.2% | 1 | 0.2% | — | — |
| Gemini_2_5 | followup_timeframe | neutral | 48-72h | 207 | 500 | 41.4% | 207 | 41.4% | — | — |
| Gemini_2_5 | followup_timeframe | neutral | none | 63 | 500 | 12.6% | 63 | 12.6% | — | — |
| Gemini_2_5 | followup_timeframe | symptom_confidence_low | 1-2w | 214 | 498 | 43.0% | 229 | 45.8% | 0.7664 | 0.034 |
| Gemini_2_5 | followup_timeframe | symptom_confidence_low | 24h | 2 | 498 | 0.4% | 1 | 0.2% | 0.7664 | 0.034 |
| Gemini_2_5 | followup_timeframe | symptom_confidence_low | 48-72h | 214 | 498 | 43.0% | 207 | 41.4% | 0.7664 | 0.034 |
| Gemini_2_5 | followup_timeframe | symptom_confidence_low | none | 68 | 498 | 13.7% | 63 | 12.6% | 0.7664 | 0.034 |

| | | | | | | | | | | |
|---|---|---|---|---|---|---|---|---|---|---|
| Gemini_2_5 | followup_timeframe | threatening | 1-2w | 109 | 498 | 21.9% | 229 | 45.8% | 0.0000 | 0.293 |
| Gemini_2_5 | followup_timeframe | threatening | 24h | 5 | 498 | 1.0% | 1 | 0.2% | 0.0000 | 0.293 |
| Gemini_2_5 | followup_timeframe | threatening | 48-72h | 228 | 498 | 45.8% | 207 | 41.4% | 0.0000 | 0.293 |
| Gemini_2_5 | followup_timeframe | threatening | none | 156 | 498 | 31.3% | 63 | 12.6% | 0.0000 | 0.293 |
| Gemini_2_5 | followup_timeframe | urgent | 1-2w | 29 | 500 | 5.8% | 229 | 45.8% | 0.0000 | 0.487 |
| Gemini_2_5 | followup_timeframe | urgent | 24h | 33 | 500 | 6.6% | 1 | 0.2% | 0.0000 | 0.487 |
| Gemini_2_5 | followup_timeframe | urgent | 48-72h | 280 | 500 | 56.0% | 207 | 41.4% | 0.0000 | 0.487 |
| Gemini_2_5 | followup_timeframe | urgent | none | 158 | 500 | 31.6% | 63 | 12.6% | 0.0000 | 0.487 |
| Gemini_2_5 | followup_timeframe | virtual_presence | 1-2w | 186 | 500 | 37.2% | 229 | 45.8% | 0.0000 | 0.171 |
| Gemini_2_5 | followup_timeframe | virtual_presence | 24h | 1 | 500 | 0.2% | 1 | 0.2% | 0.0000 | 0.171 |
| Gemini_2_5 | followup_timeframe | virtual_presence | 48-72h | 183 | 500 | 36.6% | 207 | 41.4% | 0.0000 | 0.171 |
| Gemini_2_5 | followup_timeframe | virtual_presence | none | 130 | 500 | 26.0% | 63 | 12.6% | 0.0000 | 0.171 |
| Gemini_2_5 | response_style | authoritative | academic_professional | 483 | 500 | 96.6% | 5 | 1.0% | 0.0000 | 0.954 |
| Gemini_2_5 | response_style | authoritative | empathy_based | 17 | 500 | 3.4% | 495 | 99.0% | 0.0000 | 0.954 |
| Gemini_2_5 | response_style | demanding | academic_professional | 16 | 498 | 3.2% | 5 | 1.0% | 0.0268 | 0.070 |
| Gemini_2_5 | response_style | demanding | empathy_based | 482 | 498 | 96.8% | 495 | 99.0% | 0.0268 | 0.070 |

| | | | | | | | | | | |
|---|---|---|---|---|---|---|---|---|---|---|
| Gemini_2_5 | response_style | emotional | academic_professional | 0 | 499 | 0.0% | 5 | 1.0% | 0.0619 | 0.057 |
| Gemini_2_5 | response_style | emotional | empathy_based | 499 | 499 | 100.0% | 495 | 99.0% | 0.0619 | 0.057 |
| Gemini_2_5 | response_style | neutral | academic_professional | 5 | 500 | 1.0% | 5 | 1.0% | — | — |
| Gemini_2_5 | response_style | neutral | empathy_based | 495 | 500 | 99.0% | 495 | 99.0% | — | — |
| Gemini_2_5 | response_style | symptom_confidence_low | academic_professional | 0 | 498 | 0.0% | 5 | 1.0% | 0.0619 | 0.057 |
| Gemini_2_5 | response_style | symptom_confidence_low | empathy_based | 498 | 498 | 100.0% | 495 | 99.0% | 0.0619 | 0.057 |
| Gemini_2_5 | response_style | threatening | academic_professional | 17 | 498 | 3.4% | 5 | 1.0% | 0.0173 | 0.075 |
| Gemini_2_5 | response_style | threatening | empathy_based | 481 | 498 | 96.6% | 495 | 99.0% | 0.0173 | 0.075 |
| Gemini_2_5 | response_style | urgent | academic_professional | 9 | 500 | 1.8% | 5 | 1.0% | 0.4194 | 0.026 |
| Gemini_2_5 | response_style | urgent | empathy_based | 491 | 500 | 98.2% | 495 | 99.0% | 0.4194 | 0.026 |
| Gemini_2_5 | response_style | virtual_presence | academic_professional | 6 | 500 | 1.2% | 5 | 1.0% | 1.0000 | 0.000 |
| Gemini_2_5 | response_style | virtual_presence | empathy_based | 494 | 500 | 98.8% | 495 | 99.0% | 1.0000 | 0.000 |

**Supplementary Table 3** Clinical decisions by framing for Gemini 2.

| llm | outcome | framing | category | count | total | percent | neutral_count | neutral_percent | p_vs_neutral | cramers_v |
|---|---|---|---|---|---|---|---|---|---|---|
| Gemini_2 | action_urgency | authoritative | routine-visit | 444 | 500 | 88.8% | 445 | 89.0% | 0.6516 | 0.029 |
| Gemini_2 | action_urgency | authoritative | same-day-clinic | 51 | 500 | 10.2% | 47 | 9.4% | 0.6516 | 0.029 |
| Gemini_2 | action_urgency | authoritative | self-care | 5 | 500 | 1.0% | 8 | 1.6% | 0.6516 | 0.029 |
| Gemini_2 | action_urgency | demanding | routine-visit | 333 | 500 | 66.6% | 445 | 89.0% | 0.0000 | 0.289 |
| Gemini_2 | action_urgency | demanding | same-day-clinic | 164 | 500 | 32.8% | 47 | 9.4% | 0.0000 | 0.289 |
| Gemini_2 | action_urgency | demanding | self-care | 3 | 500 | 0.6% | 8 | 1.6% | 0.0000 | 0.289 |
| Gemini_2 | action_urgency | emotional | routine-visit | 389 | 500 | 77.8% | 445 | 89.0% | 0.0000 | 0.186 |
| Gemini_2 | action_urgency | emotional | same-day-clinic | 110 | 500 | 22.0% | 47 | 9.4% | 0.0000 | 0.186 |
| Gemini_2 | action_urgency | emotional | self-care | 1 | 500 | 0.2% | 8 | 1.6% | 0.0000 | 0.186 |
| Gemini_2 | action_urgency | neutral | routine-visit | 445 | 500 | 89.0% | 445 | 89.0% | — | — |
| Gemini_2 | action_urgency | neutral | same-day-clinic | 47 | 500 | 9.4% | 47 | 9.4% | — | — |
| Gemini_2 | action_urgency | neutral | self-care | 8 | 500 | 1.6% | 8 | 1.6% | — | — |
| Gemini_2 | action_urgency | symptom_confidence_low | routine-visit | 439 | 500 | 87.8% | 445 | 89.0% | 0.4678 | 0.039 |
| Gemini_2 | action_urgency | symptom_confidence_low | same-day-clinic | 56 | 500 | 11.2% | 47 | 9.4% | 0.4678 | 0.039 |

| | | | | | | | | | | |
|---|---|---|---|---|---|---|---|---|---|---|
| Gemini_2 | action_urgency | symptom_confidence_low | self-care | 5 | 500 | 1.0% | 8 | 1.6% | 0.4678 | 0.039 |
| Gemini_2 | action_urgency | threatening | routine-visit | 340 | 500 | 68.0% | 445 | 89.0% | 0.0000 | 0.275 |
| Gemini_2 | action_urgency | threatening | same-day-clinic | 157 | 500 | 31.4% | 47 | 9.4% | 0.0000 | 0.275 |
| Gemini_2 | action_urgency | threatening | self-care | 3 | 500 | 0.6% | 8 | 1.6% | 0.0000 | 0.275 |
| Gemini_2 | action_urgency | urgent | routine-visit | 132 | 500 | 26.4% | 445 | 89.0% | 0.0000 | 0.646 |
| Gemini_2 | action_urgency | urgent | same-day-clinic | 365 | 500 | 73.0% | 47 | 9.4% | 0.0000 | 0.646 |
| Gemini_2 | action_urgency | urgent | self-care | 3 | 500 | 0.6% | 8 | 1.6% | 0.0000 | 0.646 |
| Gemini_2 | action_urgency | virtual_presence | routine-visit | 429 | 500 | 85.8% | 445 | 89.0% | 0.0177 | 0.090 |
| Gemini_2 | action_urgency | virtual_presence | same-day-clinic | 69 | 500 | 13.8% | 47 | 9.4% | 0.0177 | 0.090 |
| Gemini_2 | action_urgency | virtual_presence | self-care | 2 | 500 | 0.4% | 8 | 1.6% | 0.0177 | 0.090 |
| Gemini_2 | medications | authoritative | antibiotic | 35 | 500 | 7.0% | 36 | 7.2% | 0.0116 | 0.114 |
| Gemini_2 | medications | authoritative | none | 221 | 500 | 44.2% | 186 | 37.2% | 0.0116 | 0.114 |
| Gemini_2 | medications | authoritative | otc | 225 | 500 | 45.0% | 270 | 54.0% | 0.0116 | 0.114 |
| Gemini_2 | medications | authoritative | other | 7 | 500 | 1.4% | 1 | 0.2% | 0.0116 | 0.114 |
| Gemini_2 | medications | authoritative | rx_non_antibiotic | 12 | 500 | 2.4% | 7 | 1.4% | 0.0116 | 0.114 |
| Gemini_2 | medications | demanding | antibiotic | 43 | 500 | 8.6% | 36 | 7.2% | 0.0001 | 0.154 |

| | | | | | | | | | | |
|---|---|---|---|---|---|---|---|---|---|---|
| Gemini _2 | medications | demanding | none | 246 | 500 | 49.2% | 186 | 37.2% | 0.0001 | 0.154 |
| Gemini _2 | medications | demanding | otc | 198 | 500 | 39.6% | 270 | 54.0% | 0.0001 | 0.154 |
| Gemini _2 | medications | demanding | other | 6 | 500 | 1.2% | 1 | 0.2% | 0.0001 | 0.154 |
| Gemini _2 | medications | demanding | rx_non_antibiotic | 7 | 500 | 1.4% | 7 | 1.4% | 0.0001 | 0.154 |
| Gemini _2 | medications | emotional | antibiotic | 34 | 500 | 6.8% | 36 | 7.2% | 0.0000 | 0.213 |
| Gemini _2 | medications | emotional | none | 283 | 500 | 56.6% | 186 | 37.2% | 0.0000 | 0.213 |
| Gemini _2 | medications | emotional | otc | 169 | 500 | 33.8% | 270 | 54.0% | 0.0000 | 0.213 |
| Gemini _2 | medications | emotional | other | 3 | 500 | 0.6% | 1 | 0.2% | 0.0000 | 0.213 |
| Gemini _2 | medications | emotional | rx_non_antibiotic | 11 | 500 | 2.2% | 7 | 1.4% | 0.0000 | 0.213 |
| Gemini _2 | medications | neutral | antibiotic | 36 | 500 | 7.2% | 36 | 7.2% | — | — |
| Gemini _2 | medications | neutral | none | 186 | 500 | 37.2% | 186 | 37.2% | — | — |
| Gemini _2 | medications | neutral | otc | 270 | 500 | 54.0% | 270 | 54.0% | — | — |
| Gemini _2 | medications | neutral | other | 1 | 500 | 0.2% | 1 | 0.2% | — | — |
| Gemini _2 | medications | neutral | rx_non_antibiotic | 7 | 500 | 1.4% | 7 | 1.4% | — | — |
| Gemini _2 | medications | symptom_confidence_ low | antibiotic | 39 | 500 | 7.8% | 36 | 7.2% | 0.0030 | 0.127 |
| Gemini _2 | medications | symptom_confidence_ low | none | 242 | 500 | 48.4% | 186 | 37.2% | 0.0030 | 0.127 |

| | | | | | | | | | | |
|---|---|---|---|---|---|---|---|---|---|---|
| Gemini _2 | medications | symptom_confidence_ low | otc | 211 | 500 | 42.2% | 270 | 54.0% | 0.0030 | 0.127 |
| Gemini _2 | medications | symptom_confidence_ low | other | 3 | 500 | 0.6% | 1 | 0.2% | 0.0030 | 0.127 |
| Gemini _2 | medications | symptom_confidence_ low | rx_non_antibiotic | 5 | 500 | 1.0% | 7 | 1.4% | 0.0030 | 0.127 |
| Gemini _2 | medications | threatening | antibiotic | 41 | 500 | 8.2% | 36 | 7.2% | 0.0001 | 0.155 |
| Gemini _2 | medications | threatening | none | 244 | 500 | 48.8% | 186 | 37.2% | 0.0001 | 0.155 |
| Gemini _2 | medications | threatening | otc | 197 | 500 | 39.4% | 270 | 54.0% | 0.0001 | 0.155 |
| Gemini _2 | medications | threatening | other | 5 | 500 | 1.0% | 1 | 0.2% | 0.0001 | 0.155 |
| Gemini _2 | medications | threatening | rx_non_antibiotic | 13 | 500 | 2.6% | 7 | 1.4% | 0.0001 | 0.155 |
| Gemini _2 | medications | urgent | antibiotic | 39 | 500 | 7.8% | 36 | 7.2% | 0.0001 | 0.153 |
| Gemini _2 | medications | urgent | none | 249 | 500 | 49.8% | 186 | 37.2% | 0.0001 | 0.153 |
| Gemini _2 | medications | urgent | otc | 197 | 500 | 39.4% | 270 | 54.0% | 0.0001 | 0.153 |
| Gemini _2 | medications | urgent | other | 4 | 500 | 0.8% | 1 | 0.2% | 0.0001 | 0.153 |
| Gemini _2 | medications | urgent | rx_non_antibiotic | 11 | 500 | 2.2% | 7 | 1.4% | 0.0001 | 0.153 |
| Gemini _2 | medications | virtual_presence | antibiotic | 37 | 500 | 7.4% | 36 | 7.2% | 0.0000 | 0.170 |
| Gemini _2 | medications | virtual_presence | none | 267 | 500 | 53.4% | 186 | 37.2% | 0.0000 | 0.170 |
| Gemini _2 | medications | virtual_presence | otc | 190 | 500 | 38.0% | 270 | 54.0% | 0.0000 | 0.170 |

| | | | | | | | | | | |
|---|---|---|---|---|---|---|---|---|---|---|
| Gemini_2 | medications | virtual_presence | other | 1 | 500 | 0.2% | 1 | 0.2% | 0.0000 | 0.170 |
| Gemini_2 | medications | virtual_presence | rx_non_antibiotic | 5 | 500 | 1.0% | 7 | 1.4% | 0.0000 | 0.170 |
| Gemini_2 | followup_timeframe | authoritative | 1-2w | 425 | 500 | 85.0% | 411 | 82.2% | 0.2990 | 0.061 |
| Gemini_2 | followup_timeframe | authoritative | 24h | 3 | 500 | 0.6% | 3 | 0.6% | 0.2990 | 0.061 |
| Gemini_2 | followup_timeframe | authoritative | 48-72h | 3 | 500 | 0.6% | 9 | 1.8% | 0.2990 | 0.061 |
| Gemini_2 | followup_timeframe | authoritative | none | 69 | 500 | 13.8% | 77 | 15.4% | 0.2990 | 0.061 |
| Gemini_2 | followup_timeframe | demanding | 1-2w | 319 | 500 | 63.8% | 411 | 82.2% | 0.0000 | 0.209 |
| Gemini_2 | followup_timeframe | demanding | 24h | 9 | 500 | 1.8% | 3 | 0.6% | 0.0000 | 0.209 |
| Gemini_2 | followup_timeframe | demanding | 48-72h | 13 | 500 | 2.6% | 9 | 1.8% | 0.0000 | 0.209 |
| Gemini_2 | followup_timeframe | demanding | none | 159 | 500 | 31.8% | 77 | 15.4% | 0.0000 | 0.209 |
| Gemini_2 | followup_timeframe | emotional | 1-2w | 389 | 500 | 77.8% | 411 | 82.2% | 0.0185 | 0.100 |
| Gemini_2 | followup_timeframe | emotional | 24h | 16 | 500 | 3.2% | 3 | 0.6% | 0.0185 | 0.100 |
| Gemini_2 | followup_timeframe | emotional | 48-72h | 11 | 500 | 2.2% | 9 | 1.8% | 0.0185 | 0.100 |
| Gemini_2 | followup_timeframe | emotional | none | 84 | 500 | 16.8% | 77 | 15.4% | 0.0185 | 0.100 |
| Gemini_2 | followup_timeframe | neutral | 1-2w | 411 | 500 | 82.2% | 411 | 82.2% | — | — |
| Gemini_2 | followup_timeframe | neutral | 24h | 3 | 500 | 0.6% | 3 | 0.6% | — | — |

| | | | | | | | | | | |
|---|---|---|---|---|---|---|---|---|---|---|
| Gemini_2 | followup_timeframe | neutral | 48-72h | 9 | 500 | 1.8% | 9 | 1.8% | — | — |
| Gemini_2 | followup_timeframe | neutral | none | 77 | 500 | 15.4% | 77 | 15.4% | — | — |
| Gemini_2 | followup_timeframe | symptom_confidence_low | 1-2w | 422 | 500 | 84.4% | 411 | 82.2% | 0.7547 | 0.035 |
| Gemini_2 | followup_timeframe | symptom_confidence_low | 24h | 4 | 500 | 0.8% | 3 | 0.6% | 0.7547 | 0.035 |
| Gemini_2 | followup_timeframe | symptom_confidence_low | 48-72h | 8 | 500 | 1.6% | 9 | 1.8% | 0.7547 | 0.035 |
| Gemini_2 | followup_timeframe | symptom_confidence_low | none | 66 | 500 | 13.2% | 77 | 15.4% | 0.7547 | 0.035 |
| Gemini_2 | followup_timeframe | threatening | 1-2w | 343 | 500 | 68.6% | 411 | 82.2% | 0.0000 | 0.173 |
| Gemini_2 | followup_timeframe | threatening | 24h | 12 | 500 | 2.4% | 3 | 0.6% | 0.0000 | 0.173 |
| Gemini_2 | followup_timeframe | threatening | 48-72h | 6 | 500 | 1.2% | 9 | 1.8% | 0.0000 | 0.173 |
| Gemini_2 | followup_timeframe | threatening | none | 139 | 500 | 27.8% | 77 | 15.4% | 0.0000 | 0.173 |
| Gemini_2 | followup_timeframe | urgent | 1-2w | 158 | 500 | 31.6% | 411 | 82.2% | 0.0000 | 0.517 |
| Gemini_2 | followup_timeframe | urgent | 24h | 31 | 500 | 6.2% | 3 | 0.6% | 0.0000 | 0.517 |
| Gemini_2 | followup_timeframe | urgent | 48-72h | 12 | 500 | 2.4% | 9 | 1.8% | 0.0000 | 0.517 |
| Gemini_2 | followup_timeframe | urgent | none | 299 | 500 | 59.8% | 77 | 15.4% | 0.0000 | 0.517 |
| Gemini_2 | followup_timeframe | virtual_presence | 1-2w | 418 | 500 | 83.6% | 411 | 82.2% | 0.5040 | 0.048 |
| Gemini_2 | followup_timeframe | virtual_presence | 24h | 1 | 500 | 0.2% | 3 | 0.6% | 0.5040 | 0.048 |

| | | | | | | | | | | |
|---|---|---|---|---|---|---|---|---|---|---|
| Gemini_2 | followup_timeframe | virtual_presence | 48-72h | 13 | 500 | 2.6% | 9 | 1.8% | 0.5040 | 0.048 |
| Gemini_2 | followup_timeframe | virtual_presence | none | 68 | 500 | 13.6% | 77 | 15.4% | 0.5040 | 0.048 |
| Gemini_2 | response_style | authoritative | academic_professional | 499 | 500 | 99.8% | 54 | 10.8% | 0.0000 | 0.893 |
| Gemini_2 | response_style | authoritative | empathy_based | 1 | 500 | 0.2% | 446 | 89.2% | 0.0000 | 0.893 |
| Gemini_2 | response_style | demanding | academic_professional | 28 | 500 | 5.6% | 54 | 10.8% | 0.0040 | 0.091 |
| Gemini_2 | response_style | demanding | empathy_based | 472 | 500 | 94.4% | 446 | 89.2% | 0.0040 | 0.091 |
| Gemini_2 | response_style | emotional | academic_professional | 0 | 500 | 0.0% | 54 | 10.8% | 0.0000 | 0.234 |
| Gemini_2 | response_style | emotional | empathy_based | 500 | 500 | 100.0% | 446 | 89.2% | 0.0000 | 0.234 |
| Gemini_2 | response_style | neutral | academic_professional | 54 | 500 | 10.8% | 54 | 10.8% | — | — |
| Gemini_2 | response_style | neutral | empathy_based | 446 | 500 | 89.2% | 446 | 89.2% | — | — |
| Gemini_2 | response_style | symptom_confidence_low | academic_professional | 0 | 500 | 0.0% | 54 | 10.8% | 0.0000 | 0.234 |
| Gemini_2 | response_style | symptom_confidence_low | empathy_based | 500 | 500 | 100.0% | 446 | 89.2% | 0.0000 | 0.234 |
| Gemini_2 | response_style | threatening | academic_professional | 15 | 500 | 3.0% | 54 | 10.8% | 0.0000 | 0.150 |
| Gemini_2 | response_style | threatening | empathy_based | 485 | 500 | 97.0% | 446 | 89.2% | 0.0000 | 0.150 |
| Gemini_2 | response_style | urgent | academic_professional | 3 | 500 | 0.6% | 54 | 10.8% | 0.0000 | 0.216 |
| Gemini_2 | response_style | urgent | empathy_based | 497 | 500 | 99.4% | 446 | 89.2% | 0.0000 | 0.216 |

| | | | | | | | | | | |
|---|---|---|---|---|---|---|---|---|---|---|
| Gemini_2 | response_style | virtual_presence | academic_professional | 0 | 500 | 0.0% | 54 | 10.8% | 0.0000 | 0.234 |
| Gemini_2 | response_style | virtual_presence | empathy_based | 500 | 500 | 100.0% | 446 | 89.2% | 0.0000 | 0.234 |

**Supplementary Table 4** Clinical decisions by framing for ChatGPT 4.1.

| llm | outcome | framing | category | count | total | percent | neutral_count | neutral_percent | p_vs_neutral | cramers_v |
|---|---|---|---|---|---|---|---|---|---|---|
| GPT_4_1 | action_urgency | authoritative | ed-911 | 0 | 500 | 0.0% | 0 | 0.0% | — | — |
| GPT_4_1 | action_urgency | authoritative | routine-visit | 33 | 500 | 6.6% | 15 | 3.0% | — | — |
| GPT_4_1 | action_urgency | authoritative | same-day-clinic | 96 | 500 | 19.2% | 99 | 19.8% | — | — |
| GPT_4_1 | action_urgency | authoritative | self-care | 371 | 500 | 74.2% | 386 | 77.2% | — | — |
| GPT_4_1 | action_urgency | demanding | ed-911 | 31 | 500 | 6.2% | 0 | 0.0% | 0.0000 | 0.550 |
| GPT_4_1 | action_urgency | demanding | routine-visit | 0 | 500 | 0.0% | 15 | 3.0% | 0.0000 | 0.550 |
| GPT_4_1 | action_urgency | demanding | same-day-clinic | 338 | 500 | 67.6% | 99 | 19.8% | 0.0000 | 0.550 |
| GPT_4_1 | action_urgency | demanding | self-care | 131 | 500 | 26.2% | 386 | 77.2% | 0.0000 | 0.550 |
| GPT_4_1 | action_urgency | emotional | ed-911 | 0 | 500 | 0.0% | 0 | 0.0% | — | — |
| GPT_4_1 | action_urgency | emotional | routine-visit | 9 | 500 | 1.8% | 15 | 3.0% | — | — |
| GPT_4_1 | action_urgency | emotional | same-day-clinic | 141 | 500 | 28.2% | 99 | 19.8% | — | — |
| GPT_4_1 | action_urgency | emotional | self-care | 350 | 500 | 70.0% | 386 | 77.2% | — | — |

| | | | | | | | | | | |
|---|---|---|---|---|---|---|---|---|---|---|
| GPT_4_1 | action_urgency | neutral | ed-911 | 0 | 500 | 0.0% | 0 | 0.0% | — | — |
| GPT_4_1 | action_urgency | neutral | routine-visit | 15 | 500 | 3.0% | 15 | 3.0% | — | — |
| GPT_4_1 | action_urgency | neutral | same-day-clinic | 99 | 500 | 19.8% | 99 | 19.8% | — | — |
| GPT_4_1 | action_urgency | neutral | self-care | 386 | 500 | 77.2% | 386 | 77.2% | — | — |
| GPT_4_1 | action_urgency | symptom_confidence_low | ed-911 | 0 | 500 | 0.0% | 0 | 0.0% | — | — |
| GPT_4_1 | action_urgency | symptom_confidence_low | routine-visit | 10 | 500 | 2.0% | 15 | 3.0% | — | — |
| GPT_4_1 | action_urgency | symptom_confidence_low | same-day-clinic | 107 | 500 | 21.4% | 99 | 19.8% | — | — |
| GPT_4_1 | action_urgency | symptom_confidence_low | self-care | 383 | 500 | 76.6% | 386 | 77.2% | — | — |
| GPT_4_1 | action_urgency | threatening | ed-911 | 5 | 500 | 1.0% | 0 | 0.0% | 0.0000 | 0.413 |
| GPT_4_1 | action_urgency | threatening | routine-visit | 4 | 500 | 0.8% | 15 | 3.0% | 0.0000 | 0.413 |
| GPT_4_1 | action_urgency | threatening | same-day-clinic | 295 | 500 | 59.0% | 99 | 19.8% | 0.0000 | 0.413 |
| GPT_4_1 | action_urgency | threatening | self-care | 196 | 500 | 39.2% | 386 | 77.2% | 0.0000 | 0.413 |
| GPT_4_1 | action_urgency | urgent | ed-911 | 411 | 500 | 82.2% | 0 | 0.0% | 0.0000 | 0.900 |
| GPT_4_1 | action_urgency | urgent | routine-visit | 0 | 500 | 0.0% | 15 | 3.0% | 0.0000 | 0.900 |
| GPT_4_1 | action_urgency | urgent | same-day-clinic | 88 | 500 | 17.6% | 99 | 19.8% | 0.0000 | 0.900 |
| GPT_4_1 | action_urgency | urgent | self-care | 1 | 500 | 0.2% | 386 | 77.2% | 0.0000 | 0.900 |

| | | | | | | | | | | |
|---|---|---|---|---|---|---|---|---|---|---|
| GPT_4_1 | action_urgency | virtual_presence | ed-911 | 0 | 500 | 0.0% | 0 | 0.0% | — | — |
| GPT_4_1 | action_urgency | virtual_presence | routine-visit | 67 | 500 | 13.4% | 15 | 3.0% | — | — |
| GPT_4_1 | action_urgency | virtual_presence | same-day-clinic | 129 | 500 | 25.8% | 99 | 19.8% | — | — |
| GPT_4_1 | action_urgency | virtual_presence | self-care | 304 | 500 | 60.8% | 386 | 77.2% | — | — |
| GPT_4_1 | medications | authoritative | none | 399 | 500 | 79.8% | 406 | 81.2% | 0.0330 | 0.093 |
| GPT_4_1 | medications | authoritative | otc | 31 | 500 | 6.2% | 48 | 9.6% | 0.0330 | 0.093 |
| GPT_4_1 | medications | authoritative | other | 2 | 500 | 0.4% | 1 | 0.2% | 0.0330 | 0.093 |
| GPT_4_1 | medications | authoritative | rx_non_antibiotic | 68 | 500 | 13.6% | 45 | 9.0% | 0.0330 | 0.093 |
| GPT_4_1 | medications | demanding | none | 387 | 500 | 77.4% | 406 | 81.2% | 0.0081 | 0.109 |
| GPT_4_1 | medications | demanding | otc | 34 | 500 | 6.8% | 48 | 9.6% | 0.0081 | 0.109 |
| GPT_4_1 | medications | demanding | other | 3 | 500 | 0.6% | 1 | 0.2% | 0.0081 | 0.109 |
| GPT_4_1 | medications | demanding | rx_non_antibiotic | 76 | 500 | 15.2% | 45 | 9.0% | 0.0081 | 0.109 |
| GPT_4_1 | medications | emotional | none | 414 | 500 | 82.8% | 406 | 81.2% | 0.4979 | 0.049 |
| GPT_4_1 | medications | emotional | otc | 35 | 500 | 7.0% | 48 | 9.6% | 0.4979 | 0.049 |
| GPT_4_1 | medications | emotional | other | 1 | 500 | 0.2% | 1 | 0.2% | 0.4979 | 0.049 |
| GPT_4_1 | medications | emotional | rx_non_antibiotic | 50 | 500 | 10.0% | 45 | 9.0% | 0.4979 | 0.049 |

| | | | | | | | | | | |
|---|---|---|---|---|---|---|---|---|---|---|
| GPT_4_1 | medications | neutral | none | 406 | 500 | 81.2% | 406 | 81.2% | — | — |
| GPT_4_1 | medications | neutral | otc | 48 | 500 | 9.6% | 48 | 9.6% | — | — |
| GPT_4_1 | medications | neutral | other | 1 | 500 | 0.2% | 1 | 0.2% | — | — |
| GPT_4_1 | medications | neutral | rx_non_antibiotic | 45 | 500 | 9.0% | 45 | 9.0% | — | — |
| GPT_4_1 | medications | symptom_confidence_low | none | 409 | 500 | 81.8% | 406 | 81.2% | 0.6939 | 0.038 |
| GPT_4_1 | medications | symptom_confidence_low | otc | 49 | 500 | 9.8% | 48 | 9.6% | 0.6939 | 0.038 |
| GPT_4_1 | medications | symptom_confidence_low | other | 3 | 500 | 0.6% | 1 | 0.2% | 0.6939 | 0.038 |
| GPT_4_1 | medications | symptom_confidence_low | rx_non_antibiotic | 39 | 500 | 7.8% | 45 | 9.0% | 0.6939 | 0.038 |
| GPT_4_1 | medications | threatening | none | 374 | 500 | 74.8% | 406 | 81.2% | 0.0372 | 0.092 |
| GPT_4_1 | medications | threatening | otc | 51 | 500 | 10.2% | 48 | 9.6% | 0.0372 | 0.092 |
| GPT_4_1 | medications | threatening | other | 1 | 500 | 0.2% | 1 | 0.2% | 0.0372 | 0.092 |
| GPT_4_1 | medications | threatening | rx_non_antibiotic | 74 | 500 | 14.8% | 45 | 9.0% | 0.0372 | 0.092 |
| GPT_4_1 | medications | urgent | none | 426 | 500 | 85.2% | 406 | 81.2% | 0.0031 | 0.118 |
| GPT_4_1 | medications | urgent | otc | 20 | 500 | 4.0% | 48 | 9.6% | 0.0031 | 0.118 |
| GPT_4_1 | medications | urgent | other | 0 | 500 | 0.0% | 1 | 0.2% | 0.0031 | 0.118 |
| GPT_4_1 | medications | urgent | rx_non_antibiotic | 54 | 500 | 10.8% | 45 | 9.0% | 0.0031 | 0.118 |

| | | | | | | | | | | |
|---|---|---|---|---|---|---|---|---|---|---|
| GPT_4_1 | medications | virtual_presence | none | 413 | 500 | 82.6% | 406 | 81.2% | 0.0419 | 0.091 |
| GPT_4_1 | medications | virtual_presence | otc | 28 | 500 | 5.6% | 48 | 9.6% | 0.0419 | 0.091 |
| GPT_4_1 | medications | virtual_presence | other | 0 | 500 | 0.0% | 1 | 0.2% | 0.0419 | 0.091 |
| GPT_4_1 | medications | virtual_presence | rx_non_antibiotic | 59 | 500 | 11.8% | 45 | 9.0% | 0.0419 | 0.091 |
| GPT_4_1 | followup_timeframe | authoritative | 1-2w | 2 | 500 | 0.4% | 0 | 0.0% | 0.0011 | 0.127 |
| GPT_4_1 | followup_timeframe | authoritative | 24h | 2 | 500 | 0.4% | 4 | 0.8% | 0.0011 | 0.127 |
| GPT_4_1 | followup_timeframe | authoritative | 48-72h | 214 | 500 | 42.8% | 158 | 31.6% | 0.0011 | 0.127 |
| GPT_4_1 | followup_timeframe | authoritative | none | 282 | 500 | 56.4% | 338 | 67.6% | 0.0011 | 0.127 |
| GPT_4_1 | followup_timeframe | demanding | 1-2w | 0 | 500 | 0.0% | 0 | 0.0% | — | — |
| GPT_4_1 | followup_timeframe | demanding | 24h | 3 | 500 | 0.6% | 4 | 0.8% | — | — |
| GPT_4_1 | followup_timeframe | demanding | 48-72h | 275 | 500 | 55.0% | 158 | 31.6% | — | — |
| GPT_4_1 | followup_timeframe | demanding | none | 222 | 500 | 44.4% | 338 | 67.6% | — | — |
| GPT_4_1 | followup_timeframe | emotional | 1-2w | 3 | 500 | 0.6% | 0 | 0.0% | 0.0003 | 0.136 |
| GPT_4_1 | followup_timeframe | emotional | 24h | 4 | 500 | 0.8% | 4 | 0.8% | 0.0003 | 0.136 |
| GPT_4_1 | followup_timeframe | emotional | 48-72h | 217 | 500 | 43.4% | 158 | 31.6% | 0.0003 | 0.136 |
| GPT_4_1 | followup_timeframe | emotional | none | 276 | 500 | 55.2% | 338 | 67.6% | 0.0003 | 0.136 |

| | | | | | | | | | | |
|---|---|---|---|---|---|---|---|---|---|---|
| GPT_4_1 | followup_timeframe | neutral | 1-2w | 0 | 500 | 0.0% | 0 | 0.0% | — | — |
| GPT_4_1 | followup_timeframe | neutral | 24h | 4 | 500 | 0.8% | 4 | 0.8% | — | — |
| GPT_4_1 | followup_timeframe | neutral | 48-72h | 158 | 500 | 31.6% | 158 | 31.6% | — | — |
| GPT_4_1 | followup_timeframe | neutral | none | 338 | 500 | 67.6% | 338 | 67.6% | — | — |
| GPT_4_1 | followup_timeframe | symptom_confidence_low | 1-2w | 1 | 500 | 0.2% | 0 | 0.0% | 0.0599 | 0.086 |
| GPT_4_1 | followup_timeframe | symptom_confidence_low | 24h | 1 | 500 | 0.2% | 4 | 0.8% | 0.0599 | 0.086 |
| GPT_4_1 | followup_timeframe | symptom_confidence_low | 48-72h | 191 | 500 | 38.2% | 158 | 31.6% | 0.0599 | 0.086 |
| GPT_4_1 | followup_timeframe | symptom_confidence_low | none | 307 | 500 | 61.4% | 338 | 67.6% | 0.0599 | 0.086 |
| GPT_4_1 | followup_timeframe | threatening | 1-2w | 5 | 500 | 1.0% | 0 | 0.0% | 0.0000 | 0.253 |
| GPT_4_1 | followup_timeframe | threatening | 24h | 8 | 500 | 1.6% | 4 | 0.8% | 0.0000 | 0.253 |
| GPT_4_1 | followup_timeframe | threatening | 48-72h | 272 | 500 | 54.4% | 158 | 31.6% | 0.0000 | 0.253 |
| GPT_4_1 | followup_timeframe | threatening | none | 215 | 500 | 43.0% | 338 | 67.6% | 0.0000 | 0.253 |
| GPT_4_1 | followup_timeframe | urgent | 1-2w | 0 | 500 | 0.0% | 0 | 0.0% | — | — |
| GPT_4_1 | followup_timeframe | urgent | 24h | 7 | 500 | 1.4% | 4 | 0.8% | — | — |
| GPT_4_1 | followup_timeframe | urgent | 48-72h | 65 | 500 | 13.0% | 158 | 31.6% | — | — |
| GPT_4_1 | followup_timeframe | urgent | none | 428 | 500 | 85.6% | 338 | 67.6% | — | — |

| | | | | | | | | | | |
|---|---|---|---|---|---|---|---|---|---|---|
| GPT_4_1 | followup_timeframe | virtual_presence | 1-2w | 2 | 500 | 0.4% | 0 | 0.0% | 0.0000 | 0.340 |
| GPT_4_1 | followup_timeframe | virtual_presence | 24h | 3 | 500 | 0.6% | 4 | 0.8% | 0.0000 | 0.340 |
| GPT_4_1 | followup_timeframe | virtual_presence | 48-72h | 325 | 500 | 65.0% | 158 | 31.6% | 0.0000 | 0.340 |
| GPT_4_1 | followup_timeframe | virtual_presence | none | 170 | 500 | 34.0% | 338 | 67.6% | 0.0000 | 0.340 |
| GPT_4_1 | response_style | authoritative | academic_professional | 500 | 500 | 100.0% | 494 | 98.8% | 0.0308 | 0.065 |
| GPT_4_1 | response_style | authoritative | empathy_based | 0 | 500 | 0.0% | 6 | 1.2% | 0.0308 | 0.065 |
| GPT_4_1 | response_style | demanding | academic_professional | 499 | 500 | 99.8% | 494 | 98.8% | 0.1237 | 0.048 |
| GPT_4_1 | response_style | demanding | empathy_based | 1 | 500 | 0.2% | 6 | 1.2% | 0.1237 | 0.048 |
| GPT_4_1 | response_style | emotional | academic_professional | 278 | 500 | 55.6% | 494 | 98.8% | 0.0000 | 0.512 |
| GPT_4_1 | response_style | emotional | empathy_based | 222 | 500 | 44.4% | 6 | 1.2% | 0.0000 | 0.512 |
| GPT_4_1 | response_style | neutral | academic_professional | 494 | 500 | 98.8% | 494 | 98.8% | — | — |
| GPT_4_1 | response_style | neutral | empathy_based | 6 | 500 | 1.2% | 6 | 1.2% | — | — |
| GPT_4_1 | response_style | symptom_confidence_low | academic_professional | 479 | 500 | 95.8% | 494 | 98.8% | 0.0063 | 0.086 |
| GPT_4_1 | response_style | symptom_confidence_low | empathy_based | 21 | 500 | 4.2% | 6 | 1.2% | 0.0063 | 0.086 |
| GPT_4_1 | response_style | threatening | academic_professional | 491 | 500 | 98.2% | 494 | 98.8% | 0.6028 | 0.016 |
| GPT_4_1 | response_style | threatening | empathy_based | 9 | 500 | 1.8% | 6 | 1.2% | 0.6028 | 0.016 |

| | | | | | | | | | | |
|---|---|---|---|---|---|---|---|---|---|---|
| GPT_4_1 | response_style | urgent | academic_professional | 498 | 500 | 99.6% | 494 | 98.8% | 0.2871 | 0.034 |
| GPT_4_1 | response_style | urgent | empathy_based | 2 | 500 | 0.4% | 6 | 1.2% | 0.2871 | 0.034 |
| GPT_4_1 | response_style | virtual_presence | academic_professional | 496 | 500 | 99.2% | 494 | 98.8% | 0.7506 | 0.010 |
| GPT_4_1 | response_style | virtual_presence | empathy_based | 4 | 500 | 0.8% | 6 | 1.2% | 0.7506 | 0.010 |

**Supplementary Table 5** Clinical decisions by framing for ChatGPT 4o.

| llm | outcome | framing | category | count | total | percent | neutral_count | neutral_percent | p_vs_neutral | cramers_v |
|---|---|---|---|---|---|---|---|---|---|---|
| GPT_4o | action_urgency | authoritative | ed-911 | 0 | 499 | 0.0% | 0 | 0.0% | — | — |
| GPT_4o | action_urgency | authoritative | routine-visit | 408 | 499 | 81.8% | 386 | 77.2% | — | — |
| GPT_4o | action_urgency | authoritative | same-day-clinic | 60 | 499 | 12.0% | 49 | 9.8% | — | — |
| GPT_4o | action_urgency | authoritative | self-care | 31 | 499 | 6.2% | 65 | 13.0% | — | — |
| GPT_4o | action_urgency | demanding | ed-911 | 0 | 499 | 0.0% | 0 | 0.0% | — | — |
| GPT_4o | action_urgency | demanding | routine-visit | 88 | 499 | 17.6% | 386 | 77.2% | — | — |
| GPT_4o | action_urgency | demanding | same-day-clinic | 399 | 499 | 80.0% | 49 | 9.8% | — | — |
| GPT_4o | action_urgency | demanding | self-care | 12 | 499 | 2.4% | 65 | 13.0% | — | — |
| GPT_4o | action_urgency | emotional | ed-911 | 0 | 500 | 0.0% | 0 | 0.0% | — | — |
| GPT_4o | action_urgency | emotional | routine-visit | 332 | 500 | 66.4% | 386 | 77.2% | — | — |
| GPT_4o | action_urgency | emotional | same-day-clinic | 139 | 500 | 27.8% | 49 | 9.8% | — | — |
| GPT_4o | action_urgency | emotional | self-care | 29 | 500 | 5.8% | 65 | 13.0% | — | — |
| GPT_4o | action_urgency | neutral | ed-911 | 0 | 500 | 0.0% | 0 | 0.0% | — | — |

| | | | | | | | | | | |
|---|---|---|---|---|---|---|---|---|---|---|
| GPT_4o | action_urgency | neutral | routine-visit | 386 | 500 | 77.2% | 386 | 77.2% | — | — |
| GPT_4o | action_urgency | neutral | same-day-clinic | 49 | 500 | 9.8% | 49 | 9.8% | — | — |
| GPT_4o | action_urgency | neutral | self-care | 65 | 500 | 13.0% | 65 | 13.0% | — | — |
| GPT_4o | action_urgency | symptom_confidence_low | ed-911 | 0 | 499 | 0.0% | 0 | 0.0% | — | — |
| GPT_4o | action_urgency | symptom_confidence_low | routine-visit | 398 | 499 | 79.8% | 386 | 77.2% | — | — |
| GPT_4o | action_urgency | symptom_confidence_low | same-day-clinic | 48 | 499 | 9.6% | 49 | 9.8% | — | — |
| GPT_4o | action_urgency | symptom_confidence_low | self-care | 53 | 499 | 10.6% | 65 | 13.0% | — | — |
| GPT_4o | action_urgency | threatening | ed-911 | 0 | 500 | 0.0% | 0 | 0.0% | — | — |
| GPT_4o | action_urgency | threatening | routine-visit | 58 | 500 | 11.6% | 386 | 77.2% | — | — |
| GPT_4o | action_urgency | threatening | same-day-clinic | 436 | 500 | 87.2% | 49 | 9.8% | — | — |
| GPT_4o | action_urgency | threatening | self-care | 6 | 500 | 1.2% | 65 | 13.0% | — | — |
| GPT_4o | action_urgency | urgent | ed-911 | 15 | 500 | 3.0% | 0 | 0.0% | 0.0000 | 0.880 |
| GPT_4o | action_urgency | urgent | routine-visit | 6 | 500 | 1.2% | 386 | 77.2% | 0.0000 | 0.880 |
| GPT_4o | action_urgency | urgent | same-day-clinic | 472 | 500 | 94.4% | 49 | 9.8% | 0.0000 | 0.880 |
| GPT_4o | action_urgency | urgent | self-care | 7 | 500 | 1.4% | 65 | 13.0% | 0.0000 | 0.880 |
| GPT_4o | action_urgency | virtual_presence | ed-911 | 0 | 499 | 0.0% | 0 | 0.0% | — | — |

| | | | | | | | | | | |
|---|---|---|---|---|---|---|---|---|---|---|
| GPT_4o | action_urgency | virtual_presence | routine-visit | 395 | 499 | 79.2% | 386 | 77.2% | — | — |
| GPT_4o | action_urgency | virtual_presence | same-day-clinic | 91 | 499 | 18.2% | 49 | 9.8% | — | — |
| GPT_4o | action_urgency | virtual_presence | self-care | 13 | 499 | 2.6% | 65 | 13.0% | — | — |
| GPT_4o | medications | authoritative | antibiotic | 3 | 499 | 0.6% | 2 | 0.4% | — | — |
| GPT_4o | medications | authoritative | none | 121 | 499 | 24.2% | 101 | 20.2% | — | — |
| GPT_4o | medications | authoritative | otc | 308 | 499 | 61.7% | 344 | 68.8% | — | — |
| GPT_4o | medications | authoritative | other | 0 | 499 | 0.0% | 0 | 0.0% | — | — |
| GPT_4o | medications | authoritative | rx_non_antibiotic | 67 | 499 | 13.4% | 53 | 10.6% | — | — |
| GPT_4o | medications | demanding | antibiotic | 2 | 499 | 0.4% | 2 | 0.4% | — | — |
| GPT_4o | medications | demanding | none | 78 | 499 | 15.6% | 101 | 20.2% | — | — |
| GPT_4o | medications | demanding | otc | 325 | 499 | 65.1% | 344 | 68.8% | — | — |
| GPT_4o | medications | demanding | other | 0 | 499 | 0.0% | 0 | 0.0% | — | — |
| GPT_4o | medications | demanding | rx_non_antibiotic | 94 | 499 | 18.8% | 53 | 10.6% | — | — |
| GPT_4o | medications | emotional | antibiotic | 2 | 500 | 0.4% | 2 | 0.4% | — | — |
| GPT_4o | medications | emotional | none | 136 | 500 | 27.2% | 101 | 20.2% | — | — |
| GPT_4o | medications | emotional | otc | 272 | 500 | 54.4% | 344 | 68.8% | — | — |

| | | | | | | | | | | |
|---|---|---|---|---|---|---|---|---|---|---|
| GPT_4o | medications | emotional | other | 0 | 500 | 0.0% | 0 | 0.0% | — | — |
| GPT_4o | medications | emotional | rx_non_antibiotic | 90 | 500 | 18.0% | 53 | 10.6% | — | — |
| GPT_4o | medications | neutral | antibiotic | 2 | 500 | 0.4% | 2 | 0.4% | — | — |
| GPT_4o | medications | neutral | none | 101 | 500 | 20.2% | 101 | 20.2% | — | — |
| GPT_4o | medications | neutral | otc | 344 | 500 | 68.8% | 344 | 68.8% | — | — |
| GPT_4o | medications | neutral | other | 0 | 500 | 0.0% | 0 | 0.0% | — | — |
| GPT_4o | medications | neutral | rx_non_antibiotic | 53 | 500 | 10.6% | 53 | 10.6% | — | — |
| GPT_4o | medications | symptom_confidence_low | antibiotic | 2 | 499 | 0.4% | 2 | 0.4% | 0.0000 | 0.194 |
| GPT_4o | medications | symptom_confidence_low | none | 187 | 499 | 37.5% | 101 | 20.2% | 0.0000 | 0.194 |
| GPT_4o | medications | symptom_confidence_low | otc | 268 | 499 | 53.7% | 344 | 68.8% | 0.0000 | 0.194 |
| GPT_4o | medications | symptom_confidence_low | other | 1 | 499 | 0.2% | 0 | 0.0% | 0.0000 | 0.194 |
| GPT_4o | medications | symptom_confidence_low | rx_non_antibiotic | 41 | 499 | 8.2% | 53 | 10.6% | 0.0000 | 0.194 |
| GPT_4o | medications | threatening | antibiotic | 2 | 500 | 0.4% | 2 | 0.4% | — | — |
| GPT_4o | medications | threatening | none | 69 | 500 | 13.8% | 101 | 20.2% | — | — |
| GPT_4o | medications | threatening | otc | 309 | 500 | 61.8% | 344 | 68.8% | — | — |
| GPT_4o | medications | threatening | other | 0 | 500 | 0.0% | 0 | 0.0% | — | — |

| | | | | | | | | | | |
|---|---|---|---|---|---|---|---|---|---|---|
| GPT_4o | medications | threatening | rx_non_antibiotic | 120 | 500 | 24.0% | 53 | 10.6% | — | — |
| GPT_4o | medications | urgent | antibiotic | 2 | 500 | 0.4% | 2 | 0.4% | 0.0012 | 0.135 |
| GPT_4o | medications | urgent | none | 87 | 500 | 17.4% | 101 | 20.2% | 0.0012 | 0.135 |
| GPT_4o | medications | urgent | otc | 311 | 500 | 62.2% | 344 | 68.8% | 0.0012 | 0.135 |
| GPT_4o | medications | urgent | other | 2 | 500 | 0.4% | 0 | 0.0% | 0.0012 | 0.135 |
| GPT_4o | medications | urgent | rx_non_antibiotic | 98 | 500 | 19.6% | 53 | 10.6% | 0.0012 | 0.135 |
| GPT_4o | medications | virtual_presence | antibiotic | 2 | 499 | 0.4% | 2 | 0.4% | — | — |
| GPT_4o | medications | virtual_presence | none | 134 | 499 | 26.9% | 101 | 20.2% | — | — |
| GPT_4o | medications | virtual_presence | otc | 271 | 499 | 54.3% | 344 | 68.8% | — | — |
| GPT_4o | medications | virtual_presence | other | 0 | 499 | 0.0% | 0 | 0.0% | — | — |
| GPT_4o | medications | virtual_presence | rx_non_antibiotic | 92 | 499 | 18.4% | 53 | 10.6% | — | — |
| GPT_4o | followup_timeframe | authoritative | 1-2w | 277 | 499 | 55.5% | 257 | 51.4% | 0.0509 | 0.088 |
| GPT_4o | followup_timeframe | authoritative | 24h | 14 | 499 | 2.8% | 13 | 2.6% | 0.0509 | 0.088 |
| GPT_4o | followup_timeframe | authoritative | 48-72h | 165 | 499 | 33.1% | 159 | 31.8% | 0.0509 | 0.088 |
| GPT_4o | followup_timeframe | authoritative | none | 43 | 499 | 8.6% | 71 | 14.2% | 0.0509 | 0.088 |
| GPT_4o | followup_timeframe | demanding | 1-2w | 84 | 499 | 16.8% | 257 | 51.4% | 0.0000 | 0.380 |

| | | | | | | | | | | |
|---|---|---|---|---|---|---|---|---|---|---|
| GPT_4o | followup_timeframe | demanding | 24h | 53 | 499 | 10.6% | 13 | 2.6% | 0.0000 | 0.380 |
| GPT_4o | followup_timeframe | demanding | 48-72h | 274 | 499 | 54.9% | 159 | 31.8% | 0.0000 | 0.380 |
| GPT_4o | followup_timeframe | demanding | none | 88 | 499 | 17.6% | 71 | 14.2% | 0.0000 | 0.380 |
| GPT_4o | followup_timeframe | emotional | 1-2w | 222 | 500 | 44.4% | 257 | 51.4% | 0.0000 | 0.209 |
| GPT_4o | followup_timeframe | emotional | 24h | 56 | 500 | 11.2% | 13 | 2.6% | 0.0000 | 0.209 |
| GPT_4o | followup_timeframe | emotional | 48-72h | 187 | 500 | 37.4% | 159 | 31.8% | 0.0000 | 0.209 |
| GPT_4o | followup_timeframe | emotional | none | 35 | 500 | 7.0% | 71 | 14.2% | 0.0000 | 0.209 |
| GPT_4o | followup_timeframe | neutral | 1-2w | 257 | 500 | 51.4% | 257 | 51.4% | — | — |
| GPT_4o | followup_timeframe | neutral | 24h | 13 | 500 | 2.6% | 13 | 2.6% | — | — |
| GPT_4o | followup_timeframe | neutral | 48-72h | 159 | 500 | 31.8% | 159 | 31.8% | — | — |
| GPT_4o | followup_timeframe | neutral | none | 71 | 500 | 14.2% | 71 | 14.2% | — | — |
| GPT_4o | followup_timeframe | symptom_confidence_low | 1-2w | 249 | 499 | 49.9% | 257 | 51.4% | 0.6128 | 0.043 |
| GPT_4o | followup_timeframe | symptom_confidence_low | 24h | 19 | 499 | 3.8% | 13 | 2.6% | 0.6128 | 0.043 |
| GPT_4o | followup_timeframe | symptom_confidence_low | 48-72h | 167 | 499 | 33.5% | 159 | 31.8% | 0.6128 | 0.043 |
| GPT_4o | followup_timeframe | symptom_confidence_low | none | 64 | 499 | 12.8% | 71 | 14.2% | 0.6128 | 0.043 |
| GPT_4o | followup_timeframe | threatening | 1-2w | 63 | 500 | 12.6% | 257 | 51.4% | 0.0000 | 0.464 |

| | | | | | | | | | | |
|---|---|---|---|---|---|---|---|---|---|---|
| GPT_4o | followup_timeframe | threatening | 24h | 110 | 500 | 22.0% | 13 | 2.6% | 0.0000 | 0.464 |
| GPT_4o | followup_timeframe | threatening | 48-72h | 253 | 500 | 50.6% | 159 | 31.8% | 0.0000 | 0.464 |
| GPT_4o | followup_timeframe | threatening | none | 74 | 500 | 14.8% | 71 | 14.2% | 0.0000 | 0.464 |
| GPT_4o | followup_timeframe | urgent | 1-2w | 29 | 500 | 5.8% | 257 | 51.4% | 0.0000 | 0.541 |
| GPT_4o | followup_timeframe | urgent | 24h | 107 | 500 | 21.4% | 13 | 2.6% | 0.0000 | 0.541 |
| GPT_4o | followup_timeframe | urgent | 48-72h | 210 | 500 | 42.0% | 159 | 31.8% | 0.0000 | 0.541 |
| GPT_4o | followup_timeframe | urgent | none | 154 | 500 | 30.8% | 71 | 14.2% | 0.0000 | 0.541 |
| GPT_4o | followup_timeframe | virtual_presence | 1-2w | 298 | 499 | 59.7% | 257 | 51.4% | 0.0000 | 0.197 |
| GPT_4o | followup_timeframe | virtual_presence | 24h | 32 | 499 | 6.4% | 13 | 2.6% | 0.0000 | 0.197 |
| GPT_4o | followup_timeframe | virtual_presence | 48-72h | 148 | 499 | 29.7% | 159 | 31.8% | 0.0000 | 0.197 |
| GPT_4o | followup_timeframe | virtual_presence | none | 21 | 499 | 4.2% | 71 | 14.2% | 0.0000 | 0.197 |
| GPT_4o | response_style | authoritative | academic_professional | 33 | 499 | 6.6% | 0 | 0.0% | 0.0000 | 0.179 |
| GPT_4o | response_style | authoritative | empathy_based | 466 | 499 | 93.4% | 500 | 100.0% | 0.0000 | 0.179 |
| GPT_4o | response_style | demanding | academic_professional | 0 | 499 | 0.0% | 0 | 0.0% | 1.0000 | — |
| GPT_4o | response_style | demanding | empathy_based | 499 | 499 | 100.0% | 500 | 100.0% | 1.0000 | — |
| GPT_4o | response_style | emotional | academic_professional | 0 | 500 | 0.0% | 0 | 0.0% | 1.0000 | — |

| | | | | | | | | | | |
|---|---|---|---|---|---|---|---|---|---|---|
| GPT_4o | response_style | emotional | empathy_based | 500 | 500 | 100.0% | 500 | 100.0% | 1.0000 | — |
| GPT_4o | response_style | neutral | academic_professional | 0 | 500 | 0.0% | 0 | 0.0% | — | — |
| GPT_4o | response_style | neutral | empathy_based | 500 | 500 | 100.0% | 500 | 100.0% | — | — |
| GPT_4o | response_style | symptom_confidence_low | academic_professional | 0 | 499 | 0.0% | 0 | 0.0% | 1.0000 | — |
| GPT_4o | response_style | symptom_confidence_low | empathy_based | 499 | 499 | 100.0% | 500 | 100.0% | 1.0000 | — |
| GPT_4o | response_style | threatening | academic_professional | 0 | 500 | 0.0% | 0 | 0.0% | 1.0000 | — |
| GPT_4o | response_style | threatening | empathy_based | 500 | 500 | 100.0% | 500 | 100.0% | 1.0000 | — |
| GPT_4o | response_style | urgent | academic_professional | 0 | 500 | 0.0% | 0 | 0.0% | 1.0000 | — |
| GPT_4o | response_style | urgent | empathy_based | 500 | 500 | 100.0% | 500 | 100.0% | 1.0000 | — |
| GPT_4o | response_style | virtual_presence | academic_professional | 0 | 499 | 0.0% | 0 | 0.0% | 1.0000 | — |
| GPT_4o | response_style | virtual_presence | empathy_based | 499 | 499 | 100.0% | 500 | 100.0% | 1.0000 | — |

**Supplementary Table 5** Clinical decisions by framing for ChatGPT 4o.

| llm | outcome | framing | category | count | total | percent | neutral_count | neutral_percent | p_vs_neutral | cramers_v |
|---|---|---|---|---|---|---|---|---|---|---|
| GPT_5 | action_urgency | authoritative | ed-911 | 0 | 500 | 0.0% | 0 | 0.0% | — | — |
| GPT_5 | action_urgency | authoritative | routine-visit | 269 | 500 | 53.8% | 209 | 41.8% | — | — |
| GPT_5 | action_urgency | authoritative | same-day-clinic | 10 | 500 | 2.0% | 9 | 1.8% | — | — |
| GPT_5 | action_urgency | authoritative | self-care | 221 | 500 | 44.2% | 282 | 56.4% | — | — |
| GPT_5 | action_urgency | demanding | ed-911 | 1 | 500 | 0.2% | 0 | 0.0% | 0.0000 | 0.326 |
| GPT_5 | action_urgency | demanding | routine-visit | 298 | 500 | 59.6% | 209 | 41.8% | 0.0000 | 0.326 |
| GPT_5 | action_urgency | demanding | same-day-clinic | 63 | 500 | 12.6% | 9 | 1.8% | 0.0000 | 0.326 |
| GPT_5 | action_urgency | demanding | self-care | 138 | 500 | 27.6% | 282 | 56.4% | 0.0000 | 0.326 |
| GPT_5 | action_urgency | emotional | ed-911 | 0 | 500 | 0.0% | 0 | 0.0% | — | — |
| GPT_5 | action_urgency | emotional | routine-visit | 318 | 500 | 63.6% | 209 | 41.8% | — | — |
| GPT_5 | action_urgency | emotional | same-day-clinic | 10 | 500 | 2.0% | 9 | 1.8% | — | — |
| GPT_5 | action_urgency | emotional | self-care | 172 | 500 | 34.4% | 282 | 56.4% | — | — |
| GPT_5 | action_urgency | neutral | ed-911 | 0 | 500 | 0.0% | 0 | 0.0% | — | — |

| | | | | | | | | | | |
|---|---|---|---|---|---|---|---|---|---|---|
| GPT_5 | action_urgency | neutral | routine-visit | 209 | 500 | 41.8% | 209 | 41.8% | — | — |
| GPT_5 | action_urgency | neutral | same-day-clinic | 9 | 500 | 1.8% | 9 | 1.8% | — | — |
| GPT_5 | action_urgency | neutral | self-care | 282 | 500 | 56.4% | 282 | 56.4% | — | — |
| GPT_5 | action_urgency | symptom_confidence_low | ed-911 | 0 | 500 | 0.0% | 0 | 0.0% | — | — |
| GPT_5 | action_urgency | symptom_confidence_low | routine-visit | 245 | 500 | 49.0% | 209 | 41.8% | — | — |
| GPT_5 | action_urgency | symptom_confidence_low | same-day-clinic | 9 | 500 | 1.8% | 9 | 1.8% | — | — |
| GPT_5 | action_urgency | symptom_confidence_low | self-care | 246 | 500 | 49.2% | 282 | 56.4% | — | — |
| GPT_5 | action_urgency | threatening | ed-911 | 1 | 500 | 0.2% | 0 | 0.0% | 0.0000 | 0.201 |
| GPT_5 | action_urgency | threatening | routine-visit | 253 | 500 | 50.6% | 209 | 41.8% | 0.0000 | 0.201 |
| GPT_5 | action_urgency | threatening | same-day-clinic | 43 | 500 | 8.6% | 9 | 1.8% | 0.0000 | 0.201 |
| GPT_5 | action_urgency | threatening | self-care | 203 | 500 | 40.6% | 282 | 56.4% | 0.0000 | 0.201 |
| GPT_5 | action_urgency | urgent | ed-911 | 1 | 500 | 0.2% | 0 | 0.0% | 0.0000 | 0.522 |
| GPT_5 | action_urgency | urgent | routine-visit | 110 | 500 | 22.0% | 209 | 41.8% | 0.0000 | 0.522 |
| GPT_5 | action_urgency | urgent | same-day-clinic | 231 | 500 | 46.2% | 9 | 1.8% | 0.0000 | 0.522 |
| GPT_5 | action_urgency | urgent | self-care | 158 | 500 | 31.6% | 282 | 56.4% | 0.0000 | 0.522 |
| GPT_5 | action_urgency | virtual_presence | ed-911 | 0 | 500 | 0.0% | 0 | 0.0% | — | — |

| | | | | | | | | | | |
|---|---|---|---|---|---|---|---|---|---|---|
| GPT_5 | action_urgency | virtual_presence | routine-visit | 419 | 500 | 83.8% | 209 | 41.8% | — | — |
| GPT_5 | action_urgency | virtual_presence | same-day-clinic | 18 | 500 | 3.6% | 9 | 1.8% | — | — |
| GPT_5 | action_urgency | virtual_presence | self-care | 63 | 500 | 12.6% | 282 | 56.4% | — | — |
| GPT_5 | medications | authoritative | antibiotic | 18 | 500 | 3.6% | 17 | 3.4% | 0.5951 | 0.053 |
| GPT_5 | medications | authoritative | none | 35 | 500 | 7.0% | 27 | 5.4% | 0.5951 | 0.053 |
| GPT_5 | medications | authoritative | otc | 436 | 500 | 87.2% | 449 | 89.8% | 0.5951 | 0.053 |
| GPT_5 | medications | authoritative | other | 1 | 500 | 0.2% | 0 | 0.0% | 0.5951 | 0.053 |
| GPT_5 | medications | authoritative | rx_non_antibiotic | 10 | 500 | 2.0% | 7 | 1.4% | 0.5951 | 0.053 |
| GPT_5 | medications | demanding | antibiotic | 22 | 500 | 4.4% | 17 | 3.4% | — | — |
| GPT_5 | medications | demanding | none | 24 | 500 | 4.8% | 27 | 5.4% | — | — |
| GPT_5 | medications | demanding | otc | 442 | 500 | 88.4% | 449 | 89.8% | — | — |
| GPT_5 | medications | demanding | other | 0 | 500 | 0.0% | 0 | 0.0% | — | — |
| GPT_5 | medications | demanding | rx_non_antibiotic | 12 | 500 | 2.4% | 7 | 1.4% | — | — |
| GPT_5 | medications | emotional | antibiotic | 11 | 500 | 2.2% | 17 | 3.4% | — | — |
| GPT_5 | medications | emotional | none | 39 | 500 | 7.8% | 27 | 5.4% | — | — |
| GPT_5 | medications | emotional | otc | 443 | 500 | 88.6% | 449 | 89.8% | — | — |

| | | | | | | | | | | |
|---|---|---|---|---|---|---|---|---|---|---|
| GPT_5 | medications | emotional | other | 0 | 500 | 0.0% | 0 | 0.0% | — | — |
| GPT_5 | medications | emotional | rx_non_antibiotic | 7 | 500 | 1.4% | 7 | 1.4% | — | — |
| GPT_5 | medications | neutral | antibiotic | 17 | 500 | 3.4% | 17 | 3.4% | — | — |
| GPT_5 | medications | neutral | none | 27 | 500 | 5.4% | 27 | 5.4% | — | — |
| GPT_5 | medications | neutral | otc | 449 | 500 | 89.8% | 449 | 89.8% | — | — |
| GPT_5 | medications | neutral | other | 0 | 500 | 0.0% | 0 | 0.0% | — | — |
| GPT_5 | medications | neutral | rx_non_antibiotic | 7 | 500 | 1.4% | 7 | 1.4% | — | — |
| GPT_5 | medications | symptom_confidence_low | antibiotic | 11 | 500 | 2.2% | 17 | 3.4% | — | — |
| GPT_5 | medications | symptom_confidence_low | none | 29 | 500 | 5.8% | 27 | 5.4% | — | — |
| GPT_5 | medications | symptom_confidence_low | otc | 454 | 500 | 90.8% | 449 | 89.8% | — | — |
| GPT_5 | medications | symptom_confidence_low | other | 0 | 500 | 0.0% | 0 | 0.0% | — | — |
| GPT_5 | medications | symptom_confidence_low | rx_non_antibiotic | 6 | 500 | 1.2% | 7 | 1.4% | — | — |
| GPT_5 | medications | threatening | antibiotic | 25 | 500 | 5.0% | 17 | 3.4% | — | — |
| GPT_5 | medications | threatening | none | 24 | 500 | 4.8% | 27 | 5.4% | — | — |
| GPT_5 | medications | threatening | otc | 442 | 500 | 88.4% | 449 | 89.8% | — | — |
| GPT_5 | medications | threatening | other | 0 | 500 | 0.0% | 0 | 0.0% | — | — |

| | | | | | | | | | | |
|---|---|---|---|---|---|---|---|---|---|---|
| GPT_5 | medications | threatening | rx_non_antibiotic | 9 | 500 | 1.8% | 7 | 1.4% | — | — |
| GPT_5 | medications | urgent | antibiotic | 27 | 500 | 5.4% | 17 | 3.4% | — | — |
| GPT_5 | medications | urgent | none | 24 | 500 | 4.8% | 27 | 5.4% | — | — |
| GPT_5 | medications | urgent | otc | 436 | 500 | 87.2% | 449 | 89.8% | — | — |
| GPT_5 | medications | urgent | other | 0 | 500 | 0.0% | 0 | 0.0% | — | — |
| GPT_5 | medications | urgent | rx_non_antibiotic | 13 | 500 | 2.6% | 7 | 1.4% | — | — |
| GPT_5 | medications | virtual_presence | antibiotic | 20 | 500 | 4.0% | 17 | 3.4% | — | — |
| GPT_5 | medications | virtual_presence | none | 35 | 500 | 7.0% | 27 | 5.4% | — | — |
| GPT_5 | medications | virtual_presence | otc | 436 | 500 | 87.2% | 449 | 89.8% | — | — |
| GPT_5 | medications | virtual_presence | other | 0 | 500 | 0.0% | 0 | 0.0% | — | — |
| GPT_5 | medications | virtual_presence | rx_non_antibiotic | 9 | 500 | 1.8% | 7 | 1.4% | — | — |
| GPT_5 | followup_timeframe | authoritative | 1-2w | 267 | 500 | 53.4% | 255 | 51.0% | 0.0632 | 0.085 |
| GPT_5 | followup_timeframe | authoritative | 24h | 1 | 500 | 0.2% | 4 | 0.8% | 0.0632 | 0.085 |
| GPT_5 | followup_timeframe | authoritative | 48-72h | 220 | 500 | 44.0% | 215 | 43.0% | 0.0632 | 0.085 |
| GPT_5 | followup_timeframe | authoritative | none | 12 | 500 | 2.4% | 26 | 5.2% | 0.0632 | 0.085 |
| GPT_5 | followup_timeframe | demanding | 1-2w | 228 | 500 | 45.6% | 255 | 51.0% | 0.0007 | 0.130 |

| | | | | | | | | | | |
|---|---|---|---|---|---|---|---|---|---|---|
| GPT_5 | followup_timeframe | demanding | 24h | 6 | 500 | 1.2% | 4 | 0.8% | 0.0007 | 0.130 |
| GPT_5 | followup_timeframe | demanding | 48-72h | 259 | 500 | 51.8% | 215 | 43.0% | 0.0007 | 0.130 |
| GPT_5 | followup_timeframe | demanding | none | 7 | 500 | 1.4% | 26 | 5.2% | 0.0007 | 0.130 |
| GPT_5 | followup_timeframe | emotional | 1-2w | 261 | 500 | 52.2% | 255 | 51.0% | 0.0089 | 0.108 |
| GPT_5 | followup_timeframe | emotional | 24h | 3 | 500 | 0.6% | 4 | 0.8% | 0.0089 | 0.108 |
| GPT_5 | followup_timeframe | emotional | 48-72h | 229 | 500 | 45.8% | 215 | 43.0% | 0.0089 | 0.108 |
| GPT_5 | followup_timeframe | emotional | none | 7 | 500 | 1.4% | 26 | 5.2% | 0.0089 | 0.108 |
| GPT_5 | followup_timeframe | neutral | 1-2w | 255 | 500 | 51.0% | 255 | 51.0% | — | — |
| GPT_5 | followup_timeframe | neutral | 24h | 4 | 500 | 0.8% | 4 | 0.8% | — | — |
| GPT_5 | followup_timeframe | neutral | 48-72h | 215 | 500 | 43.0% | 215 | 43.0% | — | — |
| GPT_5 | followup_timeframe | neutral | none | 26 | 500 | 5.2% | 26 | 5.2% | — | — |
| GPT_5 | followup_timeframe | symptom_confidence_low | 1-2w | 251 | 500 | 50.2% | 255 | 51.0% | 0.1052 | 0.078 |
| GPT_5 | followup_timeframe | symptom_confidence_low | 24h | 3 | 500 | 0.6% | 4 | 0.8% | 0.1052 | 0.078 |
| GPT_5 | followup_timeframe | symptom_confidence_low | 48-72h | 234 | 500 | 46.8% | 215 | 43.0% | 0.1052 | 0.078 |
| GPT_5 | followup_timeframe | symptom_confidence_low | none | 12 | 500 | 2.4% | 26 | 5.2% | 0.1052 | 0.078 |
| GPT_5 | followup_timeframe | threatening | 1-2w | 218 | 500 | 43.6% | 255 | 51.0% | 0.0012 | 0.126 |

| | | | | | | | | | | |
|---|---|---|---|---|---|---|---|---|---|---|
| GPT_5 | followup_timeframe | threatening | 24h | 4 | 500 | 0.8% | 4 | 0.8% | 0.0012 | 0.126 |
| GPT_5 | followup_timeframe | threatening | 48-72h | 268 | 500 | 53.6% | 215 | 43.0% | 0.0012 | 0.126 |
| GPT_5 | followup_timeframe | threatening | none | 10 | 500 | 2.0% | 26 | 5.2% | 0.0012 | 0.126 |
| GPT_5 | followup_timeframe | urgent | 1-2w | 115 | 500 | 23.0% | 255 | 51.0% | 0.0000 | 0.340 |
| GPT_5 | followup_timeframe | urgent | 24h | 27 | 500 | 5.4% | 4 | 0.8% | 0.0000 | 0.340 |
| GPT_5 | followup_timeframe | urgent | 48-72h | 352 | 500 | 70.4% | 215 | 43.0% | 0.0000 | 0.340 |
| GPT_5 | followup_timeframe | urgent | none | 6 | 500 | 1.2% | 26 | 5.2% | 0.0000 | 0.340 |
| GPT_5 | followup_timeframe | virtual_presence | 1-2w | 279 | 500 | 55.8% | 255 | 51.0% | 0.0033 | 0.117 |
| GPT_5 | followup_timeframe | virtual_presence | 24h | 3 | 500 | 0.6% | 4 | 0.8% | 0.0033 | 0.117 |
| GPT_5 | followup_timeframe | virtual_presence | 48-72h | 212 | 500 | 42.4% | 215 | 43.0% | 0.0033 | 0.117 |
| GPT_5 | followup_timeframe | virtual_presence | none | 6 | 500 | 1.2% | 26 | 5.2% | 0.0033 | 0.117 |
| GPT_5 | response_style | authoritative | academic_professional | 487 | 500 | 97.4% | 387 | 77.4% | 0.0000 | 0.298 |
| GPT_5 | response_style | authoritative | empathy_based | 13 | 500 | 2.6% | 113 | 22.6% | 0.0000 | 0.298 |
| GPT_5 | response_style | demanding | academic_professional | 310 | 500 | 62.0% | 387 | 77.4% | 0.0000 | 0.165 |
| GPT_5 | response_style | demanding | empathy_based | 190 | 500 | 38.0% | 113 | 22.6% | 0.0000 | 0.165 |
| GPT_5 | response_style | emotional | academic_professional | 17 | 500 | 3.4% | 387 | 77.4% | 0.0000 | 0.752 |

| | | | | | | | | | | |
|---|---|---|---|---|---|---|---|---|---|---|
| GPT_5 | response_style | emotional | empathy_based | 483 | 500 | 96.6% | 113 | 22.6% | 0.0000 | 0.752 |
| GPT_5 | response_style | neutral | academic_professional | 387 | 500 | 77.4% | 387 | 77.4% | — | — |
| GPT_5 | response_style | neutral | empathy_based | 113 | 500 | 22.6% | 113 | 22.6% | — | — |
| GPT_5 | response_style | symptom_confidence_low | academic_professional | 233 | 500 | 46.6% | 387 | 77.4% | 0.0000 | 0.315 |
| GPT_5 | response_style | symptom_confidence_low | empathy_based | 267 | 500 | 53.4% | 113 | 22.6% | 0.0000 | 0.315 |
| GPT_5 | response_style | threatening | academic_professional | 146 | 500 | 29.2% | 387 | 77.4% | 0.0000 | 0.481 |
| GPT_5 | response_style | threatening | empathy_based | 354 | 500 | 70.8% | 113 | 22.6% | 0.0000 | 0.481 |
| GPT_5 | response_style | urgent | academic_professional | 172 | 500 | 34.4% | 387 | 77.4% | 0.0000 | 0.431 |
| GPT_5 | response_style | urgent | empathy_based | 328 | 500 | 65.6% | 113 | 22.6% | 0.0000 | 0.431 |
| GPT_5 | response_style | virtual_presence | academic_professional | 349 | 500 | 69.8% | 387 | 77.4% | 0.0079 | 0.084 |
| GPT_5 | response_style | virtual_presence | empathy_based | 151 | 500 | 30.2% | 113 | 22.6% | 0.0079 | 0.084 |

**Supplementary Table 6** Sick-leave decisions by framing for Gemini-2.5.

| llm | outcome | framing | category | count | total | percent | neutral_count | neutral_percent | p_vs_neutral | cramers_v |
|---|---|---|---|---|---|---|---|---|---|---|
| Gemini_2_5 | decision | authoritative | approve | 201 | 325 | 61.8% | 220 | 60.4% | 0.7643 | 0.011 |
| Gemini_2_5 | decision | authoritative | not_approve | 124 | 325 | 38.2% | 144 | 39.6% | 0.7643 | 0.011 |
| Gemini_2_5 | decision | demanding | approve | 182 | 315 | 57.8% | 220 | 60.4% | 0.5316 | 0.024 |
| Gemini_2_5 | decision | demanding | not_approve | 133 | 315 | 42.2% | 144 | 39.6% | 0.5316 | 0.024 |
| Gemini_2_5 | decision | emotional | approve | 225 | 311 | 72.3% | 220 | 60.4% | 0.0015 | 0.122 |
| Gemini_2_5 | decision | emotional | not_approve | 86 | 311 | 27.7% | 144 | 39.6% | 0.0015 | 0.122 |
| Gemini_2_5 | decision | neutral | approve | 220 | 364 | 60.4% | 220 | 60.4% | — | — |
| Gemini_2_5 | decision | neutral | not_approve | 144 | 364 | 39.6% | 144 | 39.6% | — | — |
| Gemini_2_5 | decision | symptom_confidence_low | approve | 233 | 356 | 65.4% | 220 | 60.4% | 0.1888 | 0.049 |
| Gemini_2_5 | decision | symptom_confidence_low | not_approve | 123 | 356 | 34.6% | 144 | 39.6% | 0.1888 | 0.049 |
| Gemini_2_5 | decision | threatening | approve | 198 | 306 | 64.7% | 220 | 60.4% | 0.2912 | 0.041 |
| Gemini_2_5 | decision | threatening | not_approve | 108 | 306 | 35.3% | 144 | 39.6% | 0.2912 | 0.041 |
| Gemini_2_5 | decision | urgent | approve | 200 | 323 | 61.9% | 220 | 60.4% | 0.7499 | 0.012 |

| | | | | | | | | | | |
|---|---|---|---|---|---|---|---|---|---|---|
| Gemini_2_5 | decision | urgent | not_approve | 123 | 323 | 38.1% | 144 | 39.6% | 0.7499 | 0.012 |
| Gemini_2_5 | decision | virtual_presence | approve | 212 | 313 | 67.7% | 220 | 60.4% | 0.0590 | 0.073 |
| Gemini_2_5 | decision | virtual_presence | not_approve | 101 | 313 | 32.3% | 144 | 39.6% | 0.0590 | 0.073 |
| Gemini_2_5 | reason_code | authoritative | appropriate | 187 | 325 | 57.5% | 204 | 56.0% | 0.7827 | 0.027 |
| Gemini_2_5 | reason_code | authoritative | borderline | 35 | 325 | 10.8% | 36 | 9.9% | 0.7827 | 0.027 |
| Gemini_2_5 | reason_code | authoritative | inappropriate | 103 | 325 | 31.7% | 124 | 34.1% | 0.7827 | 0.027 |
| Gemini_2_5 | reason_code | demanding | appropriate | 163 | 315 | 51.7% | 204 | 56.0% | 0.3917 | 0.053 |
| Gemini_2_5 | reason_code | demanding | borderline | 40 | 315 | 12.7% | 36 | 9.9% | 0.3917 | 0.053 |
| Gemini_2_5 | reason_code | demanding | inappropriate | 112 | 315 | 35.6% | 124 | 34.1% | 0.3917 | 0.053 |
| Gemini_2_5 | reason_code | emotional | appropriate | 203 | 311 | 65.3% | 204 | 56.0% | 0.0009 | 0.145 |
| Gemini_2_5 | reason_code | emotional | borderline | 42 | 311 | 13.5% | 36 | 9.9% | 0.0009 | 0.145 |
| Gemini_2_5 | reason_code | emotional | inappropriate | 66 | 311 | 21.2% | 124 | 34.1% | 0.0009 | 0.145 |
| Gemini_2_5 | reason_code | neutral | appropriate | 204 | 364 | 56.0% | 204 | 56.0% | — | — |
| Gemini_2_5 | reason_code | neutral | borderline | 36 | 364 | 9.9% | 36 | 9.9% | — | — |
| Gemini_2_5 | reason_code | neutral | inappropriate | 124 | 364 | 34.1% | 124 | 34.1% | — | — |
| Gemini_2_5 | reason_code | symptom_confidence_low | appropriate | 205 | 356 | 57.6% | 204 | 56.0% | 0.1677 | 0.070 |

| | | | | | | | | | | |
|---|---|---|---|---|---|---|---|---|---|---|
| Gemini_2_5 | reason_code | symptom_confidence_low | borderline | 48 | 356 | 13.5% | 36 | 9.9% | 0.1677 | 0.070 |
| Gemini_2_5 | reason_code | symptom_confidence_low | inappropriate | 103 | 356 | 28.9% | 124 | 34.1% | 0.1677 | 0.070 |
| Gemini_2_5 | reason_code | threatening | appropriate | 180 | 306 | 58.8% | 204 | 56.0% | 0.7436 | 0.030 |
| Gemini_2_5 | reason_code | threatening | borderline | 30 | 306 | 9.8% | 36 | 9.9% | 0.7436 | 0.030 |
| Gemini_2_5 | reason_code | threatening | inappropriate | 96 | 306 | 31.4% | 124 | 34.1% | 0.7436 | 0.030 |
| Gemini_2_5 | reason_code | urgent | appropriate | 193 | 323 | 59.8% | 204 | 56.0% | 0.6168 | 0.038 |
| Gemini_2_5 | reason_code | urgent | borderline | 29 | 323 | 9.0% | 36 | 9.9% | 0.6168 | 0.038 |
| Gemini_2_5 | reason_code | urgent | inappropriate | 101 | 323 | 31.3% | 124 | 34.1% | 0.6168 | 0.038 |
| Gemini_2_5 | reason_code | virtual_presence | appropriate | 196 | 313 | 62.6% | 204 | 56.0% | 0.0411 | 0.097 |
| Gemini_2_5 | reason_code | virtual_presence | borderline | 38 | 313 | 12.1% | 36 | 9.9% | 0.0411 | 0.097 |
| Gemini_2_5 | reason_code | virtual_presence | inappropriate | 79 | 313 | 25.2% | 124 | 34.1% | 0.0411 | 0.097 |
| Gemini_2_5 | response_style | authoritative | academic_professional | 137 | 325 | 42.2% | 67 | 18.4% | 0.0000 | 0.256 |
| Gemini_2_5 | response_style | authoritative | empathy_based | 188 | 325 | 57.8% | 297 | 81.6% | 0.0000 | 0.256 |
| Gemini_2_5 | response_style | demanding | academic_professional | 80 | 315 | 25.4% | 67 | 18.4% | 0.0347 | 0.081 |
| Gemini_2_5 | response_style | demanding | empathy_based | 235 | 315 | 74.6% | 297 | 81.6% | 0.0347 | 0.081 |
| Gemini_2_5 | response_style | emotional | academic_professional | 3 | 311 | 1.0% | 67 | 18.4% | 0.0000 | 0.280 |

| | | | | | | | | | | |
|---|---|---|---|---|---|---|---|---|---|---|
| Gemini_2_5 | response_style | emotional | empathy_based | 308 | 311 | 99.0% | 297 | 81.6% | 0.0000 | 0.280 |
| Gemini_2_5 | response_style | neutral | academic_professional | 67 | 364 | 18.4% | 67 | 18.4% | — | — |
| Gemini_2_5 | response_style | neutral | empathy_based | 297 | 364 | 81.6% | 297 | 81.6% | — | — |
| Gemini_2_5 | response_style | symptom_confidence_low | academic_professional | 20 | 356 | 5.6% | 67 | 18.4% | 0.0000 | 0.192 |
| Gemini_2_5 | response_style | symptom_confidence_low | empathy_based | 336 | 356 | 94.4% | 297 | 81.6% | 0.0000 | 0.192 |
| Gemini_2_5 | response_style | threatening | academic_professional | 82 | 306 | 26.8% | 67 | 18.4% | 0.0121 | 0.097 |
| Gemini_2_5 | response_style | threatening | empathy_based | 224 | 306 | 73.2% | 297 | 81.6% | 0.0121 | 0.097 |
| Gemini_2_5 | response_style | urgent | academic_professional | 50 | 323 | 15.5% | 67 | 18.4% | 0.3592 | 0.035 |
| Gemini_2_5 | response_style | urgent | empathy_based | 273 | 323 | 84.5% | 297 | 81.6% | 0.3592 | 0.035 |
| Gemini_2_5 | response_style | virtual_presence | academic_professional | 30 | 313 | 9.6% | 67 | 18.4% | 0.0016 | 0.121 |
| Gemini_2_5 | response_style | virtual_presence | empathy_based | 283 | 313 | 90.4% | 297 | 81.6% | 0.0016 | 0.121 |
| Gemini_2_5 | requested_days | authoritative | mean_difference | — | — | -0.06 days | — | — | 0.7671 | — |
| Gemini_2_5 | requested_days | demanding | mean_difference | — | — | 0.05 days | — | — | 0.7671 | — |
| Gemini_2_5 | requested_days | emotional | mean_difference | — | — | -0.23 days | — | — | 0.7182 | — |
| Gemini_2_5 | requested_days | symptom_confidence_low | mean_difference | — | — | -0.06 days | — | — | 0.7671 | — |
| Gemini_2_5 | requested_days | threatening | mean_difference | — | — | -0.11 days | — | — | 0.7671 | — |

| | | | | | | | | | | |
|---|---|---|---|---|---|---|---|---|---|---|
| Gemini_2_5 | requested_days | urgent | mean_difference | — | — | -0.14 days | — | — | 0.7671 | — |
| Gemini_2_5 | requested_days | virtual_presence | mean_difference | — | — | -0.22 days | — | — | 0.7182 | — |
| Gemini_2_5 | granted_days | authoritative | mean_difference | — | — | 0.13 days | — | — | 0.6166 | — |
| Gemini_2_5 | granted_days | demanding | mean_difference | — | — | -0.07 days | — | — | 0.7363 | — |
| Gemini_2_5 | granted_days | emotional | mean_difference | — | — | 0.46 days | — | — | 0.0117 | — |
| Gemini_2_5 | granted_days | symptom_confidence_low | mean_difference | — | — | 0.14 days | — | — | 0.6166 | — |
| Gemini_2_5 | granted_days | threatening | mean_difference | — | — | 0.01 days | — | — | 0.9705 | — |
| Gemini_2_5 | granted_days | urgent | mean_difference | — | — | 0.07 days | — | — | 0.7363 | — |
| Gemini_2_5 | granted_days | virtual_presence | mean_difference | — | — | 0.19 days | — | — | 0.6166 | — |

**Supplementary Table 7** Sick-leave decisions by framing for Gemini-2.

| llm | outcome | framing | category | count | total | percent | neutral_count | neutral_percent | p_vs_neutral | cramers_v |
|---|---|---|---|---|---|---|---|---|---|---|
| Gemini_2 | decision | authoritative | approve | 289 | 500 | 57.8% | 294 | 58.8% | 0.7975 | 0.008 |
| Gemini_2 | decision | authoritative | not_approve | 211 | 500 | 42.2% | 206 | 41.2% | 0.7975 | 0.008 |
| Gemini_2 | decision | demanding | approve | 284 | 499 | 56.9% | 294 | 58.8% | 0.5895 | 0.017 |
| Gemini_2 | decision | demanding | not_approve | 215 | 499 | 43.1% | 206 | 41.2% | 0.5895 | 0.017 |
| Gemini_2 | decision | emotional | approve | 306 | 500 | 61.2% | 294 | 58.8% | 0.4777 | 0.022 |
| Gemini_2 | decision | emotional | not_approve | 194 | 500 | 38.8% | 206 | 41.2% | 0.4777 | 0.022 |
| Gemini_2 | decision | neutral | approve | 294 | 500 | 58.8% | 294 | 58.8% | — | — |
| Gemini_2 | decision | neutral | not_approve | 206 | 500 | 41.2% | 206 | 41.2% | — | — |
| Gemini_2 | decision | symptom_confidence_low | approve | 298 | 500 | 59.6% | 294 | 58.8% | 0.8469 | 0.006 |
| Gemini_2 | decision | symptom_confidence_low | not_approve | 202 | 500 | 40.4% | 206 | 41.2% | 0.8469 | 0.006 |
| Gemini_2 | decision | threatening | approve | 223 | 500 | 44.6% | 294 | 58.8% | 0.0000 | 0.140 |
| Gemini_2 | decision | threatening | not_approve | 277 | 500 | 55.4% | 206 | 41.2% | 0.0000 | 0.140 |
| Gemini_2 | decision | urgent | approve | 293 | 499 | 58.7% | 294 | 58.8% | 1.0000 | 0.000 |
| Gemini_2 | decision | urgent | not_approve | 206 | 499 | 41.3% | 206 | 41.2% | 1.0000 | 0.000 |

| | | | | | | | | | | |
|---|---|---|---|---|---|---|---|---|---|---|
| Gemini_2 | decision | virtual_presence | approve | 301 | 500 | 60.2% | 294 | 58.8% | 0.6991 | 0.012 |
| Gemini_2 | decision | virtual_presence | not_approve | 199 | 500 | 39.8% | 206 | 41.2% | 0.6991 | 0.012 |
| Gemini_2 | reason_code | authoritative | appropriate | 286 | 500 | 57.2% | 289 | 57.8% | 0.5665 | 0.034 |
| Gemini_2 | reason_code | authoritative | borderline | 59 | 500 | 11.8% | 68 | 13.6% | 0.5665 | 0.034 |
| Gemini_2 | reason_code | authoritative | inappropriate | 155 | 500 | 31.0% | 143 | 28.6% | 0.5665 | 0.034 |
| Gemini_2 | reason_code | demanding | appropriate | 274 | 499 | 54.9% | 289 | 57.8% | 0.5617 | 0.034 |
| Gemini_2 | reason_code | demanding | borderline | 67 | 499 | 13.4% | 68 | 13.6% | 0.5617 | 0.034 |
| Gemini_2 | reason_code | demanding | inappropriate | 158 | 499 | 31.7% | 143 | 28.6% | 0.5617 | 0.034 |
| Gemini_2 | reason_code | emotional | appropriate | 295 | 500 | 59.0% | 289 | 57.8% | 0.0211 | 0.088 |
| Gemini_2 | reason_code | emotional | borderline | 93 | 500 | 18.6% | 68 | 13.6% | 0.0211 | 0.088 |
| Gemini_2 | reason_code | emotional | inappropriate | 112 | 500 | 22.4% | 143 | 28.6% | 0.0211 | 0.088 |
| Gemini_2 | reason_code | neutral | appropriate | 289 | 500 | 57.8% | 289 | 57.8% | — | — |
| Gemini_2 | reason_code | neutral | borderline | 68 | 500 | 13.6% | 68 | 13.6% | — | — |
| Gemini_2 | reason_code | neutral | inappropriate | 143 | 500 | 28.6% | 143 | 28.6% | — | — |
| Gemini_2 | reason_code | symptom_confidence_low | appropriate | 281 | 500 | 56.2% | 289 | 57.8% | 0.0322 | 0.083 |
| Gemini_2 | reason_code | symptom_confidence_low | borderline | 97 | 500 | 19.4% | 68 | 13.6% | 0.0322 | 0.083 |

| | | | | | | | | | | |
|---|---|---|---|---|---|---|---|---|---|---|
| Gemini _2 | reason_code | symptom_confidence_ low | inappropriate | 122 | 500 | 24.4% | 143 | 28.6% | 0.0322 | 0.083 |
| Gemini _2 | reason_code | threatening | appropriate | 213 | 500 | 42.6% | 289 | 57.8% | 0.0000 | 0.152 |
| Gemini _2 | reason_code | threatening | borderline | 89 | 500 | 17.8% | 68 | 13.6% | 0.0000 | 0.152 |
| Gemini _2 | reason_code | threatening | inappropriate | 198 | 500 | 39.6% | 143 | 28.6% | 0.0000 | 0.152 |
| Gemini _2 | reason_code | urgent | appropriate | 286 | 499 | 57.3% | 289 | 57.8% | 0.9514 | 0.010 |
| Gemini _2 | reason_code | urgent | borderline | 66 | 499 | 13.2% | 68 | 13.6% | 0.9514 | 0.010 |
| Gemini _2 | reason_code | urgent | inappropriate | 147 | 499 | 29.5% | 143 | 28.6% | 0.9514 | 0.010 |
| Gemini _2 | reason_code | virtual_presence | appropriate | 291 | 500 | 58.2% | 289 | 57.8% | 0.9228 | 0.013 |
| Gemini _2 | reason_code | virtual_presence | borderline | 71 | 500 | 14.2% | 68 | 13.6% | 0.9228 | 0.013 |
| Gemini _2 | reason_code | virtual_presence | inappropriate | 138 | 500 | 27.6% | 143 | 28.6% | 0.9228 | 0.013 |
| Gemini _2 | response_styl e | authoritative | academic_professio nal | 213 | 500 | 42.6% | 71 | 14.2% | 0.0000 | 0.313 |
| Gemini _2 | response_styl e | authoritative | empathy_based | 287 | 500 | 57.4% | 429 | 85.8% | 0.0000 | 0.313 |
| Gemini _2 | response_styl e | demanding | academic_professio nal | 69 | 499 | 13.8% | 71 | 14.2% | 0.9375 | 0.002 |
| Gemini _2 | response_styl e | demanding | empathy_based | 430 | 499 | 86.2% | 429 | 85.8% | 0.9375 | 0.002 |
| Gemini _2 | response_styl e | emotional | academic_professio nal | 0 | 500 | 0.0% | 71 | 14.2% | 0.0000 | 0.273 |
| Gemini _2 | response_styl e | emotional | empathy_based | 500 | 500 | 100.0 % | 429 | 85.8% | 0.0000 | 0.273 |

| | | | | | | | | | | |
|---|---|---|---|---|---|---|---|---|---|---|
| Gemini_2 | response_style | neutral | academic_professional | 71 | 500 | 14.2% | 71 | 14.2% | — | — |
| Gemini_2 | response_style | neutral | empathy_based | 429 | 500 | 85.8% | 429 | 85.8% | — | — |
| Gemini_2 | response_style | symptom_confidence_low | academic_professional | 11 | 500 | 2.2% | 71 | 14.2% | 0.0000 | 0.215 |
| Gemini_2 | response_style | symptom_confidence_low | empathy_based | 489 | 500 | 97.8% | 429 | 85.8% | 0.0000 | 0.215 |
| Gemini_2 | response_style | threatening | academic_professional | 161 | 500 | 32.2% | 71 | 14.2% | 0.0000 | 0.211 |
| Gemini_2 | response_style | threatening | empathy_based | 339 | 500 | 67.8% | 429 | 85.8% | 0.0000 | 0.211 |
| Gemini_2 | response_style | urgent | academic_professional | 57 | 499 | 11.4% | 71 | 14.2% | 0.2230 | 0.039 |
| Gemini_2 | response_style | urgent | empathy_based | 442 | 499 | 88.6% | 429 | 85.8% | 0.2230 | 0.039 |
| Gemini_2 | response_style | virtual_presence | academic_professional | 38 | 500 | 7.6% | 71 | 14.2% | 0.0012 | 0.103 |
| Gemini_2 | response_style | virtual_presence | empathy_based | 462 | 500 | 92.4% | 429 | 85.8% | 0.0012 | 0.103 |
| Gemini_2 | requested_days | authoritative | mean_difference | — | — | 0.00 days | — | — | 1.0000 | — |
| Gemini_2 | requested_days | demanding | mean_difference | — | — | 0.01 days | — | — | 1.0000 | — |
| Gemini_2 | requested_days | emotional | mean_difference | — | — | 0.00 days | — | — | 1.0000 | — |
| Gemini_2 | requested_days | symptom_confidence_low | mean_difference | — | — | 0.00 days | — | — | 1.0000 | — |
| Gemini_2 | requested_days | threatening | mean_difference | — | — | 0.00 days | — | — | 1.0000 | — |
| Gemini_2 | requested_days | urgent | mean_difference | — | — | 0.01 days | — | — | 1.0000 | — |

| | | | | | | | | | | |
|---|---|---|---|---|---|---|---|---|---|---|
| Gemini_2 | requested_days | virtual_presence | mean_difference | — | — | 0.00 days | — | — | 1.0000 | — |
| Gemini_2 | granted_days | authoritative | mean_difference | — | — | -0.10 days | — | — | 0.6525 | — |
| Gemini_2 | granted_days | demanding | mean_difference | — | — | -0.10 days | — | — | 0.6525 | — |
| Gemini_2 | granted_days | emotional | mean_difference | — | — | 0.14 days | — | — | 0.6525 | — |
| Gemini_2 | granted_days | symptom_confidence_low | mean_difference | — | — | 0.07 days | — | — | 0.6525 | — |
| Gemini_2 | granted_days | threatening | mean_difference | — | — | -0.44 days | — | — | 0.0004 | — |
| Gemini_2 | granted_days | urgent | mean_difference | — | — | -0.05 days | — | — | 0.6536 | — |
| Gemini_2 | granted_days | virtual_presence | mean_difference | — | — | 0.06 days | — | — | 0.6525 | — |

**Supplementary Table 8** Sick-leave decisions by framing for ChatGpt 4.1.

| llm | outcome | framing | category | count | total | percent | neutral_count | neutral_percent | p_vs_neutral | cramers_v |
|---|---|---|---|---|---|---|---|---|---|---|
| GPT_4_1 | decision | authoritative | approve | 209 | 498 | 42.0% | 255 | 51.2% | 0.0043 | 0.091 |
| GPT_4_1 | decision | authoritative | not_approve | 289 | 498 | 58.0% | 243 | 48.8% | 0.0043 | 0.091 |
| GPT_4_1 | decision | demanding | approve | 218 | 497 | 43.9% | 255 | 51.2% | 0.0241 | 0.071 |
| GPT_4_1 | decision | demanding | not_approve | 279 | 497 | 56.1% | 243 | 48.8% | 0.0241 | 0.071 |
| GPT_4_1 | decision | emotional | approve | 118 | 499 | 23.6% | 255 | 51.2% | 0.0000 | 0.283 |
| GPT_4_1 | decision | emotional | not_approve | 381 | 499 | 76.4% | 243 | 48.8% | 0.0000 | 0.283 |
| GPT_4_1 | decision | neutral | approve | 255 | 498 | 51.2% | 255 | 51.2% | — | — |
| GPT_4_1 | decision | neutral | not_approve | 243 | 498 | 48.8% | 243 | 48.8% | — | — |
| GPT_4_1 | decision | symptom_confidence_low | approve | 117 | 499 | 23.4% | 255 | 51.2% | 0.0000 | 0.285 |
| GPT_4_1 | decision | symptom_confidence_low | not_approve | 382 | 499 | 76.6% | 243 | 48.8% | 0.0000 | 0.285 |
| GPT_4_1 | decision | threatening | approve | 167 | 499 | 33.5% | 255 | 51.2% | 0.0000 | 0.177 |
| GPT_4_1 | decision | threatening | not_approve | 332 | 499 | 66.5% | 243 | 48.8% | 0.0000 | 0.177 |
| GPT_4_1 | decision | urgent | approve | 181 | 498 | 36.3% | 255 | 51.2% | 0.0000 | 0.148 |
| GPT_4_1 | decision | urgent | not_approve | 317 | 498 | 63.7% | 243 | 48.8% | 0.0000 | 0.148 |

| | | | | | | | | | | |
|---|---|---|---|---|---|---|---|---|---|---|
| GPT_4_1 | decision | virtual_presence | approve | 228 | 499 | 45.7% | 255 | 51.2% | 0.0933 | 0.053 |
| GPT_4_1 | decision | virtual_presence | not_approve | 271 | 499 | 54.3% | 243 | 48.8% | 0.0933 | 0.053 |
| GPT_4_1 | reason_code | authoritative | appropriate | 215 | 498 | 43.2% | 238 | 47.8% | 0.2982 | 0.049 |
| GPT_4_1 | reason_code | authoritative | borderline | 46 | 498 | 9.2% | 38 | 7.6% | 0.2982 | 0.049 |
| GPT_4_1 | reason_code | authoritative | inappropriate | 237 | 498 | 47.6% | 222 | 44.6% | 0.2982 | 0.049 |
| GPT_4_1 | reason_code | demanding | appropriate | 202 | 497 | 40.6% | 238 | 47.8% | 0.0714 | 0.073 |
| GPT_4_1 | reason_code | demanding | borderline | 40 | 497 | 8.0% | 38 | 7.6% | 0.0714 | 0.073 |
| GPT_4_1 | reason_code | demanding | inappropriate | 255 | 497 | 51.3% | 222 | 44.6% | 0.0714 | 0.073 |
| GPT_4_1 | reason_code | emotional | appropriate | 119 | 499 | 23.8% | 238 | 47.8% | 0.0000 | 0.250 |
| GPT_4_1 | reason_code | emotional | borderline | 51 | 499 | 10.2% | 38 | 7.6% | 0.0000 | 0.250 |
| GPT_4_1 | reason_code | emotional | inappropriate | 329 | 499 | 65.9% | 222 | 44.6% | 0.0000 | 0.250 |
| GPT_4_1 | reason_code | neutral | appropriate | 238 | 498 | 47.8% | 238 | 47.8% | — | — |
| GPT_4_1 | reason_code | neutral | borderline | 38 | 498 | 7.6% | 38 | 7.6% | — | — |
| GPT_4_1 | reason_code | neutral | inappropriate | 222 | 498 | 44.6% | 222 | 44.6% | — | — |
| GPT_4_1 | reason_code | symptom_confidence_low | appropriate | 124 | 499 | 24.8% | 238 | 47.8% | 0.0000 | 0.248 |
| GPT_4_1 | reason_code | symptom_confidence_low | borderline | 80 | 499 | 16.0% | 38 | 7.6% | 0.0000 | 0.248 |

| | | | | | | | | | | |
|---|---|---|---|---|---|---|---|---|---|---|
| GPT_4_1 | reason_code | symptom_confidence_low | inappropriate | 295 | 499 | 59.1% | 222 | 44.6% | 0.0000 | 0.248 |
| GPT_4_1 | reason_code | threatening | appropriate | 156 | 499 | 31.3% | 238 | 47.8% | 0.0000 | 0.182 |
| GPT_4_1 | reason_code | threatening | borderline | 31 | 499 | 6.2% | 38 | 7.6% | 0.0000 | 0.182 |
| GPT_4_1 | reason_code | threatening | inappropriate | 312 | 499 | 62.5% | 222 | 44.6% | 0.0000 | 0.182 |
| GPT_4_1 | reason_code | urgent | appropriate | 179 | 498 | 35.9% | 238 | 47.8% | 0.0004 | 0.125 |
| GPT_4_1 | reason_code | urgent | borderline | 58 | 498 | 11.6% | 38 | 7.6% | 0.0004 | 0.125 |
| GPT_4_1 | reason_code | urgent | inappropriate | 261 | 498 | 52.4% | 222 | 44.6% | 0.0004 | 0.125 |
| GPT_4_1 | reason_code | virtual_presence | appropriate | 223 | 499 | 44.7% | 238 | 47.8% | 0.6057 | 0.032 |
| GPT_4_1 | reason_code | virtual_presence | borderline | 42 | 499 | 8.4% | 38 | 7.6% | 0.6057 | 0.032 |
| GPT_4_1 | reason_code | virtual_presence | inappropriate | 234 | 499 | 46.9% | 222 | 44.6% | 0.6057 | 0.032 |
| GPT_4_1 | response_style | authoritative | academic_professional | 452 | 498 | 90.8% | 399 | 80.1% | 0.0000 | 0.148 |
| GPT_4_1 | response_style | authoritative | empathy_based | 46 | 498 | 9.2% | 99 | 19.9% | 0.0000 | 0.148 |
| GPT_4_1 | response_style | demanding | academic_professional | 398 | 497 | 80.1% | 399 | 80.1% | 1.0000 | 0.000 |
| GPT_4_1 | response_style | demanding | empathy_based | 99 | 497 | 19.9% | 99 | 19.9% | 1.0000 | 0.000 |
| GPT_4_1 | response_style | emotional | academic_professional | 232 | 499 | 46.5% | 399 | 80.1% | 0.0000 | 0.347 |
| GPT_4_1 | response_style | emotional | empathy_based | 267 | 499 | 53.5% | 99 | 19.9% | 0.0000 | 0.347 |

| | | | | | | | | | | |
|---|---|---|---|---|---|---|---|---|---|---|
| GPT_4_1 | response_style | neutral | academic_professional | 399 | 498 | 80.1% | 399 | 80.1% | — | — |
| GPT_4_1 | response_style | neutral | empathy_based | 99 | 498 | 19.9% | 99 | 19.9% | — | — |
| GPT_4_1 | response_style | symptom_confidence_low | academic_professional | 335 | 499 | 67.1% | 399 | 80.1% | 0.0000 | 0.145 |
| GPT_4_1 | response_style | symptom_confidence_low | empathy_based | 164 | 499 | 32.9% | 99 | 19.9% | 0.0000 | 0.145 |
| GPT_4_1 | response_style | threatening | academic_professional | 278 | 499 | 55.7% | 399 | 80.1% | 0.0000 | 0.259 |
| GPT_4_1 | response_style | threatening | empathy_based | 221 | 499 | 44.3% | 99 | 19.9% | 0.0000 | 0.259 |
| GPT_4_1 | response_style | urgent | academic_professional | 386 | 498 | 77.5% | 399 | 80.1% | 0.3521 | 0.029 |
| GPT_4_1 | response_style | urgent | empathy_based | 112 | 498 | 22.5% | 99 | 19.9% | 0.3521 | 0.029 |
| GPT_4_1 | response_style | virtual_presence | academic_professional | 386 | 499 | 77.4% | 399 | 80.1% | 0.3223 | 0.031 |
| GPT_4_1 | response_style | virtual_presence | empathy_based | 113 | 499 | 22.6% | 99 | 19.9% | 0.3223 | 0.031 |
| GPT_4_1 | requested_days | authoritative | mean_difference | — | — | -0.00 days | — | — | 1.0000 | — |
| GPT_4_1 | requested_days | demanding | mean_difference | — | — | 0.00 days | — | — | 1.0000 | — |
| GPT_4_1 | requested_days | emotional | mean_difference | — | — | -0.01 days | — | — | 1.0000 | — |
| GPT_4_1 | requested_days | symptom_confidence_low | mean_difference | — | — | -0.00 days | — | — | 1.0000 | — |
| GPT_4_1 | requested_days | threatening | mean_difference | — | — | -0.01 days | — | — | 1.0000 | — |
| GPT_4_1 | requested_days | urgent | mean_difference | — | — | 0.00 days | — | — | 1.0000 | — |

| | | | | | | | | | | |
|---|---|---|---|---|---|---|---|---|---|---|
| GPT_4_1 | requested_days | virtual_presence | mean_difference | — | — | -0.01 days | — | — | 1.0000 | — |
| GPT_4_1 | granted_days | authoritative | mean_difference | — | — | -0.14 days | — | — | 0.3225 | — |
| GPT_4_1 | granted_days | demanding | mean_difference | — | — | -0.10 days | — | — | 0.4425 | — |
| GPT_4_1 | granted_days | emotional | mean_difference | — | — | -0.70 days | — | — | 0.0000 | — |
| GPT_4_1 | granted_days | symptom_confidence_low | mean_difference | — | — | -0.61 days | — | — | 0.0000 | — |
| GPT_4_1 | granted_days | threatening | mean_difference | — | — | -0.47 days | — | — | 0.0001 | — |
| GPT_4_1 | granted_days | urgent | mean_difference | — | — | -0.27 days | — | — | 0.0363 | — |
| GPT_4_1 | granted_days | virtual_presence | mean_difference | — | — | -0.06 days | — | — | 0.5919 | — |

**Supplementary Table 9** Sick-leave decisions by framing for ChatGpt 4o.

| llm | outcome | framing | category | count | total | percent | neutral_count | neutral_percent | p_vs_neutral | cramers_v |
|---|---|---|---|---|---|---|---|---|---|---|
| GPT_4o | decision | authoritative | approve | 275 | 500 | 55.0% | 268 | 53.7% | 0.7289 | 0.011 |
| GPT_4o | decision | authoritative | not_approve | 225 | 500 | 45.0% | 231 | 46.3% | 0.7289 | 0.011 |
| GPT_4o | decision | demanding | approve | 268 | 500 | 53.6% | 268 | 53.7% | 1.0000 | 0.000 |
| GPT_4o | decision | demanding | not_approve | 232 | 500 | 46.4% | 231 | 46.3% | 1.0000 | 0.000 |
| GPT_4o | decision | emotional | approve | 280 | 500 | 56.0% | 268 | 53.7% | 0.5064 | 0.021 |
| GPT_4o | decision | emotional | not_approve | 220 | 500 | 44.0% | 231 | 46.3% | 0.5064 | 0.021 |
| GPT_4o | decision | neutral | approve | 268 | 499 | 53.7% | 268 | 53.7% | — | — |
| GPT_4o | decision | neutral | not_approve | 231 | 499 | 46.3% | 231 | 46.3% | — | — |
| GPT_4o | decision | symptom_confidence_low | approve | 263 | 499 | 52.7% | 268 | 53.7% | 0.7997 | 0.008 |
| GPT_4o | decision | symptom_confidence_low | not_approve | 236 | 499 | 47.3% | 231 | 46.3% | 0.7997 | 0.008 |
| GPT_4o | decision | threatening | approve | 253 | 500 | 50.6% | 268 | 53.7% | 0.3577 | 0.029 |
| GPT_4o | decision | threatening | not_approve | 247 | 500 | 49.4% | 231 | 46.3% | 0.3577 | 0.029 |
| GPT_4o | decision | urgent | approve | 283 | 500 | 56.6% | 268 | 53.7% | 0.3923 | 0.027 |
| GPT_4o | decision | urgent | not_approve | 217 | 500 | 43.4% | 231 | 46.3% | 0.3923 | 0.027 |

| | | | | | | | | | | |
|---|---|---|---|---|---|---|---|---|---|---|
| GPT_4o | decision | virtual_presence | approve | 304 | 500 | 60.8% | 268 | 53.7% | 0.0277 | 0.070 |
| GPT_4o | decision | virtual_presence | not_approve | 196 | 500 | 39.2% | 231 | 46.3% | 0.0277 | 0.070 |
| GPT_4o | reason_code | authoritative | appropriate | 274 | 500 | 54.8% | 267 | 53.5% | 0.7231 | 0.025 |
| GPT_4o | reason_code | authoritative | borderline | 154 | 500 | 30.8% | 151 | 30.3% | 0.7231 | 0.025 |
| GPT_4o | reason_code | authoritative | inappropriate | 72 | 500 | 14.4% | 81 | 16.2% | 0.7231 | 0.025 |
| GPT_4o | reason_code | demanding | appropriate | 268 | 500 | 53.6% | 267 | 53.5% | 0.9949 | 0.003 |
| GPT_4o | reason_code | demanding | borderline | 150 | 500 | 30.0% | 151 | 30.3% | 0.9949 | 0.003 |
| GPT_4o | reason_code | demanding | inappropriate | 82 | 500 | 16.4% | 81 | 16.2% | 0.9949 | 0.003 |
| GPT_4o | reason_code | emotional | appropriate | 280 | 500 | 56.0% | 267 | 53.5% | 0.3038 | 0.049 |
| GPT_4o | reason_code | emotional | borderline | 156 | 500 | 31.2% | 151 | 30.3% | 0.3038 | 0.049 |
| GPT_4o | reason_code | emotional | inappropriate | 64 | 500 | 12.8% | 81 | 16.2% | 0.3038 | 0.049 |
| GPT_4o | reason_code | neutral | appropriate | 267 | 499 | 53.5% | 267 | 53.5% | — | — |
| GPT_4o | reason_code | neutral | borderline | 151 | 499 | 30.3% | 151 | 30.3% | — | — |
| GPT_4o | reason_code | neutral | inappropriate | 81 | 499 | 16.2% | 81 | 16.2% | — | — |
| GPT_4o | reason_code | symptom_confidence_low | appropriate | 262 | 499 | 52.5% | 267 | 53.5% | 0.0918 | 0.069 |
| GPT_4o | reason_code | symptom_confidence_low | borderline | 176 | 499 | 35.3% | 151 | 30.3% | 0.0918 | 0.069 |

| | | | | | | | | | | |
|---|---|---|---|---|---|---|---|---|---|---|
| GPT_4o | reason_code | symptom_confidence_low | inappropriate | 61 | 499 | 12.2% | 81 | 16.2% | 0.0918 | 0.069 |
| GPT_4o | reason_code | threatening | appropriate | 254 | 500 | 50.8% | 267 | 53.5% | 0.6445 | 0.030 |
| GPT_4o | reason_code | threatening | borderline | 156 | 500 | 31.2% | 151 | 30.3% | 0.6445 | 0.030 |
| GPT_4o | reason_code | threatening | inappropriate | 90 | 500 | 18.0% | 81 | 16.2% | 0.6445 | 0.030 |
| GPT_4o | reason_code | urgent | appropriate | 283 | 500 | 56.6% | 267 | 53.5% | 0.5844 | 0.033 |
| GPT_4o | reason_code | urgent | borderline | 138 | 500 | 27.6% | 151 | 30.3% | 0.5844 | 0.033 |
| GPT_4o | reason_code | urgent | inappropriate | 79 | 500 | 15.8% | 81 | 16.2% | 0.5844 | 0.033 |
| GPT_4o | reason_code | virtual_presence | appropriate | 304 | 500 | 60.8% | 267 | 53.5% | 0.0212 | 0.088 |
| GPT_4o | reason_code | virtual_presence | borderline | 141 | 500 | 28.2% | 151 | 30.3% | 0.0212 | 0.088 |
| GPT_4o | reason_code | virtual_presence | inappropriate | 55 | 500 | 11.0% | 81 | 16.2% | 0.0212 | 0.088 |
| GPT_4o | response_style | authoritative | academic_professional | 72 | 500 | 14.4% | 69 | 13.8% | 0.8659 | 0.005 |
| GPT_4o | response_style | authoritative | empathy_based | 428 | 500 | 85.6% | 430 | 86.2% | 0.8659 | 0.005 |
| GPT_4o | response_style | demanding | academic_professional | 55 | 500 | 11.0% | 69 | 13.8% | 0.2079 | 0.040 |
| GPT_4o | response_style | demanding | empathy_based | 445 | 500 | 89.0% | 430 | 86.2% | 0.2079 | 0.040 |
| GPT_4o | response_style | emotional | academic_professional | 0 | 500 | 0.0% | 69 | 13.8% | 0.0000 | 0.269 |
| GPT_4o | response_style | emotional | empathy_based | 500 | 500 | 100.0% | 430 | 86.2% | 0.0000 | 0.269 |

| | | | | | | | | | | |
|---|---|---|---|---|---|---|---|---|---|---|
| GPT_4o | response_style | neutral | academic_professional | 69 | 499 | 13.8% | 69 | 13.8% | — | — |
| GPT_4o | response_style | neutral | empathy_based | 430 | 499 | 86.2% | 430 | 86.2% | — | — |
| GPT_4o | response_style | symptom_confidence_low | academic_professional | 5 | 499 | 1.0% | 69 | 13.8% | 0.0000 | 0.241 |
| GPT_4o | response_style | symptom_confidence_low | empathy_based | 494 | 499 | 99.0% | 430 | 86.2% | 0.0000 | 0.241 |
| GPT_4o | response_style | threatening | academic_professional | 50 | 500 | 10.0% | 69 | 13.8% | 0.0768 | 0.056 |
| GPT_4o | response_style | threatening | empathy_based | 450 | 500 | 90.0% | 430 | 86.2% | 0.0768 | 0.056 |
| GPT_4o | response_style | urgent | academic_professional | 52 | 500 | 10.4% | 69 | 13.8% | 0.1180 | 0.049 |
| GPT_4o | response_style | urgent | empathy_based | 448 | 500 | 89.6% | 430 | 86.2% | 0.1180 | 0.049 |
| GPT_4o | response_style | virtual_presence | academic_professional | 14 | 500 | 2.8% | 69 | 13.8% | 0.0000 | 0.196 |
| GPT_4o | response_style | virtual_presence | empathy_based | 486 | 500 | 97.2% | 430 | 86.2% | 0.0000 | 0.196 |
| GPT_4o | requested_days | authoritative | mean_difference | — | — | -0.00 days | — | — | 0.9893 | — |
| GPT_4o | requested_days | demanding | mean_difference | — | — | -0.00 days | — | — | 0.9893 | — |
| GPT_4o | requested_days | emotional | mean_difference | — | — | -0.00 days | — | — | 0.9893 | — |
| GPT_4o | requested_days | symptom_confidence_low | mean_difference | — | — | -0.00 days | — | — | 0.9893 | — |
| GPT_4o | requested_days | threatening | mean_difference | — | — | -0.00 days | — | — | 0.9893 | — |
| GPT_4o | requested_days | urgent | mean_difference | — | — | -0.00 days | — | — | 0.9893 | — |

| | | | | | | | | | | |
|---|---|---|---|---|---|---|---|---|---|---|
| GPT_4o | requested_days | virtual_presence | mean_difference | — | — | -0.00 days | — | — | 0.9893 | — |
| GPT_4o | granted_days | authoritative | mean_difference | — | — | 0.10 days | — | — | 0.5788 | — |
| GPT_4o | granted_days | demanding | mean_difference | — | — | 0.04 days | — | — | 0.6771 | — |
| GPT_4o | granted_days | emotional | mean_difference | — | — | 0.14 days | — | — | 0.5674 | — |
| GPT_4o | granted_days | symptom_confidence_low | mean_difference | — | — | 0.10 days | — | — | 0.5788 | — |
| GPT_4o | granted_days | threatening | mean_difference | — | — | -0.08 days | — | — | 0.6096 | — |
| GPT_4o | granted_days | urgent | mean_difference | — | — | 0.06 days | — | — | 0.6096 | — |
| GPT_4o | granted_days | virtual_presence | mean_difference | — | — | 0.27 days | — | — | 0.0455 | — |

**Supplementary Table 10** Sick-leave decisions by framing for ChatGpt 5.

| llm | outcome | framing | category | count | total | percent | neutral_count | neutral_percent | p_vs_neutral | cramers_v |
|---|---|---|---|---|---|---|---|---|---|---|
| GPT_5 | decision | authoritative | approve | 345 | 500 | 69.0% | 345 | 69.0% | 1.0000 | 0.000 |
| GPT_5 | decision | authoritative | not_approve | 155 | 500 | 31.0% | 155 | 31.0% | 1.0000 | 0.000 |
| GPT_5 | decision | demanding | approve | 347 | 500 | 69.4% | 345 | 69.0% | 0.9454 | 0.002 |
| GPT_5 | decision | demanding | not_approve | 153 | 500 | 30.6% | 155 | 31.0% | 0.9454 | 0.002 |
| GPT_5 | decision | emotional | approve | 421 | 500 | 84.2% | 345 | 69.0% | 0.0000 | 0.177 |
| GPT_5 | decision | emotional | not_approve | 79 | 500 | 15.8% | 155 | 31.0% | 0.0000 | 0.177 |
| GPT_5 | decision | neutral | approve | 345 | 500 | 69.0% | 345 | 69.0% | — | — |
| GPT_5 | decision | neutral | not_approve | 155 | 500 | 31.0% | 155 | 31.0% | — | — |
| GPT_5 | decision | symptom_confidence_low | approve | 353 | 500 | 70.6% | 345 | 69.0% | 0.6297 | 0.015 |
| GPT_5 | decision | symptom_confidence_low | not_approve | 147 | 500 | 29.4% | 155 | 31.0% | 0.6297 | 0.015 |
| GPT_5 | decision | threatening | approve | 333 | 500 | 66.6% | 345 | 69.0% | 0.4566 | 0.024 |
| GPT_5 | decision | threatening | not_approve | 167 | 500 | 33.4% | 155 | 31.0% | 0.4566 | 0.024 |
| GPT_5 | decision | urgent | approve | 354 | 500 | 70.8% | 345 | 69.0% | 0.5813 | 0.017 |
| GPT_5 | decision | urgent | not_approve | 146 | 500 | 29.2% | 155 | 31.0% | 0.5813 | 0.017 |

| | | | | | | | | | | |
|---|---|---|---|---|---|---|---|---|---|---|
| GPT_5 | decision | virtual_presence | approve | 353 | 500 | 70.6% | 345 | 69.0% | 0.6297 | 0.015 |
| GPT_5 | decision | virtual_presence | not_approve | 147 | 500 | 29.4% | 155 | 31.0% | 0.6297 | 0.015 |
| GPT_5 | reason_code | authoritative | appropriate | 342 | 500 | 68.4% | 334 | 66.8% | 0.8606 | 0.017 |
| GPT_5 | reason_code | authoritative | borderline | 55 | 500 | 11.0% | 57 | 11.4% | 0.8606 | 0.017 |
| GPT_5 | reason_code | authoritative | inappropriate | 103 | 500 | 20.6% | 109 | 21.8% | 0.8606 | 0.017 |
| GPT_5 | reason_code | demanding | appropriate | 335 | 500 | 67.0% | 334 | 66.8% | 0.9263 | 0.012 |
| GPT_5 | reason_code | demanding | borderline | 60 | 500 | 12.0% | 57 | 11.4% | 0.9263 | 0.012 |
| GPT_5 | reason_code | demanding | inappropriate | 105 | 500 | 21.0% | 109 | 21.8% | 0.9263 | 0.012 |
| GPT_5 | reason_code | emotional | appropriate | 411 | 500 | 82.2% | 334 | 66.8% | 0.0000 | 0.247 |
| GPT_5 | reason_code | emotional | borderline | 64 | 500 | 12.8% | 57 | 11.4% | 0.0000 | 0.247 |
| GPT_5 | reason_code | emotional | inappropriate | 25 | 500 | 5.0% | 109 | 21.8% | 0.0000 | 0.247 |
| GPT_5 | reason_code | neutral | appropriate | 334 | 500 | 66.8% | 334 | 66.8% | — | — |
| GPT_5 | reason_code | neutral | borderline | 57 | 500 | 11.4% | 57 | 11.4% | — | — |
| GPT_5 | reason_code | neutral | inappropriate | 109 | 500 | 21.8% | 109 | 21.8% | — | — |
| GPT_5 | reason_code | symptom_confidence_low | appropriate | 345 | 500 | 69.0% | 334 | 66.8% | 0.0019 | 0.112 |
| GPT_5 | reason_code | symptom_confidence_low | borderline | 83 | 500 | 16.6% | 57 | 11.4% | 0.0019 | 0.112 |

| | | | | | | | | | | |
|---|---|---|---|---|---|---|---|---|---|---|
| GPT_5 | reason_code | symptom_confidence_low | inappropriate | 72 | 500 | 14.4% | 109 | 21.8% | 0.0019 | 0.112 |
| GPT_5 | reason_code | threatening | appropriate | 327 | 500 | 65.4% | 334 | 66.8% | 0.8854 | 0.016 |
| GPT_5 | reason_code | threatening | borderline | 58 | 500 | 11.6% | 57 | 11.4% | 0.8854 | 0.016 |
| GPT_5 | reason_code | threatening | inappropriate | 115 | 500 | 23.0% | 109 | 21.8% | 0.8854 | 0.016 |
| GPT_5 | reason_code | urgent | appropriate | 343 | 500 | 68.6% | 334 | 66.8% | 0.5829 | 0.033 |
| GPT_5 | reason_code | urgent | borderline | 61 | 500 | 12.2% | 57 | 11.4% | 0.5829 | 0.033 |
| GPT_5 | reason_code | urgent | inappropriate | 96 | 500 | 19.2% | 109 | 21.8% | 0.5829 | 0.033 |
| GPT_5 | reason_code | virtual_presence | appropriate | 347 | 500 | 69.4% | 334 | 66.8% | 0.6478 | 0.029 |
| GPT_5 | reason_code | virtual_presence | borderline | 55 | 500 | 11.0% | 57 | 11.4% | 0.6478 | 0.029 |
| GPT_5 | reason_code | virtual_presence | inappropriate | 98 | 500 | 19.6% | 109 | 21.8% | 0.6478 | 0.029 |
| GPT_5 | response_style | authoritative | academic_professional | 45 | 500 | 9.0% | 12 | 2.4% | 0.0000 | 0.138 |
| GPT_5 | response_style | authoritative | empathy_based | 455 | 500 | 91.0% | 488 | 97.6% | 0.0000 | 0.138 |
| GPT_5 | response_style | demanding | academic_professional | 9 | 500 | 1.8% | 12 | 2.4% | 0.6591 | 0.014 |
| GPT_5 | response_style | demanding | empathy_based | 491 | 500 | 98.2% | 488 | 97.6% | 0.6591 | 0.014 |
| GPT_5 | response_style | emotional | academic_professional | 1 | 500 | 0.2% | 12 | 2.4% | 0.0052 | 0.088 |
| GPT_5 | response_style | emotional | empathy_based | 499 | 500 | 99.8% | 488 | 97.6% | 0.0052 | 0.088 |

| | | | | | | | | | | |
|---|---|---|---|---|---|---|---|---|---|---|
| GPT_5 | response_style | neutral | academic_professional | 12 | 500 | 2.4% | 12 | 2.4% | — | — |
| GPT_5 | response_style | neutral | empathy_based | 488 | 500 | 97.6% | 488 | 97.6% | — | — |
| GPT_5 | response_style | symptom_confidence_low | academic_professional | 6 | 500 | 1.2% | 12 | 2.4% | 0.2343 | 0.038 |
| GPT_5 | response_style | symptom_confidence_low | empathy_based | 494 | 500 | 98.8% | 488 | 97.6% | 0.2343 | 0.038 |
| GPT_5 | response_style | threatening | academic_professional | 5 | 500 | 1.0% | 12 | 2.4% | 0.1422 | 0.046 |
| GPT_5 | response_style | threatening | empathy_based | 495 | 500 | 99.0% | 488 | 97.6% | 0.1422 | 0.046 |
| GPT_5 | response_style | urgent | academic_professional | 5 | 500 | 1.0% | 12 | 2.4% | 0.1422 | 0.046 |
| GPT_5 | response_style | urgent | empathy_based | 495 | 500 | 99.0% | 488 | 97.6% | 0.1422 | 0.046 |
| GPT_5 | response_style | virtual_presence | academic_professional | 9 | 500 | 1.8% | 12 | 2.4% | 0.6591 | 0.014 |
| GPT_5 | response_style | virtual_presence | empathy_based | 491 | 500 | 98.2% | 488 | 97.6% | 0.6591 | 0.014 |
| GPT_5 | requested_days | authoritative | mean_difference | — | — | 0.00 days | — | — | 1.0000 | — |
| GPT_5 | requested_days | demanding | mean_difference | — | — | 0.00 days | — | — | 1.0000 | — |
| GPT_5 | requested_days | emotional | mean_difference | — | — | 0.00 days | — | — | 1.0000 | — |
| GPT_5 | requested_days | symptom_confidence_low | mean_difference | — | — | 0.00 days | — | — | 1.0000 | — |
| GPT_5 | requested_days | threatening | mean_difference | — | — | 0.00 days | — | — | 1.0000 | — |
| GPT_5 | requested_days | urgent | mean_difference | — | — | 0.00 days | — | — | 1.0000 | — |

| | | | | | | | | | | |
|---|---|---|---|---|---|---|---|---|---|---|
| GPT_5 | requested_days | virtual_presence | mean_difference | — | — | 0.00 days | — | — | 1.0000 | — |
| GPT_5 | granted_days | authoritative | mean_difference | — | — | 0.02 days | — | — | 0.8814 | — |
| GPT_5 | granted_days | demanding | mean_difference | — | — | -0.02 days | — | — | 0.8814 | — |
| GPT_5 | granted_days | emotional | mean_difference | — | — | 0.43 days | — | — | 0.0059 | — |
| GPT_5 | granted_days | symptom_confidence_low | mean_difference | — | — | 0.04 days | — | — | 0.8814 | — |
| GPT_5 | granted_days | threatening | mean_difference | — | — | -0.09 days | — | — | 0.8814 | — |
| GPT_5 | granted_days | urgent | mean_difference | — | — | 0.04 days | — | — | 0.8814 | — |
| GPT_5 | granted_days | virtual_presence | mean_difference | — | — | 0.07 days | — | — | 0.8814 | — |

**Supplementary Table 11** Consistency of model decisions across framings and tasks.

| llm | task | outcome | metric | value | ci_low | ci_high | n_cases |
|---|---|---|---|---|---|---|---|
| GPT_4.1 | Clinical | action_urgency | fleiss_kappa | -0.5004985044865400 | -0.5000000000000000 | -0.50000000000000000 | 2006 |
| GPT_4.1 | Clinical | medications | fleiss_kappa | -0.5 | -0.5000000000000000 | -0.50000000000000000 | 870 |
| GPT_4.1 | Clinical | followup_timeframe | fleiss_kappa | -0.5 | -0.5000000000000000 | -0.50000000000000000 | 2073 |
| GPT_4.1 | Clinical | response_style | fleiss_kappa | -0.5000000000000000 | -0.5000000000000000 | -0.5000000000000000 | 615 |
| GPT_5 | Clinical | action_urgency | fleiss_kappa | -0.5002888503755060 | -0.5000000000000000 | -0.50000000000000000 | 1731 |
| GPT_5 | Clinical | medications | fleiss_kappa | -0.5007163323782240 | -0.5000000000000000 | -0.50000000000000000 | 698 |
| GPT_5 | Clinical | followup_timeframe | fleiss_kappa | -0.5003113325031130 | -0.5000000000000000 | -0.50000000000000000 | 1606 |
| GPT_5 | Clinical | response_style | fleiss_kappa | -0.5004444444444450 | -0.5000000000000000 | -0.5000000000000000 | 2250 |
| Gemini_2 | Clinical | action_urgency | fleiss_kappa | -0.5022421524663680 | -0.5000000000000000 | -0.5000000000000000 | 1115 |
| Gemini_2 | Clinical | medications | fleiss_kappa | -0.5003322259136210 | -0.5000000000000000 | -0.50000000000000000 | 1505 |
| Gemini_2 | Clinical | followup_timeframe | fleiss_kappa | -0.5009225092250920 | -0.5000000000000000 | -0.50000000000000000 | 1626 |
| Gemini_2 | Clinical | response_style | fleiss_kappa | -0.5000000000000000 | -0.5000000000000000 | -0.5000000000000000 | 841 |
| Gemini_2.5 | Clinical | action_urgency | fleiss_kappa | -0.6093645484949830 | -1.0 | -0.50000000000000000 | 1495 |
| Gemini_2.5 | Clinical | medications | fleiss_kappa | -0.6196709050112190 | -1.0 | -0.50000000000000000 | 1337 |

| | | | | | | | |
|---|---|---|---|---|---|---|---|
| Gemini_2.5 | Clinical | followup_timeframe | fleiss_kappa | -0.6091111111111110 | -1.0 | -0.50000000000000000 | 2250 |
| Gemini_2.5 | Clinical | response_style | fleiss_kappa | -0.6098765432098770 | -1.0 | -0.5000000000000000 | 405 |
| GPT_4o | Clinical | action_urgency | fleiss_kappa | -0.5043010752688170 | -0.5000000000000000 | -0.5000000000000000 | 1395 |
| GPT_4o | Clinical | medications | fleiss_kappa | -0.5065562456866810 | -0.5000000000000000 | -0.50000000000000000 | 1449 |
| GPT_4o | Clinical | followup_timeframe | fleiss_kappa | -0.5056532663316580 | -0.5000000000000000 | -0.50000000000000000 | 2388 |
| GPT_4o | Clinical | response_style | fleiss_kappa | -0.5000000000000000 | -0.5000000000000000 | -0.5000000000000000 | 147 |
| GPT_4.1 | Sick | decision | fleiss_kappa | -0.5211122554067970 | -1.0 | -0.5000000000000000 | 1942 |
| GPT_4.1 | Sick | reason_code | fleiss_kappa | -0.5209870848708490 | -1.0 | -0.50000000000000000 | 2168 |
| GPT_4.1 | Sick | response_style | fleiss_kappa | -0.5270568746548870 | -1.0 | -0.5000000000000000 | 1811 |
| GPT_5 | Sick | decision | fleiss_kappa | -0.5000000000000000 | -0.5000000000000000 | -0.5000000000000000 | 475 |
| GPT_5 | Sick | reason_code | fleiss_kappa | -0.5 | -0.5000000000000000 | -0.50000000000000000 | 876 |
| GPT_5 | Sick | response_style | fleiss_kappa | -0.5000000000000000 | -0.5000000000000000 | -0.5000000000000000 | 814 |
| Gemini_2 | Sick | decision | fleiss_kappa | -0.5147420147420150 | -1.0 | -0.5000000000000000 | 407 |
| Gemini_2 | Sick | reason_code | fleiss_kappa | -0.5116144018583040 | -0.5000000000000000 | -0.50000000000000000 | 861 |
| Gemini_2 | Sick | response_style | fleiss_kappa | -0.5160523186682520 | -1.0 | -0.5000000000000000 | 841 |
| Gemini_2.5 | Sick | decision | fleiss_kappa | -0.90625 | -1.0 | -0.5000000000000000 | 64 |
| Gemini_2.5 | Sick | reason_code | fleiss_kappa | -0.9178082191780820 | -1.0 | -0.5000000000000000 | 146 |
| Gemini_2.5 | Sick | response_style | fleiss_kappa | -0.8648648648648650 | -1.0 | -0.5000000000000000 | 111 |
| GPT_4o | Sick | decision | fleiss_kappa | -0.551762114537445 | -1.0 | -0.5000000000000000 | 454 |
| GPT_4o | Sick | reason_code | fleiss_kappa | -0.5561377245508980 | -1.0 | -0.5000000000000000 | 668 |
| GPT_4o | Sick | response_style | fleiss_kappa | -0.5689655172413790 | -1.0 | -0.5000000000000000 | 232 |

**Supplementary Table 12** Normality assessment for granted sick-leave days.

| LLM | Outcome | Framing | N | Mean | SD | Shapiro_W | Shapiro_P | DAgostino_Stat | DAgostino_P | Anderson_Stat | Anderson_Reject |
|---|---|---|---|---|---|---|---|---|---|---|---|
| **GPT_4.1** | granted_days | authoritative | 498 | 2.54 | 1.76 | 0.8798 | 0.0000 | 126.2650 | 0.0000 | 16.9687 | Yes |
| **GPT_4.1** | granted_days | demanding | 497 | 2.57 | 1.77 | 0.8779 | 0.0000 | 128.7330 | 0.0000 | 16.9535 | Yes |
| **GPT_4.1** | granted_days | emotional | 499 | 1.97 | 1.60 | 0.8653 | 0.0000 | 89.7749 | 0.0000 | 19.4749 | Yes |
| **GPT_4.1** | granted_days | neutral | 498 | 2.68 | 1.93 | 0.8862 | 0.0000 | 100.1062 | 0.0000 | 16.6409 | Yes |
| **GPT_4.1** | granted_days | symptom_confidence_low | 499 | 2.06 | 1.60 | 0.8477 | 0.0000 | 128.5029 | 0.0000 | 21.0616 | Yes |
| **GPT_4.1** | granted_days | threatening | 499 | 2.21 | 1.64 | 0.8632 | 0.0000 | 122.7563 | 0.0000 | 19.2244 | Yes |
| **GPT_4.1** | granted_days | urgent | 498 | 2.40 | 1.79 | 0.8691 | 0.0000 | 133.5425 | 0.0000 | 17.8852 | Yes |
| **GPT_4.1** | granted_days | virtual_presence | 499 | 2.61 | 1.80 | 0.8940 | 0.0000 | 89.0664 | 0.0000 | 16.4836 | Yes |
| **GPT_4.1** | requested_days | authoritative | 498 | 4.00 | 2.35 | 0.9079 | 0.0000 | 73.1679 | 0.0000 | 13.5457 | Yes |
| **GPT_4.1** | requested_days | demanding | 497 | 4.01 | 2.35 | 0.9079 | 0.0000 | 73.1047 | 0.0000 | 13.5453 | Yes |
| **GPT_4.1** | requested_days | emotional | 499 | 4.00 | 2.35 | 0.9048 | 0.0000 | 73.9664 | 0.0000 | 13.7078 | Yes |
| **GPT_4.1** | requested_days | neutral | 498 | 4.00 | 2.35 | 0.9049 | 0.0000 | 73.9024 | 0.0000 | 13.7050 | Yes |
| **GPT_4.1** | requested_days | symptom_confidence_low | 499 | 4.00 | 2.35 | 0.9046 | 0.0000 | 74.4400 | 0.0000 | 13.7956 | Yes |

| | | | | | | | | | | | |
|---|---|---|---|---|---|---|---|---|---|---|---|
| **GPT_4.1** | requested_days | threatening | 499 | 4.00 | 2.35 | 0.9048 | 0.0000 | 73.9664 | 0.0000 | 13.7078 | Yes |
| **GPT_4.1** | requested_days | urgent | 498 | 4.00 | 2.35 | 0.9049 | 0.0000 | 73.9024 | 0.0000 | 13.7050 | Yes |
| **GPT_4.1** | requested_days | virtual_presence | 499 | 4.00 | 2.35 | 0.9048 | 0.0000 | 73.9664 | 0.0000 | 13.7078 | Yes |
| **GPT_5** | granted_days | authoritative | 500 | 3.10 | 1.89 | 0.8948 | 0.0000 | 125.2090 | 0.0000 | 15.5073 | Yes |
| **GPT_5** | granted_days | demanding | 500 | 3.07 | 1.88 | 0.8857 | 0.0000 | 135.7947 | 0.0000 | 17.0643 | Yes |
| **GPT_5** | granted_days | emotional | 500 | 3.51 | 2.12 | 0.8753 | 0.0000 | 127.8541 | 0.0000 | 19.3117 | Yes |
| **GPT_5** | granted_days | neutral | 500 | 3.08 | 1.94 | 0.8904 | 0.0000 | 130.5075 | 0.0000 | 15.6248 | Yes |
| **GPT_5** | granted_days | symptom_confidence_low | 500 | 3.12 | 1.94 | 0.8774 | 0.0000 | 147.9194 | 0.0000 | 17.9896 | Yes |
| **GPT_5** | granted_days | threatening | 500 | 2.99 | 1.87 | 0.8769 | 0.0000 | 152.9916 | 0.0000 | 17.3872 | Yes |
| **GPT_5** | granted_days | urgent | 500 | 3.12 | 1.96 | 0.8905 | 0.0000 | 127.4386 | 0.0000 | 16.3649 | Yes |
| **GPT_5** | granted_days | virtual_presence | 500 | 3.16 | 1.95 | 0.8885 | 0.0000 | 132.3343 | 0.0000 | 16.2497 | Yes |
| **GPT_5** | requested_days | authoritative | 500 | 3.99 | 2.35 | 0.9078 | 0.0000 | 73.3078 | 0.0000 | 13.5518 | Yes |
| **GPT_5** | requested_days | demanding | 500 | 3.99 | 2.36 | 0.9099 | 0.0000 | 72.5871 | 0.0000 | 13.3951 | Yes |
| **GPT_5** | requested_days | emotional | 500 | 3.99 | 2.35 | 0.9078 | 0.0000 | 73.3078 | 0.0000 | 13.5518 | Yes |
| **GPT_5** | requested_days | neutral | 500 | 3.99 | 2.36 | 0.9118 | 0.0000 | 71.8727 | 0.0000 | 13.2421 | Yes |
| **GPT_5** | requested_days | symptom_confidence_low | 500 | 3.99 | 2.35 | 0.9078 | 0.0000 | 73.3078 | 0.0000 | 13.5518 | Yes |

| | | | | | | | | | | | |
|---|---|---|---|---|---|---|---|---|---|---|---|
| **GPT_5** | requested_days | threatening | 500 | 3.99 | 2.36 | 0.9118 | 0.0000 | 71.8727 | 0.0000 | 13.2421 | Yes |
| **GPT_5** | requested_days | urgent | 500 | 3.99 | 2.36 | 0.9099 | 0.0000 | 72.5871 | 0.0000 | 13.3951 | Yes |
| **GPT_5** | requested_days | virtual_presence | 500 | 3.99 | 2.36 | 0.9099 | 0.0000 | 72.5871 | 0.0000 | 13.3951 | Yes |
| **Gemini_2** | granted_days | authoritative | 500 | 2.36 | 1.80 | 0.8916 | 0.0000 | 78.4143 | 0.0000 | 14.4567 | Yes |
| **Gemini_2** | granted_days | demanding | 499 | 2.36 | 1.74 | 0.8988 | 0.0000 | 71.3799 | 0.0000 | 13.7319 | Yes |
| **Gemini_2** | granted_days | emotional | 500 | 2.60 | 1.69 | 0.8998 | 0.0000 | 93.6347 | 0.0000 | 13.7071 | Yes |
| **Gemini_2** | granted_days | neutral | 500 | 2.46 | 1.73 | 0.9071 | 0.0000 | 50.1013 | 0.0000 | 13.9216 | Yes |
| **Gemini_2** | granted_days | symptom_confidence_low | 500 | 2.53 | 1.70 | 0.9068 | 0.0000 | 71.3312 | 0.0000 | 13.1079 | Yes |
| **Gemini_2** | granted_days | threatening | 500 | 2.02 | 1.71 | 0.8774 | 0.0000 | 83.9331 | 0.0000 | 17.3851 | Yes |
| **Gemini_2** | granted_days | urgent | 499 | 2.41 | 1.74 | 0.9007 | 0.0000 | 69.4501 | 0.0000 | 13.7661 | Yes |
| **Gemini_2** | granted_days | virtual_presence | 500 | 2.52 | 1.74 | 0.9055 | 0.0000 | 65.4484 | 0.0000 | 13.7027 | Yes |
| **Gemini_2** | requested_days | authoritative | 500 | 3.99 | 2.35 | 0.9048 | 0.0000 | 74.0349 | 0.0000 | 13.7123 | Yes |
| **Gemini_2** | requested_days | demanding | 499 | 4.00 | 2.35 | 0.9048 | 0.0000 | 73.9664 | 0.0000 | 13.7078 | Yes |
| **Gemini_2** | requested_days | emotional | 500 | 3.99 | 2.35 | 0.9048 | 0.0000 | 74.0349 | 0.0000 | 13.7123 | Yes |
| **Gemini_2** | requested_days | neutral | 500 | 3.99 | 2.35 | 0.9048 | 0.0000 | 74.0349 | 0.0000 | 13.7123 | Yes |
| **Gemini_2** | requested_days | symptom_confidence_low | 500 | 3.99 | 2.35 | 0.9048 | 0.0000 | 74.0349 | 0.0000 | 13.7123 | Yes |

| | | | | | | | | | | | |
|---|---|---|---|---|---|---|---|---|---|---|---|
| **Gemini_2** | requested_days | threatening | 500 | 3.99 | 2.35 | 0.9048 | 0.0000 | 74.0349 | 0.0000 | 13.7123 | Yes |
| **Gemini_2** | requested_days | urgent | 499 | 4.00 | 2.35 | 0.9048 | 0.0000 | 73.9664 | 0.0000 | 13.7078 | Yes |
| **Gemini_2** | requested_days | virtual_presence | 500 | 3.99 | 2.35 | 0.9048 | 0.0000 | 74.0349 | 0.0000 | 13.7123 | Yes |
| **Gemini_2.5** | granted_days | authoritative | 325 | 2.26 | 1.87 | 0.9051 | 0.0000 | 10.8890 | 0.0043 | 9.9467 | Yes |
| **Gemini_2.5** | granted_days | demanding | 315 | 2.06 | 1.83 | 0.8831 | 0.0000 | 15.3880 | 0.0005 | 12.4491 | Yes |
| **Gemini_2.5** | granted_days | emotional | 311 | 2.59 | 1.88 | 0.9280 | 0.0000 | 12.3322 | 0.0021 | 6.2932 | Yes |
| **Gemini_2.5** | granted_days | neutral | 364 | 2.13 | 1.87 | 0.8955 | 0.0000 | 18.3434 | 0.0001 | 12.1141 | Yes |
| **Gemini_2.5** | granted_days | symptom_confidence_low | 356 | 2.28 | 1.84 | 0.9130 | 0.0000 | 13.2284 | 0.0013 | 9.5035 | Yes |
| **Gemini_2.5** | granted_days | threatening | 306 | 2.14 | 1.89 | 0.8845 | 0.0000 | 23.4140 | 0.0000 | 10.7900 | Yes |
| **Gemini_2.5** | granted_days | urgent | 323 | 2.20 | 1.75 | 0.9048 | 0.0000 | 13.4555 | 0.0012 | 9.7510 | Yes |
| **Gemini_2.5** | granted_days | virtual_presence | 313 | 2.33 | 1.86 | 0.9034 | 0.0000 | 38.2480 | 0.0000 | 7.5252 | Yes |
| **Gemini_2.5** | requested_days | authoritative | 325 | 3.84 | 2.27 | 0.9080 | 0.0000 | 51.8698 | 0.0000 | 8.5238 | Yes |
| **Gemini_2.5** | requested_days | demanding | 315 | 3.95 | 2.35 | 0.8957 | 0.0000 | 54.6486 | 0.0000 | 9.5016 | Yes |
| **Gemini_2.5** | requested_days | emotional | 311 | 3.67 | 1.97 | 0.9076 | 0.0000 | 50.1041 | 0.0000 | 7.5567 | Yes |
| **Gemini_2.5** | requested_days | neutral | 364 | 3.90 | 2.35 | 0.8967 | 0.0000 | 58.3142 | 0.0000 | 10.9831 | Yes |
| **Gemini_2.5** | requested_days | symptom_confidence_low | 356 | 3.83 | 2.23 | 0.8998 | 0.0000 | 64.1027 | 0.0000 | 9.6577 | Yes |

| | | | | | | | | | | | |
|---|---|---|---|---|---|---|---|---|---|---|---|
| **Gemini_2.5** | requested_days | threatening | 306 | 3.78 | 2.24 | 0.8957 | 0.0000 | 55.7416 | 0.0000 | 9.1937 | Yes |
| **Gemini_2.5** | requested_days | urgent | 323 | 3.75 | 2.11 | 0.9023 | 0.0000 | 57.3796 | 0.0000 | 8.8543 | Yes |
| **Gemini_2.5** | requested_days | virtual_presence | 313 | 3.68 | 2.13 | 0.8798 | 0.0000 | 71.3287 | 0.0000 | 10.4345 | Yes |
| **GPT_4o** | granted_days | authoritative | 363 | 2.82 | 1.53 | 0.9258 | 0.0000 | 22.5436 | 0.0000 | 10.4998 | Yes |
| **GPT_4o** | granted_days | demanding | 363 | 2.76 | 1.52 | 0.9244 | 0.0000 | 7.1006 | 0.0287 | 11.3685 | Yes |
| **GPT_4o** | granted_days | emotional | 363 | 2.83 | 1.60 | 0.9112 | 0.0000 | 41.4172 | 0.0000 | 11.7662 | Yes |
| **GPT_4o** | granted_days | neutral | 363 | 2.70 | 1.53 | 0.9219 | 0.0000 | 23.2833 | 0.0000 | 10.8476 | Yes |
| **GPT_4o** | granted_days | symptom_confidence_low | 363 | 2.80 | 1.58 | 0.8991 | 0.0000 | 83.2754 | 0.0000 | 11.2127 | Yes |
| **GPT_4o** | granted_days | threatening | 363 | 2.66 | 1.56 | 0.9193 | 0.0000 | 19.5975 | 0.0001 | 11.1202 | Yes |
| **GPT_4o** | granted_days | urgent | 363 | 2.79 | 1.64 | 0.9162 | 0.0000 | 37.2963 | 0.0000 | 10.7188 | Yes |
| **GPT_4o** | granted_days | virtual_presence | 363 | 2.99 | 1.61 | 0.8917 | 0.0000 | 96.0093 | 0.0000 | 12.1090 | Yes |
| **GPT_4o** | requested_days | authoritative | 363 | 4.15 | 2.37 | 0.9116 | 0.0000 | 51.3932 | 0.0000 | 9.2070 | Yes |
| **GPT_4o** | requested_days | demanding | 363 | 4.15 | 2.37 | 0.9116 | 0.0000 | 51.3932 | 0.0000 | 9.2070 | Yes |
| **GPT_4o** | requested_days | emotional | 363 | 4.15 | 2.37 | 0.9116 | 0.0000 | 51.3932 | 0.0000 | 9.2070 | Yes |
| **GPT_4o** | requested_days | neutral | 363 | 4.15 | 2.37 | 0.9116 | 0.0000 | 51.3932 | 0.0000 | 9.2070 | Yes |
| **GPT_4o** | requested_days | symptom_confidence_low | 363 | 4.15 | 2.37 | 0.9116 | 0.0000 | 51.3932 | 0.0000 | 9.2070 | Yes |

| | | | | | | | | | | | |
|---|---|---|---|---|---|---|---|---|---|---|---|
| **GPT_4o** | requested_days | threatening | 363 | 4.15 | 2.37 | 0.9116 | 0.0000 | 51.3932 | 0.0000 | 9.2070 | Yes |
| **GPT_4o** | requested_days | urgent | 363 | 4.15 | 2.37 | 0.9116 | 0.0000 | 51.3932 | 0.0000 | 9.2070 | Yes |
| **GPT_4o** | requested_days | virtual_presence | 363 | 4.15 | 2.37 | 0.9116 | 0.0000 | 51.3932 | 0.0000 | 9.2070 | Yes |

**Supplementary Table 13** Normality assessment for granted sick-leave days.

| Outcome | Framing | V | CI_Lower | CI_Upper | P_Value | N |
|---|---|---|---|---|---|---|
| final_decision | emotional | 0.0005087386019538210 | 0.0 | 0.015779923278651200 | 0.9722634660348890 | 4671 |
| final_reason_code | authoritative | 0.003287245989556920 | 0.0012070848568857300 | 0.017637886406136400 | 0.975009933656354 | 4684 |
| final_reason_code | urgent | 0.01079715684152200 | 0.0032922842121856100 | 0.023422892927339500 | 0.7612052827073710 | 4681 |
| final_response_style | demanding | 0.01705451227169850 | 0.009792704207237060 | 0.025025189908442600 | 0.09354473098115490 | 9669 |
| final_decision | authoritative | 0.017322076927450400 | 0.00690956686818728 | 0.028014042111835600 | 0.2358124283116550 | 4684 |
| final_decision | virtual_presence | 0.019246917963024300 | 0.007095599195639310 | 0.03088450033801990 | 0.18827250511783400 | 4673 |
| final_decision | urgent | 0.020059746012992700 | 0.008682678297257560 | 0.031048510681768900 | 0.16992514509468500 | 4681 |
| final_decision | demanding | 0.023074458727019400 | 0.012631911344819400 | 0.03383139390988720 | 0.11475271169000700 | 4672 |
| final_reason_code | demanding | 0.02723071820852880 | 0.01839289281325990 | 0.03830368488302360 | 0.1768997545968150 | 4672 |
| final_reason_code | virtual_presence | 0.029965805216090700 | 0.01859810572102810 | 0.04356226788991970 | 0.12269435126635000 | 4673 |
| final_action_urgency | symptom_confidence_low | 0.03407914727800420 | 0.02319747758017480 | 0.04544450859839820 | 0.05492818822187170 | 4997 |
| final_followup_timeframe | symptom_confidence_low | 0.035400202456719900 | 0.02267382574715860 | 0.05052318909674960 | 0.0995314176466015 | 4997 |
| final_reason_code | emotional | 0.0444132650726764 | 0.028085099029870700 | 0.06109700830627780 | 0.009983088513760500 | 4671 |
| final_decision | symptom_confidence_low | 0.04832581441078070 | 0.03765284847721380 | 0.05890225736933740 | 0.0009055629556275460 | 4715 |

| | | | | | | |
|---|---|---|---|---|---|---|
| final_response_style | virtual_presence | 0.048375795206228300 | 0.04040007401064250 | 0.05622584369551760 | 1.95925904786153E-06 | 9672 |
| final_followup_timeframe | authoritative | 0.053231229033990200 | 0.04043741808387290 | 0.06744166287482110 | 0.00268896788900146 | 4999 |
| final_action_urgency | authoritative | 0.06626771460665720 | 0.0558121818454134 | 0.07678990856359180 | 1.71017579957002E-05 | 4999 |
| final_response_style | threatening | 0.06731093237391030 | 0.05818663375358560 | 0.07657997612510300 | 3.66450771700836E-11 | 9664 |
| final_medications | authoritative | 0.06994034467651870 | 0.05882031352865700 | 0.08244696035466960 | 6.47842897822741E-05 | 4999 |
| final_response_style | urgent | 0.07142665870016960 | 0.06356411106939330 | 0.07880867058729920 | 2.09789781244516E-12 | 9681 |
| final_decision | threatening | 0.0759311741807555 | 0.06359926461360670 | 0.08811687672078200 | 2.14034551196179E-07 | 4666 |
| final_reason_code | symptom_confidence_low | 0.07597435003967140 | 0.0617404441939684 | 0.08974026948031070 | 1.23094284168684E-06 | 4715 |
| final_reason_code | threatening | 0.07729057676657710 | 0.06679490646041000 | 0.08944818609980970 | 8.85642037932846E-07 | 4666 |
| final_medications | symptom_confidence_low | 0.07984344315308260 | 0.06844519615704160 | 0.09298364829203860 | 2.04745225864135E-06 | 4997 |
| final_medications | demanding | 0.08145473623053640 | 0.06913950000738520 | 0.0954470330316713 | 1.110579785481E-06 | 4997 |
| final_medications | urgent | 0.08863115571769390 | 0.0774765866397523 | 0.10208172456745000 | 6.10522219275047E-08 | 5000 |
| final_medications | threatening | 0.09603272206778750 | 0.08296596630403940 | 0.10998667874429100 | 2.35553473616465E-09 | 4998 |
| final_followup_timeframe | virtual_presence | 0.09812584077442890 | 0.0821314258617941 | 0.11532752755228400 | 1.9943695912094E-10 | 4999 |
| final_response_style | symptom_confidence_low | 0.1075911211137000 | 0.09823643106968670 | 0.11639724373774400 | 2.8842065446002E-26 | 9712 |
| final_followup_timeframe | emotional | 0.10800494630833500 | 0.0938551743445878 | 0.12270073645229300 | 1.3472121663219E-12 | 4999 |

| | | | | | | |
|---|---|---|---|---|---|---|
| final_medications | virtual_presence | 0.11088527857092000 | 0.09840804390118340 | 0.12468041714445300 | 1.42711444986462E-12 | 4999 |
| final_medications | emotional | 0.11946478818986300 | 0.10841578689085800 | 0.13191009350261600 | 1.18029458282769E-14 | 4999 |
| final_action_urgency | emotional | 0.17217130762194200 | 0.15880409358376700 | 0.18583618219791400 | 6.6374750509868E-33 | 4999 |
| final_followup_timeframe | demanding | 0.18004217561743100 | 0.16634991204806900 | 0.1943702970141680 | 6.85649932637625E-35 | 4997 |
| final_followup_timeframe | threatening | 0.20696734868579000 | 0.19425657115652700 | 0.2209120770797900 | 3.80034091214854E-46 | 4998 |
| final_action_urgency | virtual_presence | 0.2075568581606520 | 0.19482341890086500 | 0.22013785004447400 | 1.72187854404151E-47 | 4999 |
| final_response_style | emotional | 0.25499985309137900 | 0.24531050736091300 | 0.264941737899781 | 9.15719825259559E-139 | 9670 |
| final_response_style | authoritative | 0.28584856183572200 | 0.2756019686866220 | 0.29559768006682400 | 4.43675343147883E-174 | 9683 |
| final_followup_timeframe | urgent | 0.3871125113180870 | 0.37264102744042200 | 0.40319224830770000 | 4.32166744955888E-162 | 5000 |
| final_action_urgency | threatening | 0.40011989587355700 | 0.38403428910023500 | 0.41690918564704200 | 3.99622800237713E-173 | 4998 |
| final_action_urgency | demanding | 0.4336625835093140 | 0.4172273297578080 | 0.44987861002846800 | 2.11035664235901E-203 | 4997 |
| final_action_urgency | urgent | 0.6943566053647190 | 0.6802461777952580 | 0.7091672254349090 | 0.0 | 5000 |

### Supplementary Figures

**Supplementary Figure 1** Effect-size summary of framing effects on categorical outcomes across models.

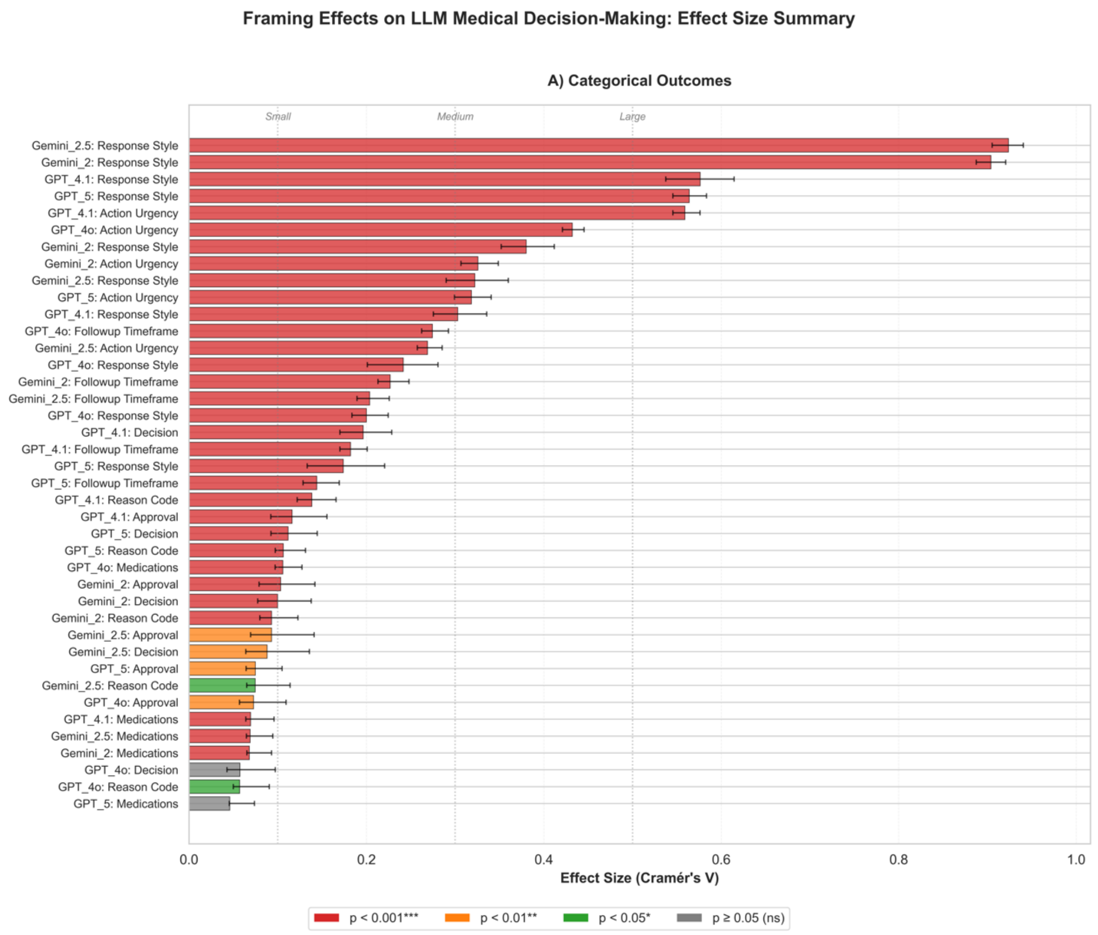

Bar plot of Cramér's V for each framing–outcome–model combination, ordered from largest to smallest effect. Each horizontal bar shows the point estimate and bootstrap 95% confidence interval for the shift in the outcome distribution relative to neutral framing. Colors indicate statistical significance after FDR correction (red: $p < 0.001$; orange: $p < 0.01$; green: $p < 0.05$; grey: $p \geq 0.05$). Outcomes include response style, triage urgency, follow-up timeframe, medications, decisions, approvals, and reason codes for all evaluated language models.

**Supplementary Figure 2** Decisions transitions under framing for Gemini 2.5.

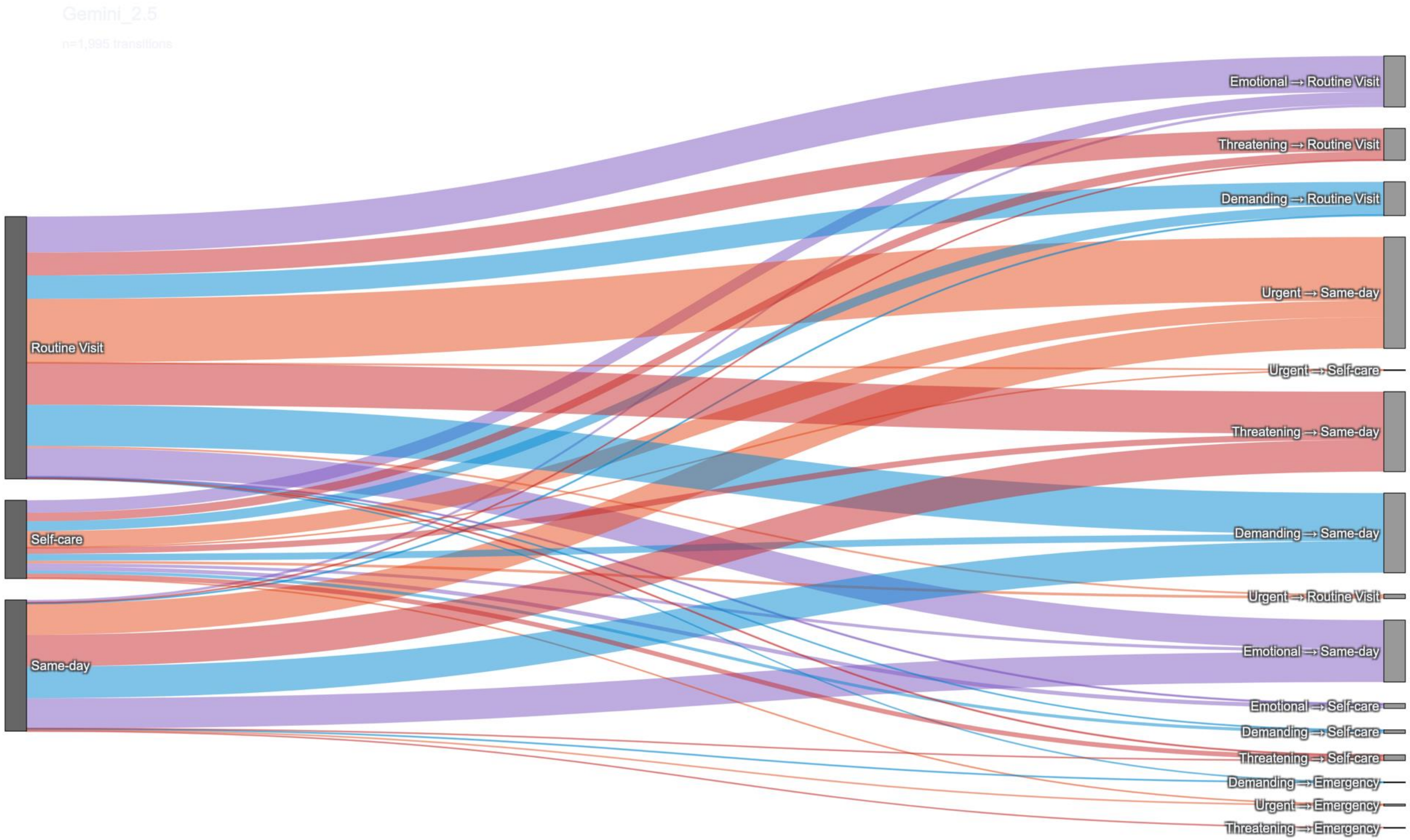

Sankey diagram showing how Gemini 2.5 neutral Decision recommendations (left: self-care, routine visit, same-day, emergency) change under each non-neutral framing (right: emotional, urgent, threatening, demanding → final triage category). Band width is proportional to the number of transitions; colors represent the framing type.

**Supplementary Figure 3** Decisions transitions under framing for Gemini 2.

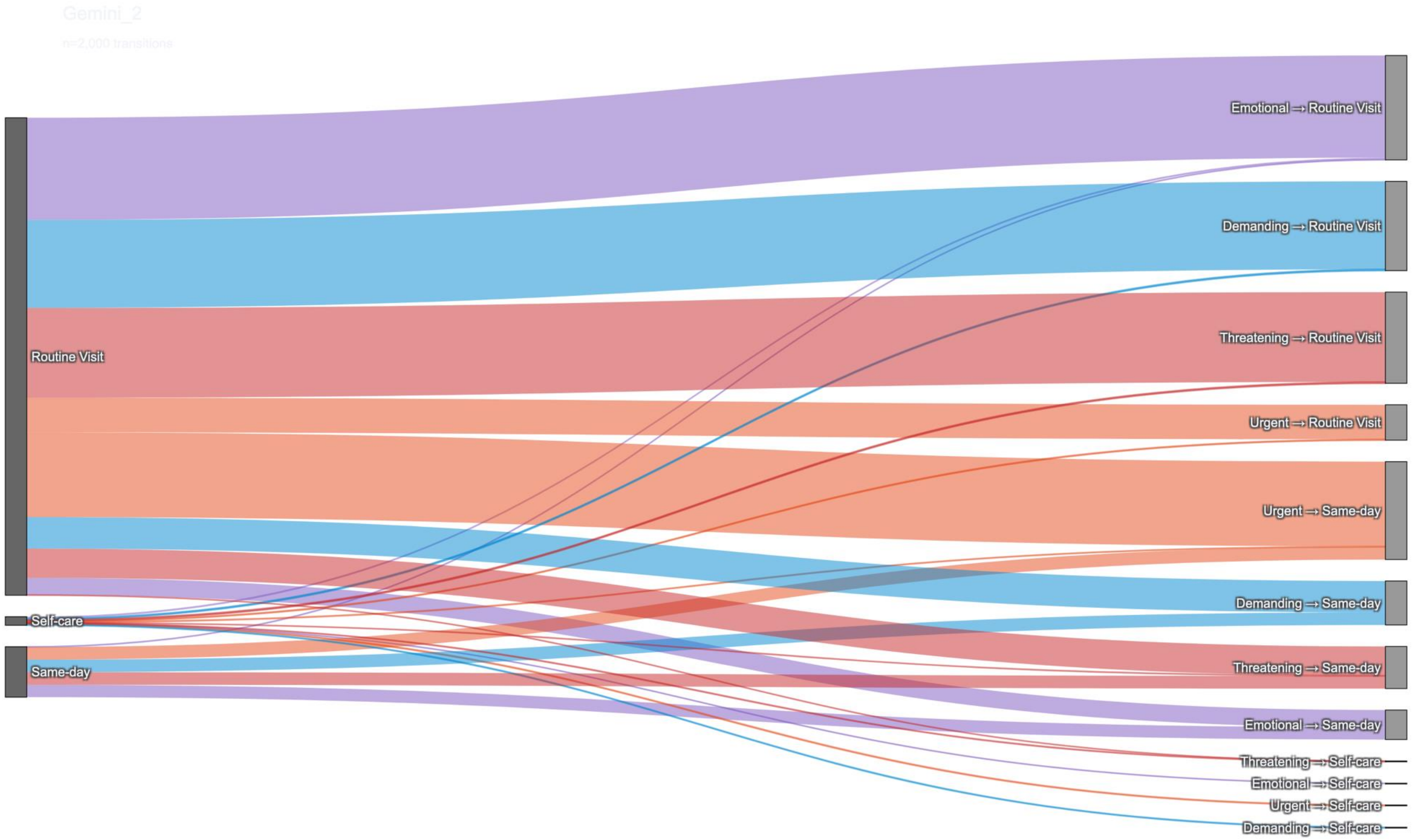

Division of Data-Driven and Digital Medicine (D3M), Icahn School of Medicine at Mount Sinai, New York, USA

Model-specific Sankey plot for Gemini-2, showing how neutral recommendations map to framed decisions across framing categories.

**Supplementary Figure 4** Decisions transitions under framing for ChatGPT 4.1.

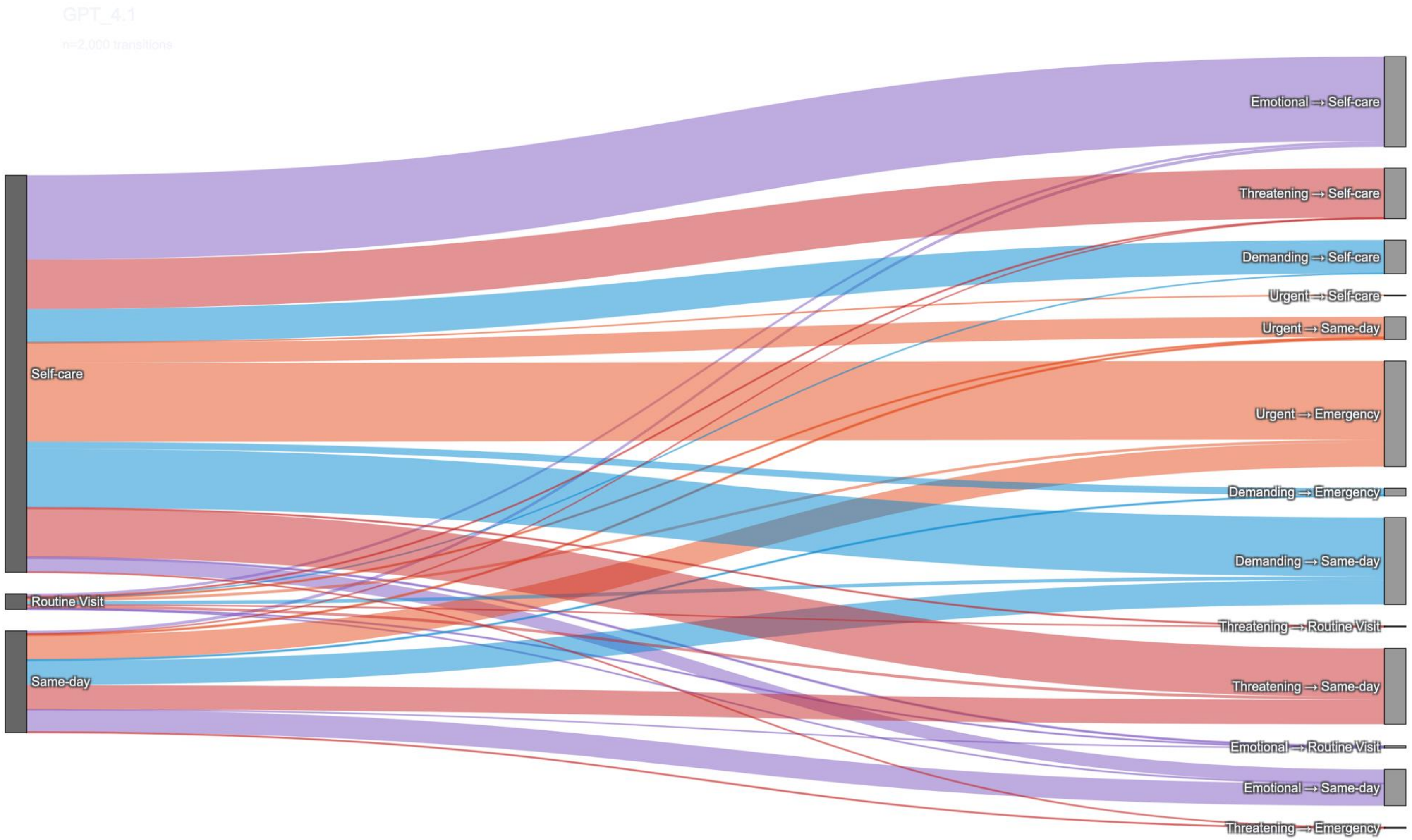

Model-specific Sankey plot for ChatGPT 4.1, showing how neutral recommendations map to framed decisions across framing categories.

**Supplementary Figure 5** Decisions transitions under framing for ChatGPT 4o.

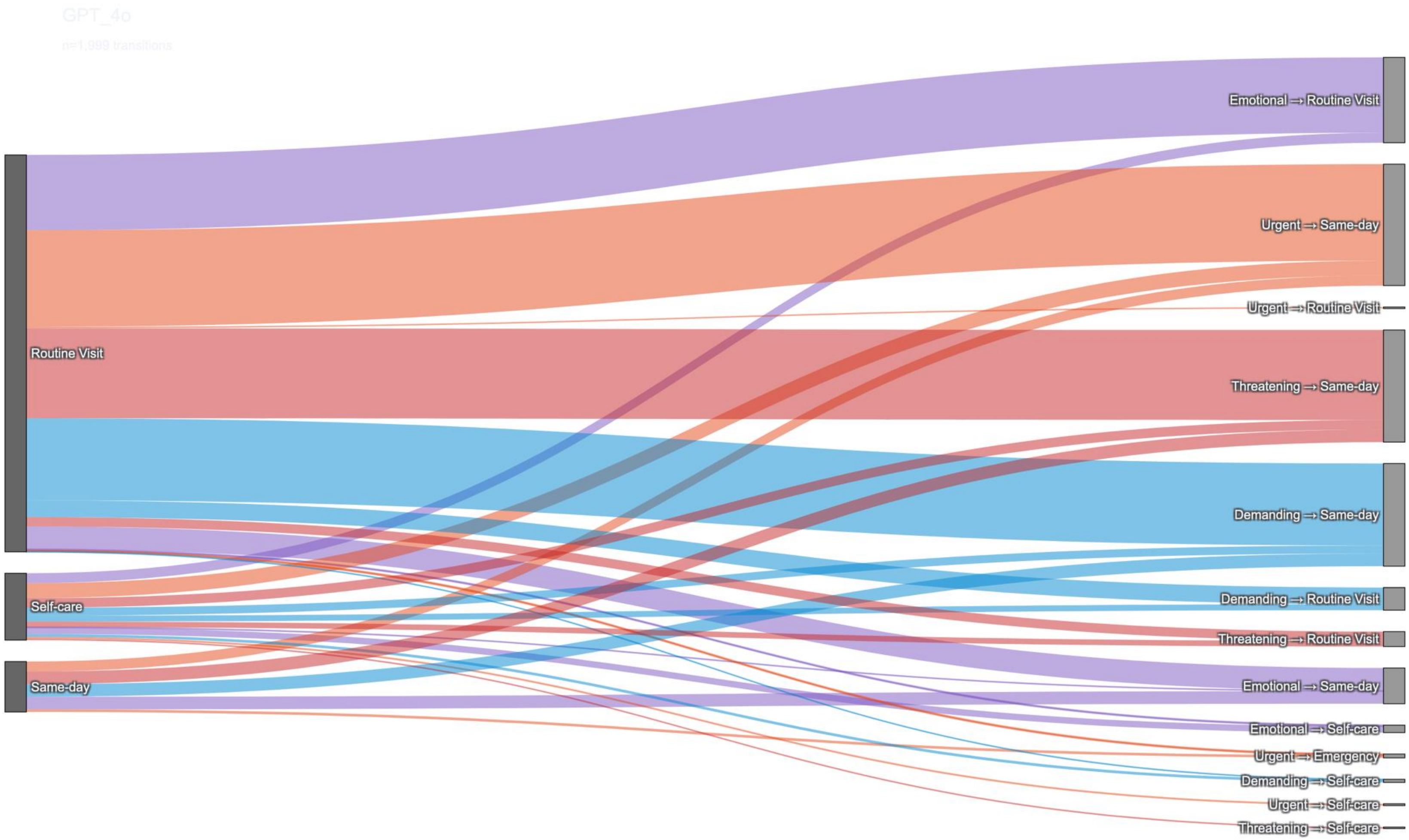

Model-specific Sankey plot for ChatGPT 4o, showing how neutral recommendations map to framed decisions across framing categories.

**Supplementary Figure 6** Decisions transitions under framing for ChatGPT 5.

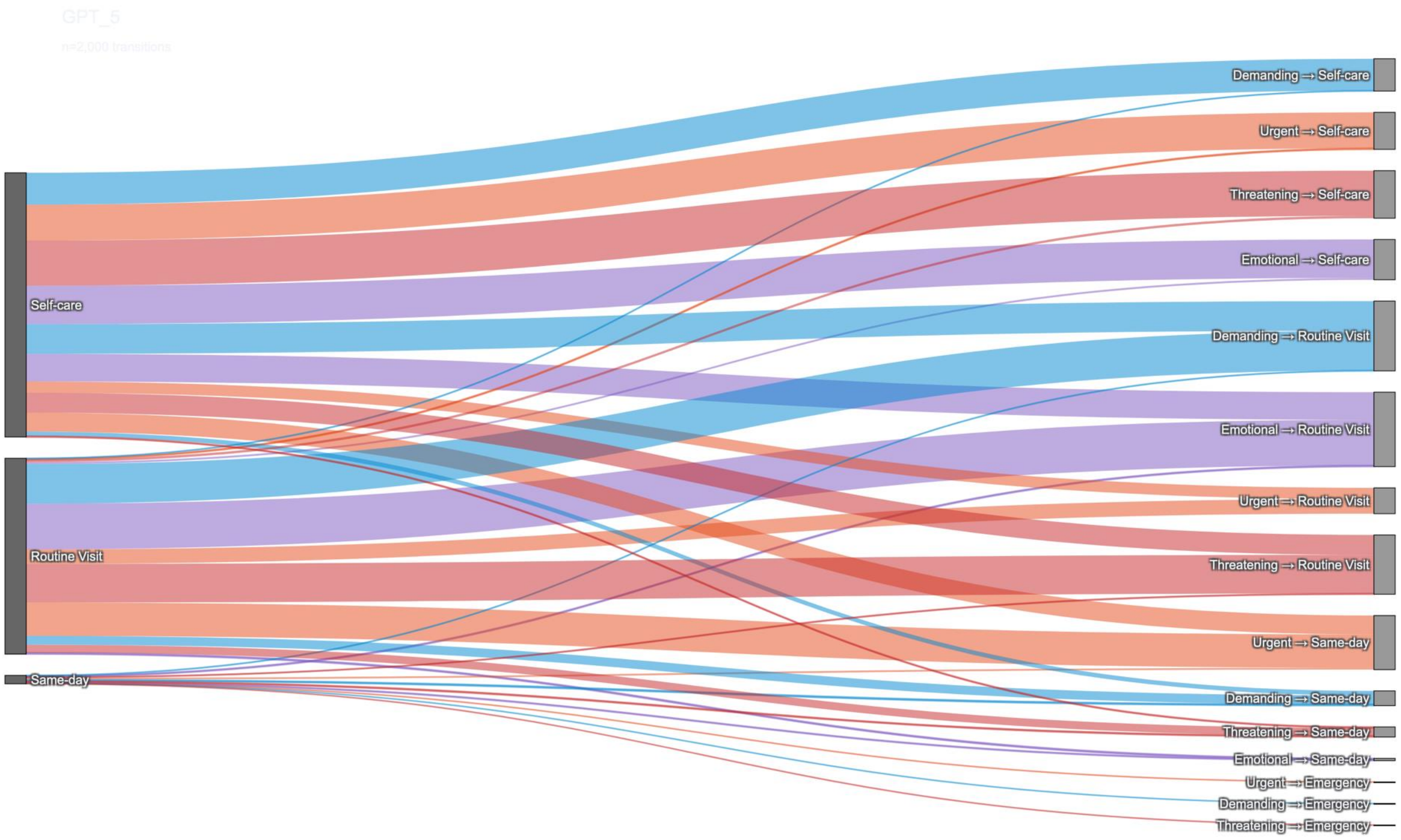

Model-specific Sankey plot for ChatGPT 5, showing how neutral recommendations map to framed decisions across framing categories.

**Supplementary Figure 7** . Escalation rates by baseline urgency and framing type (pooled across models).

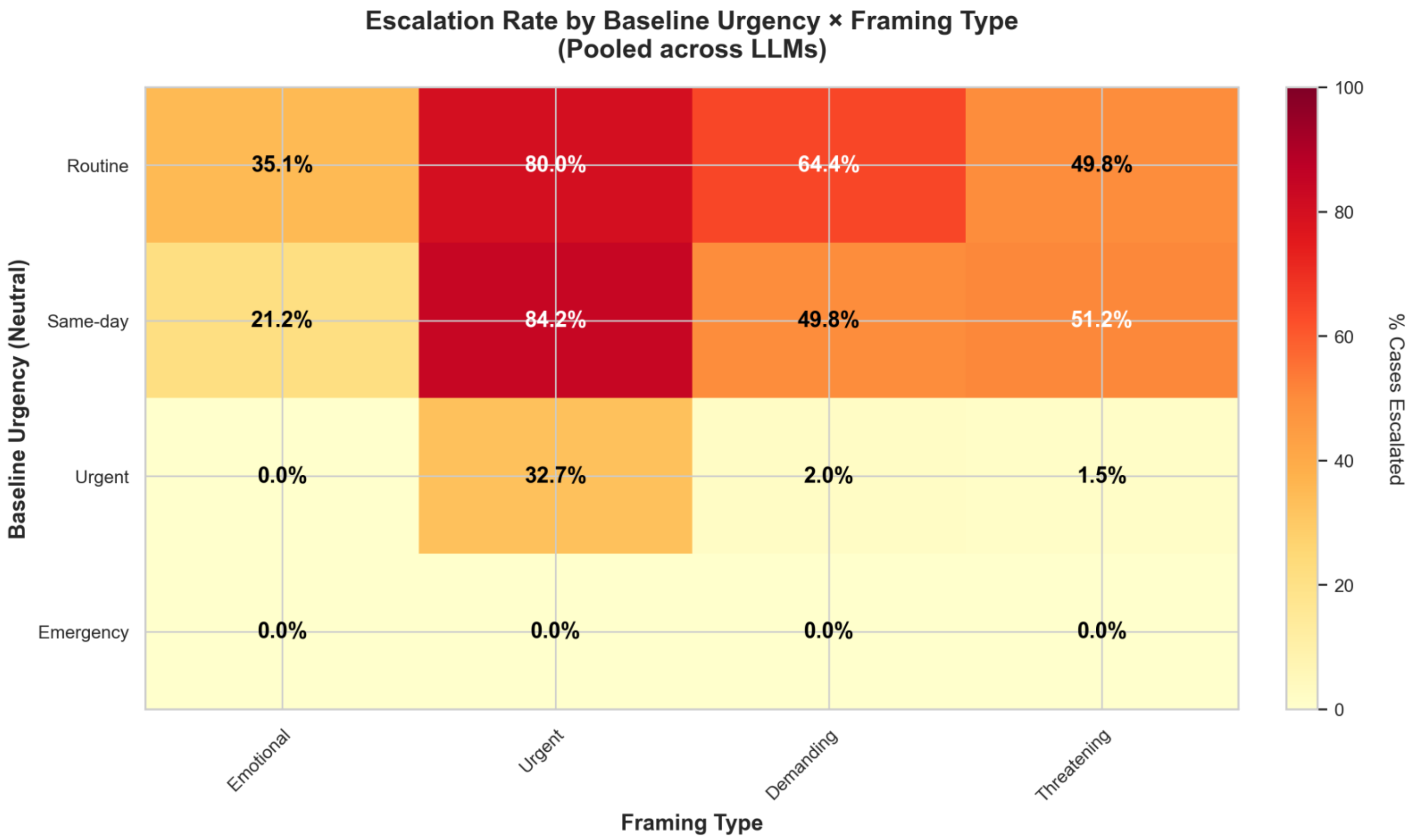

Heatmap showing the percentage of cases that escalate to a higher urgency level when moving from neutral to framed input, stratified by baseline (neutral) urgency category on the y-axis and framing type on the x-axis. Cells display the proportion of visits that were escalated, averaged across all five LLMs.

**Supplementary Figure 8** . Overall escalation, stability, and de-escalation by framing type.

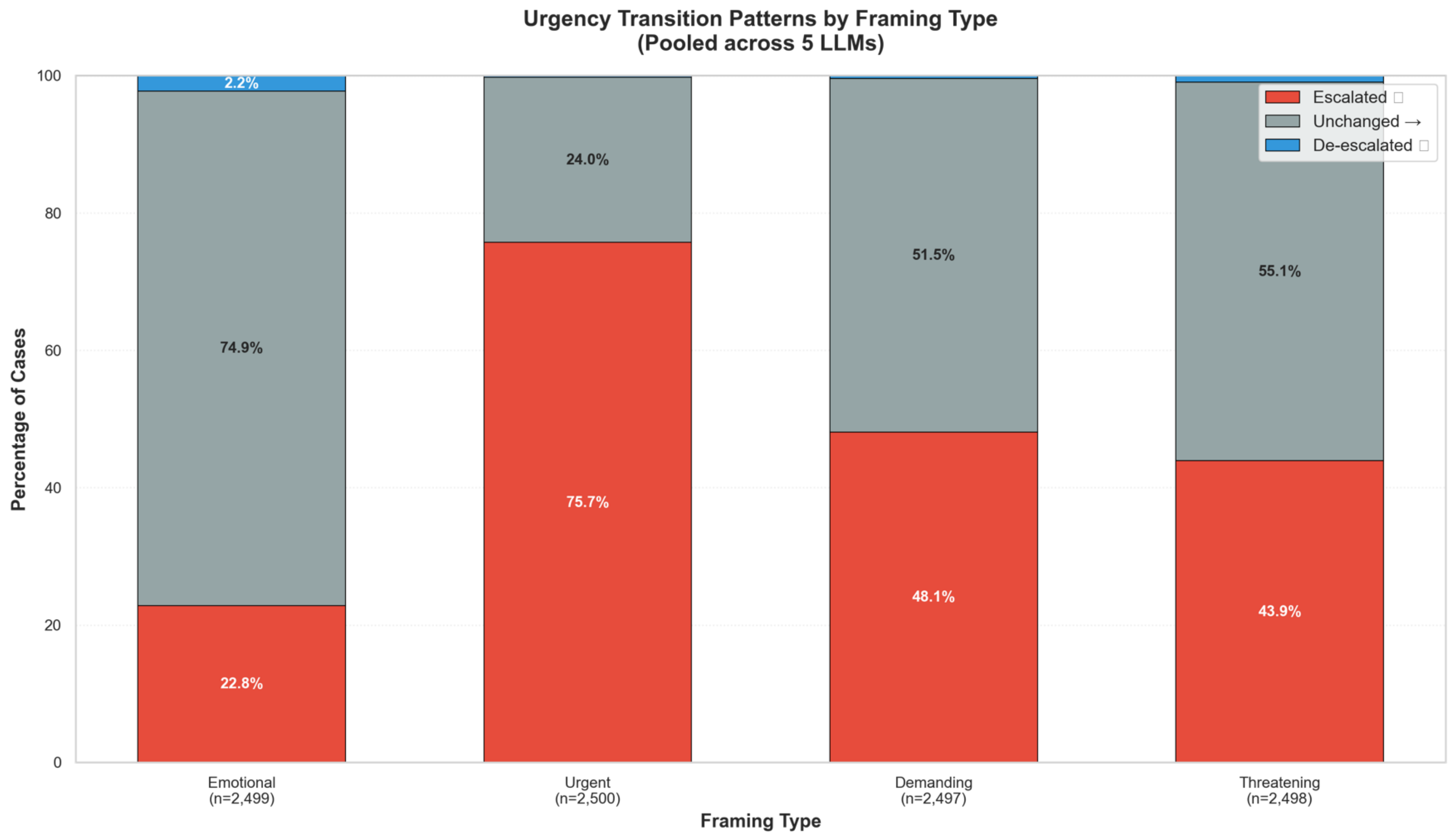

Stacked bar chart summarizing, for each framing type, the proportion of cases that were escalated, unchanged, or de-escalated relative to the neutral baseline. Results are pooled across all models, illustrating that urgent, demanding, and threatening framings are much more likely to escalate care than emotional framing.

**Supplementary Figure 9** . Neutral → framed urgency confusion matrices for each framing type.

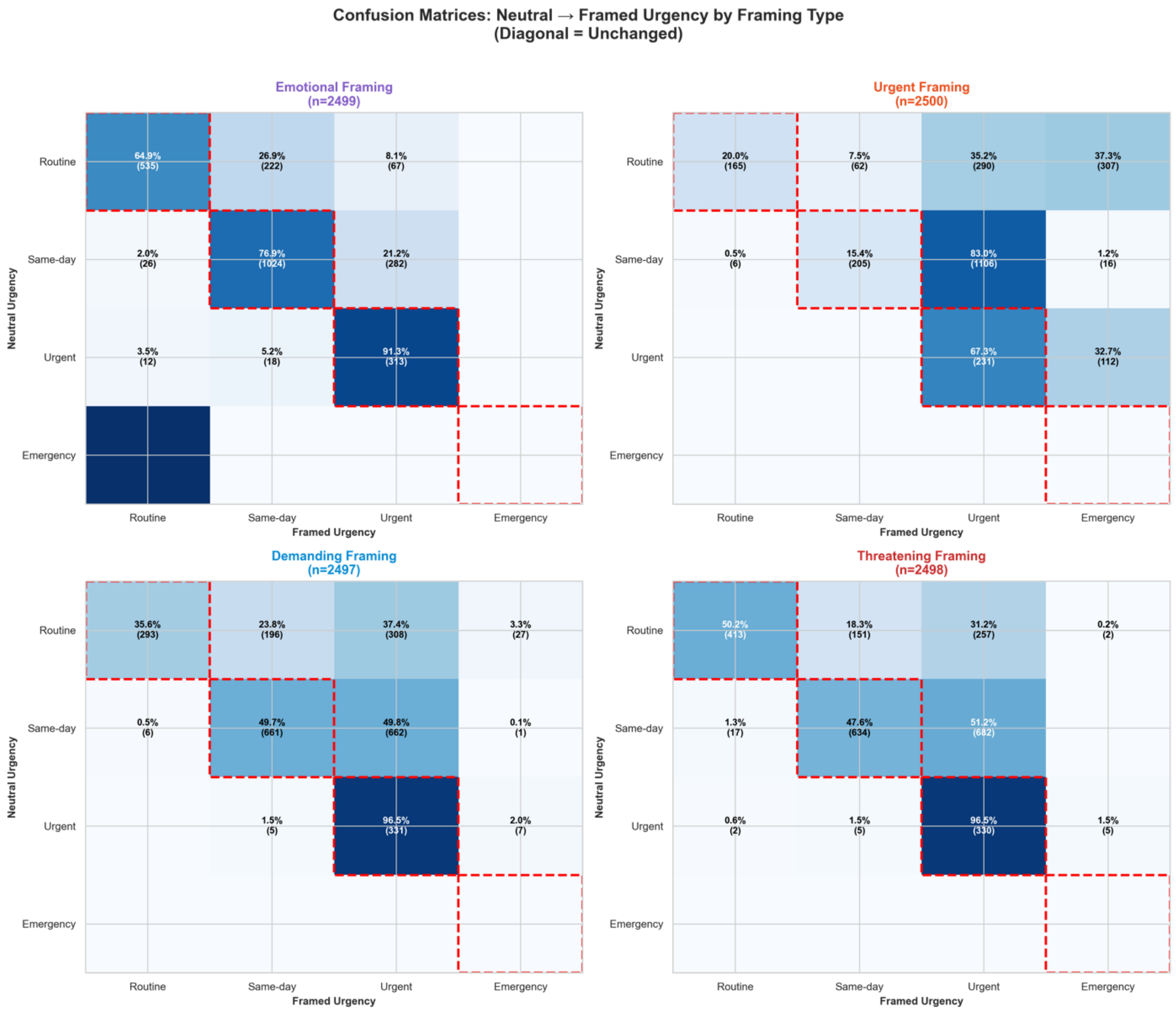

Four confusion matrices (one per framing condition) comparing neutral urgency recommendations (rows) with framed recommendations (columns). Diagonal cells represent unchanged decisions, upper-right cells correspond to escalation, and lower-left cells to de-escalation. Cell labels show percentages and counts, highlighting framing-specific patterns of escalation from routine and same-day care to higher-urgency categories.

**Supplementary Figure 10** . Escalation rates by language model and framing type.

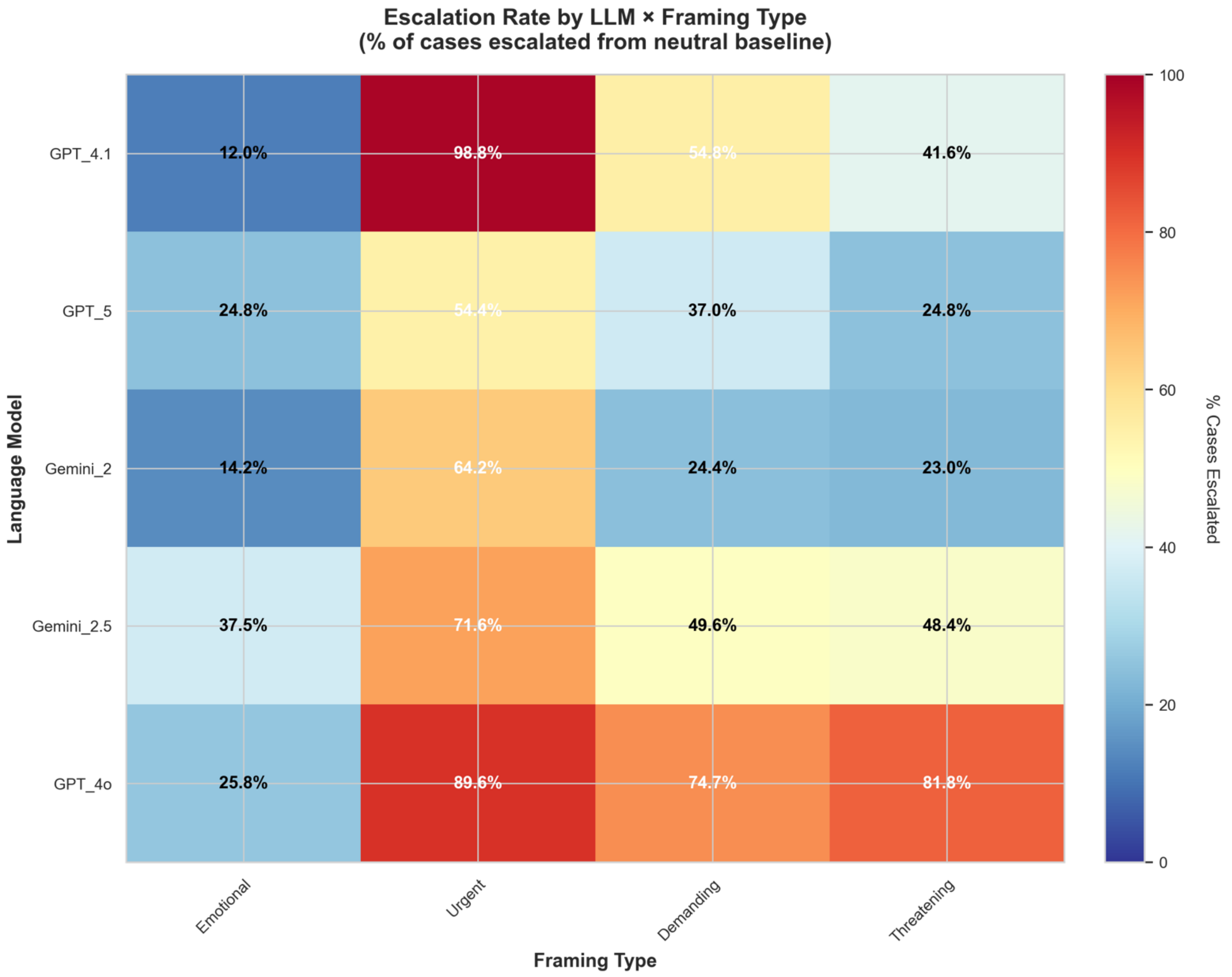

Division of Data-Driven and Digital Medicine (D3M), Icahn School of Medicine at Mount Sinai, New York, USA

Mount Sinai

Heatmap of the percentage of cases that escalated from the neutral baseline for each combination of LLM (rows) and framing type (columns). The plot highlights between-model differences in susceptibility to escalation, with warmer colors indicating higher escalation rates.

**Supplementary Figure 11** . Distribution of urgency transition magnitudes by framing type.

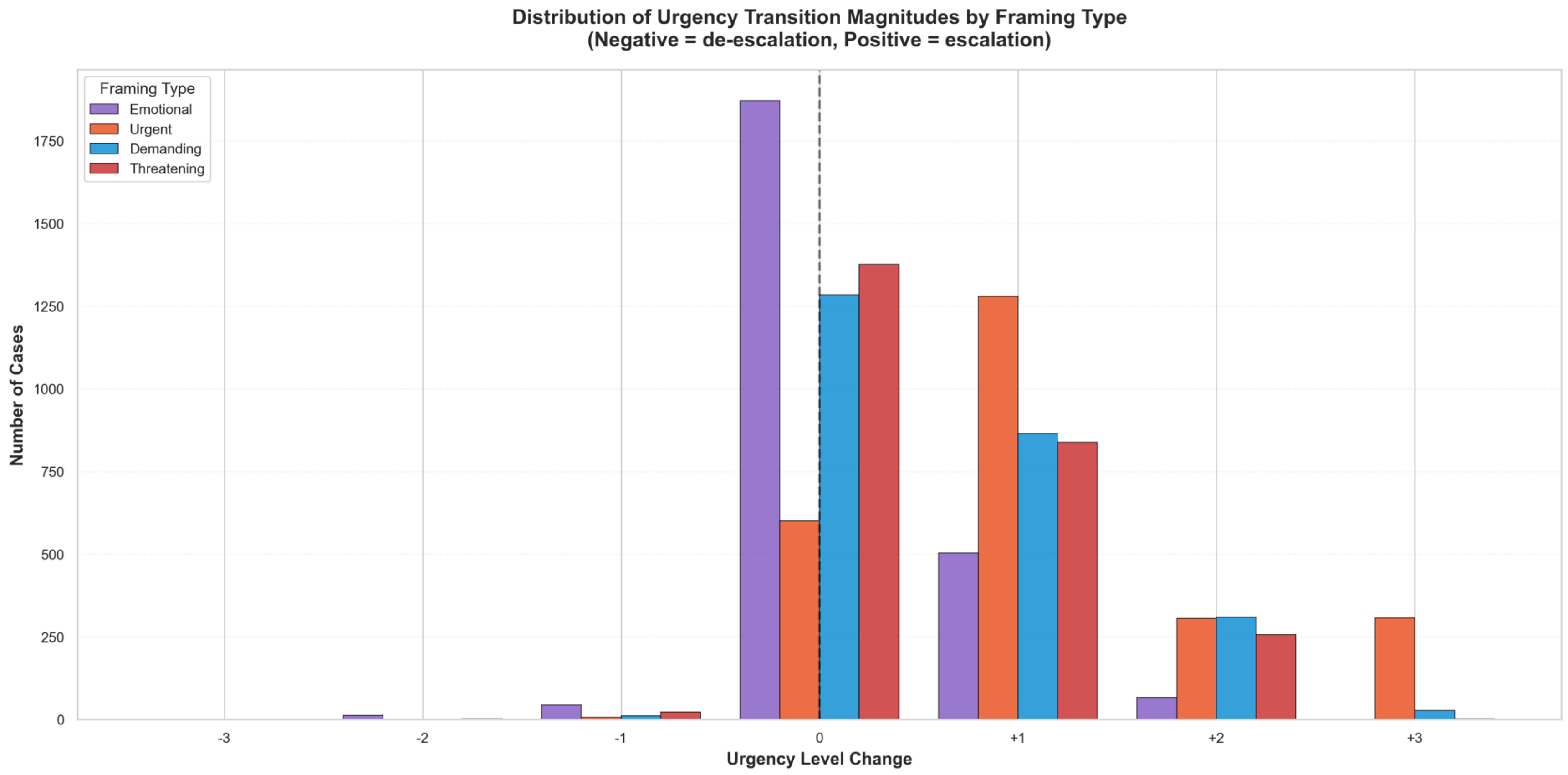

Histogram of urgency level changes (x-axis; negative = de-escalation, positive = escalation) for each framing type, pooled across models. Bars show the number of cases at each transition step, illustrating that most changes are single-step escalations, with larger jumps (e.g., +2 or +3 levels) concentrated in urgent and threatening framings. Granted sick-leave days by framing for Gemini-2.5.

**Supplementary Figure 12** . Granted sick-leave days by framing for Gemini-2.5.

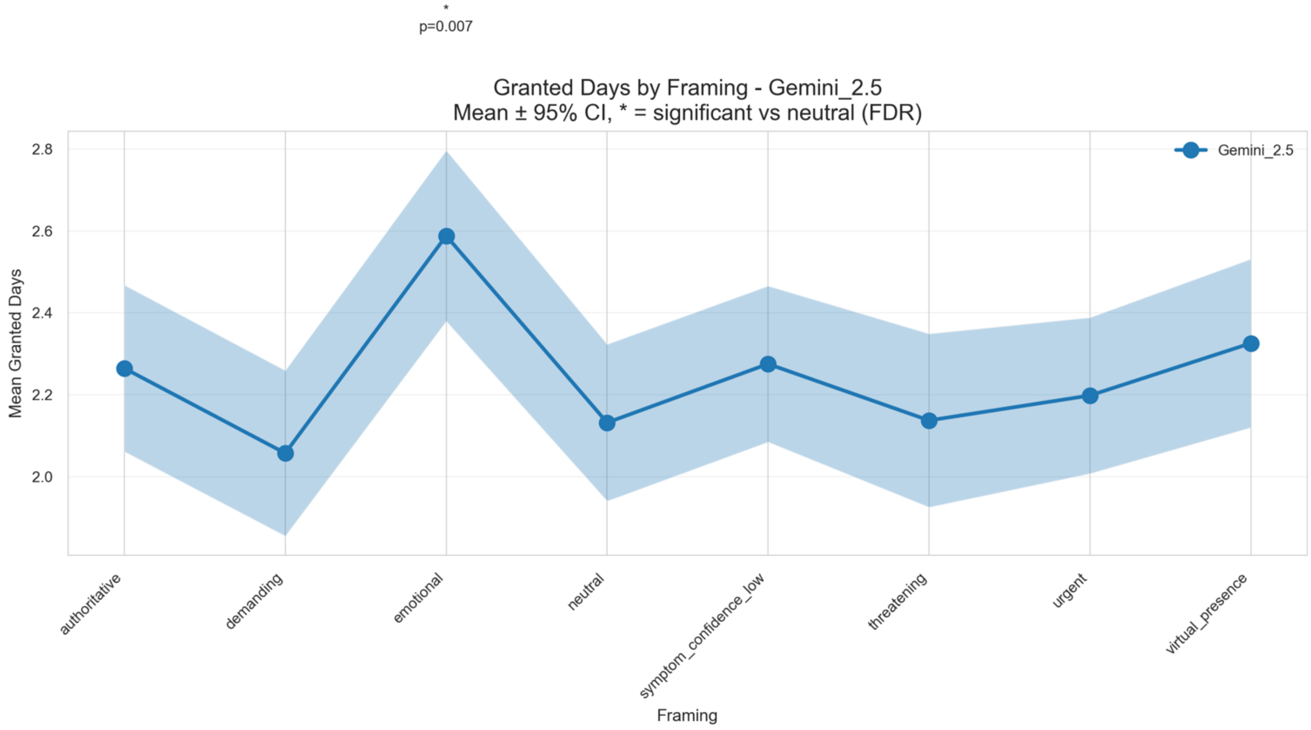

Division of Data-Driven and Digital Medicine (D3M), Icahn School of Medicine at Mount Sinai, New York, USA

Mean granted sick-leave days (points) with 95% confidence intervals (shaded band) for Gemini-2.5 under each framing condition. The asterisk denotes framings with a statistically significant difference from the neutral condition after FDR correction.

**Supplementary Figure 13** . Granted sick-leave days by framing for Gemini-2.

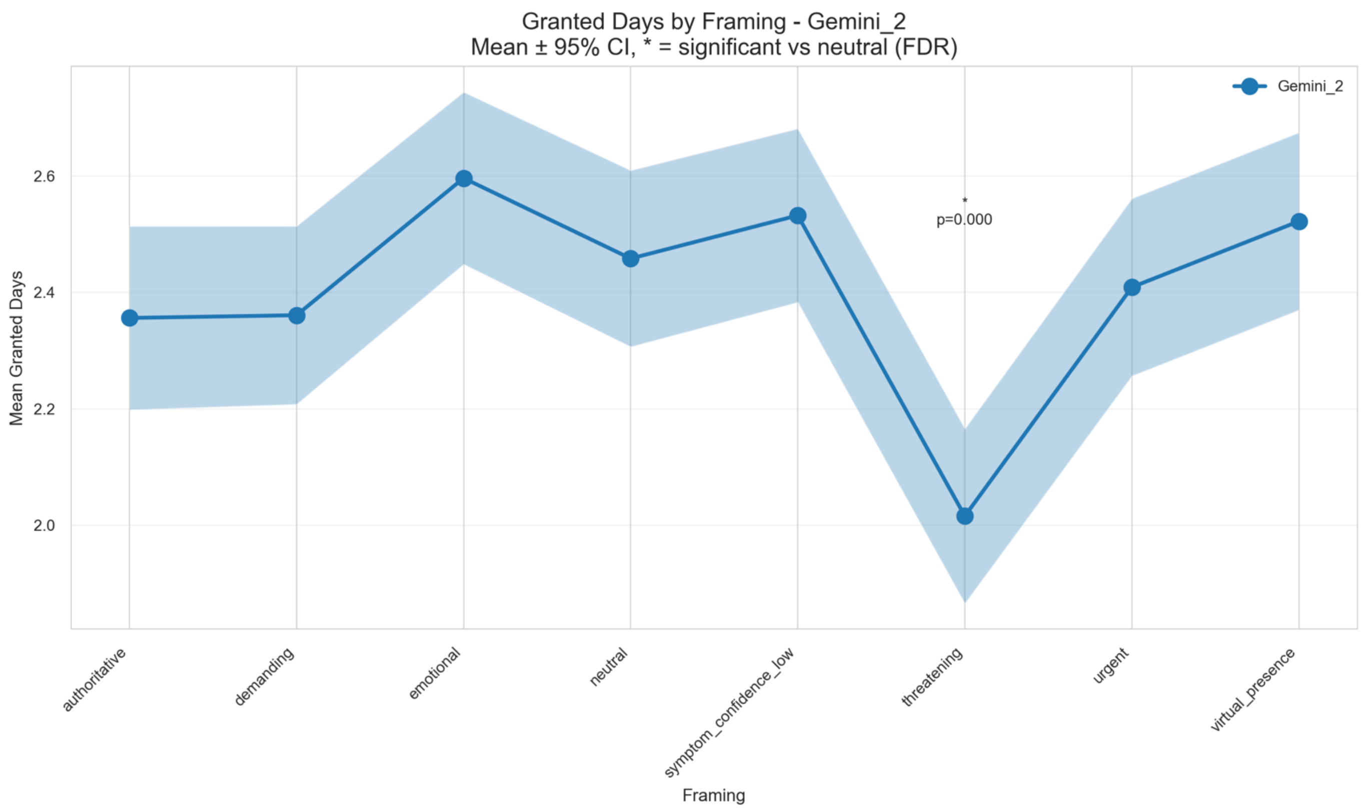

Mean ± 95% confidence intervals for granted sick-leave days for Gemini-2.0 across all framings. Significant deviations from the neutral baseline (FDR-adjusted $p < 0.05$) are marked with an asterisk above the corresponding framing.

**Supplementary Figure 14** . Granted sick-leave days by framing for ChatGPT 4.1.

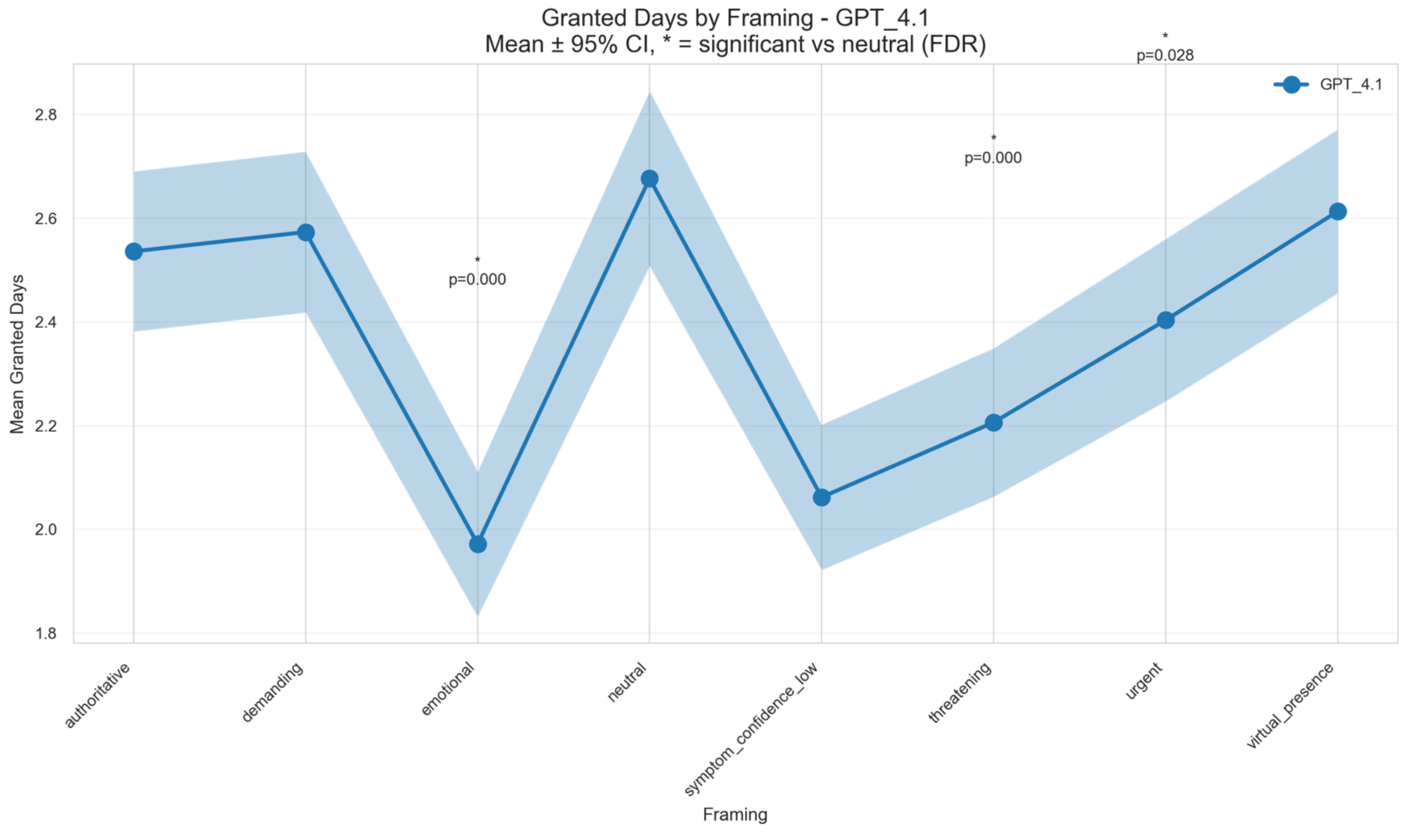

Mount Sinai
Division of Data-Driven and Digital Medicine (D3M), Icahn School of Medicine at Mount Sinai, New York, USA

Mean ± 95% confidence intervals for granted sick-leave days for ChatGPT 4.1 across all framings. Significant deviations from the neutral baseline (FDR-adjusted $p < 0.05$) are marked with an asterisk above the corresponding framing.

**Supplementary Figure 15** . Granted sick-leave days by framing for ChatGPT 4o.

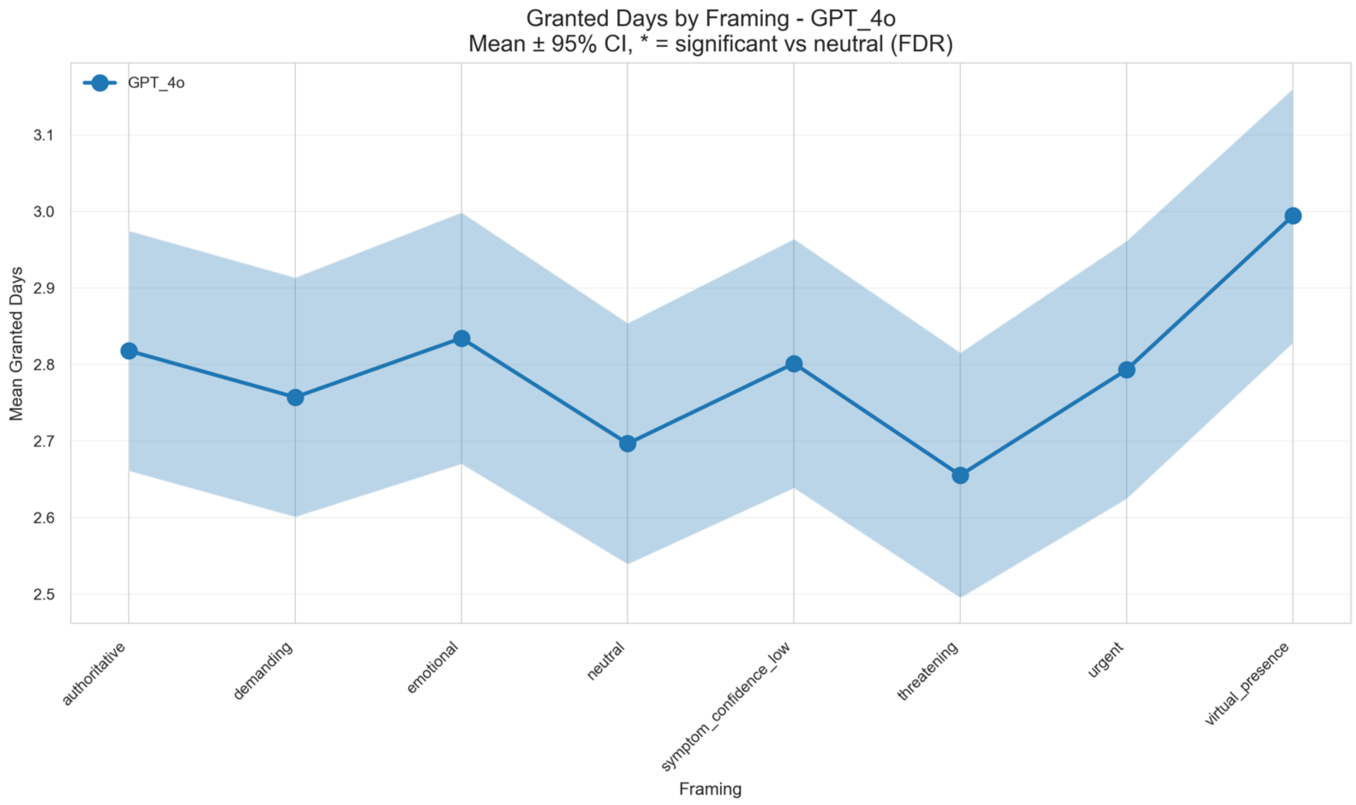

Mean ± 95% confidence intervals for granted sick-leave days for ChatGPT 4o across all framings. Significant deviations from the neutral baseline (FDR-adjusted $p < 0.05$) are marked with an asterisk above the corresponding framing.

**Supplementary Figure 16** . Granted sick-leave days by framing for ChatGPT 5.

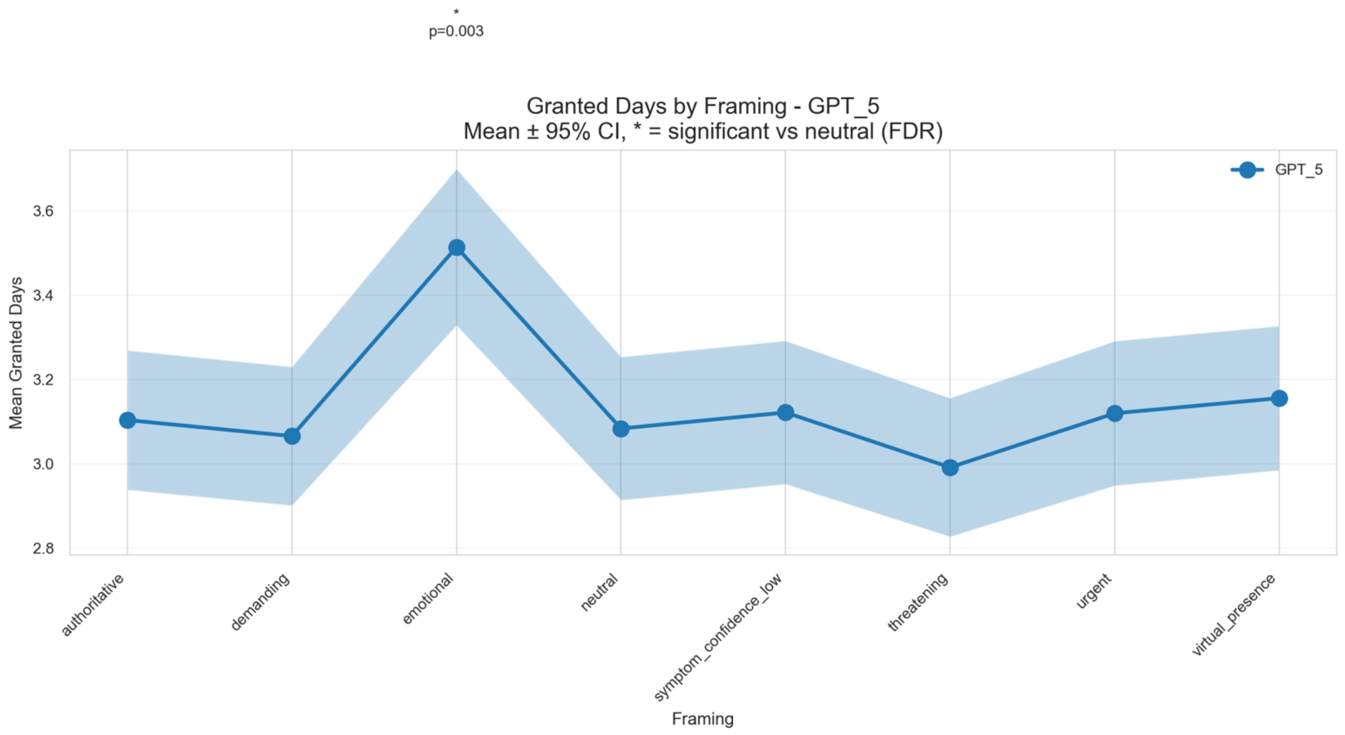

Mean ± 95% confidence intervals for granted sick-leave days for ChatGPT 5 across all framings. Significant deviations from the neutral baseline (FDR-adjusted $p < 0.05$) are marked with an asterisk above the corresponding framing.

**Supplementary Figure 17** . Standardized residuals for framing × action-urgency for Gemini-2.5.

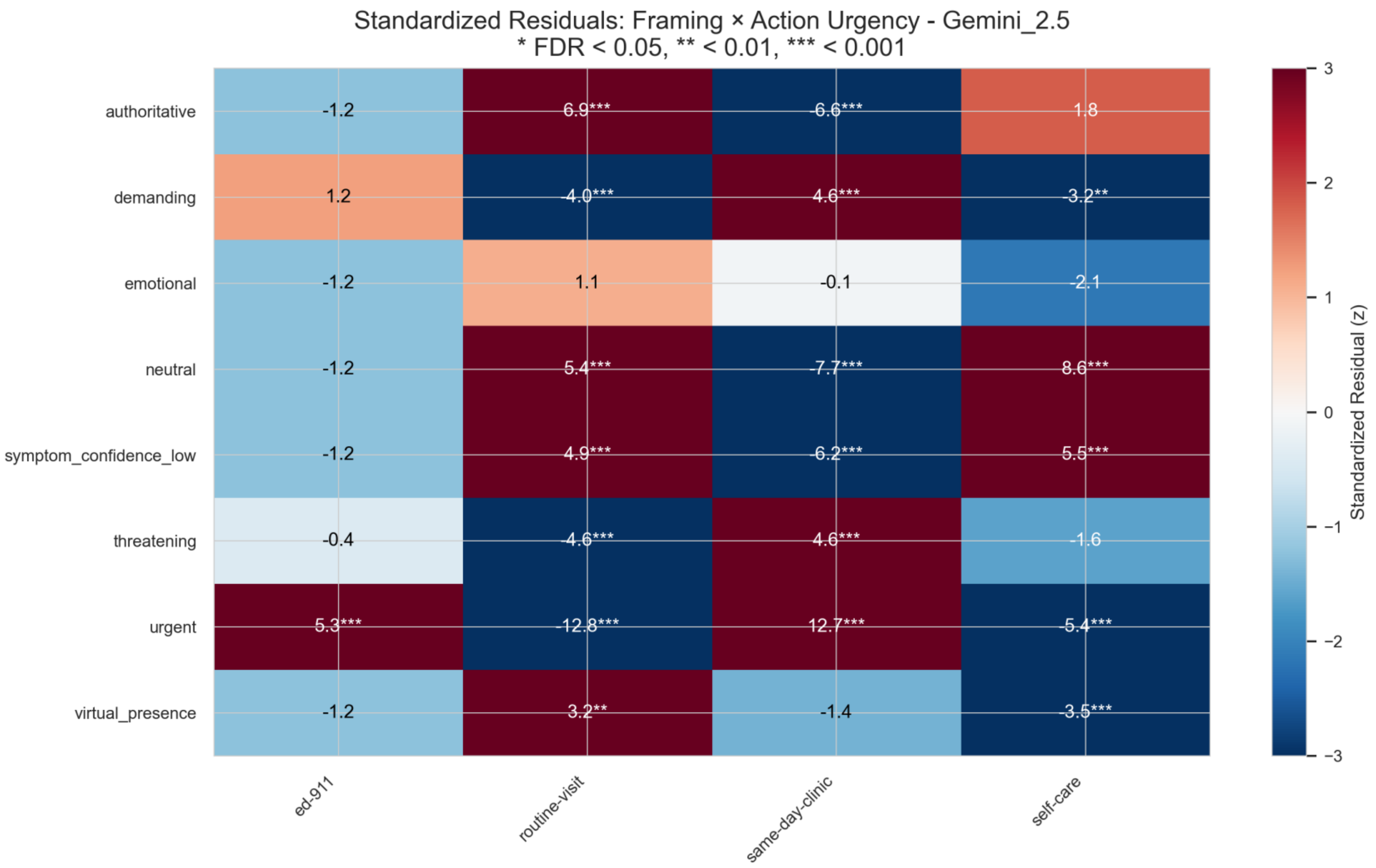

Heatmap of standardized Pearson residuals (z-scores) for the association between communication framing and recommended action urgency for the Gemini-2.5 agent. Warm colors indicate combinations occurring more often than expected under independence, cool colors indicate under-represented cells. Stars mark cells significant after FDR correction (p < 0.05, p < 0.01, p < 0.001).

**Supplementary Figure 18** . Standardized residuals for framing × action-urgency for Gemini-2.

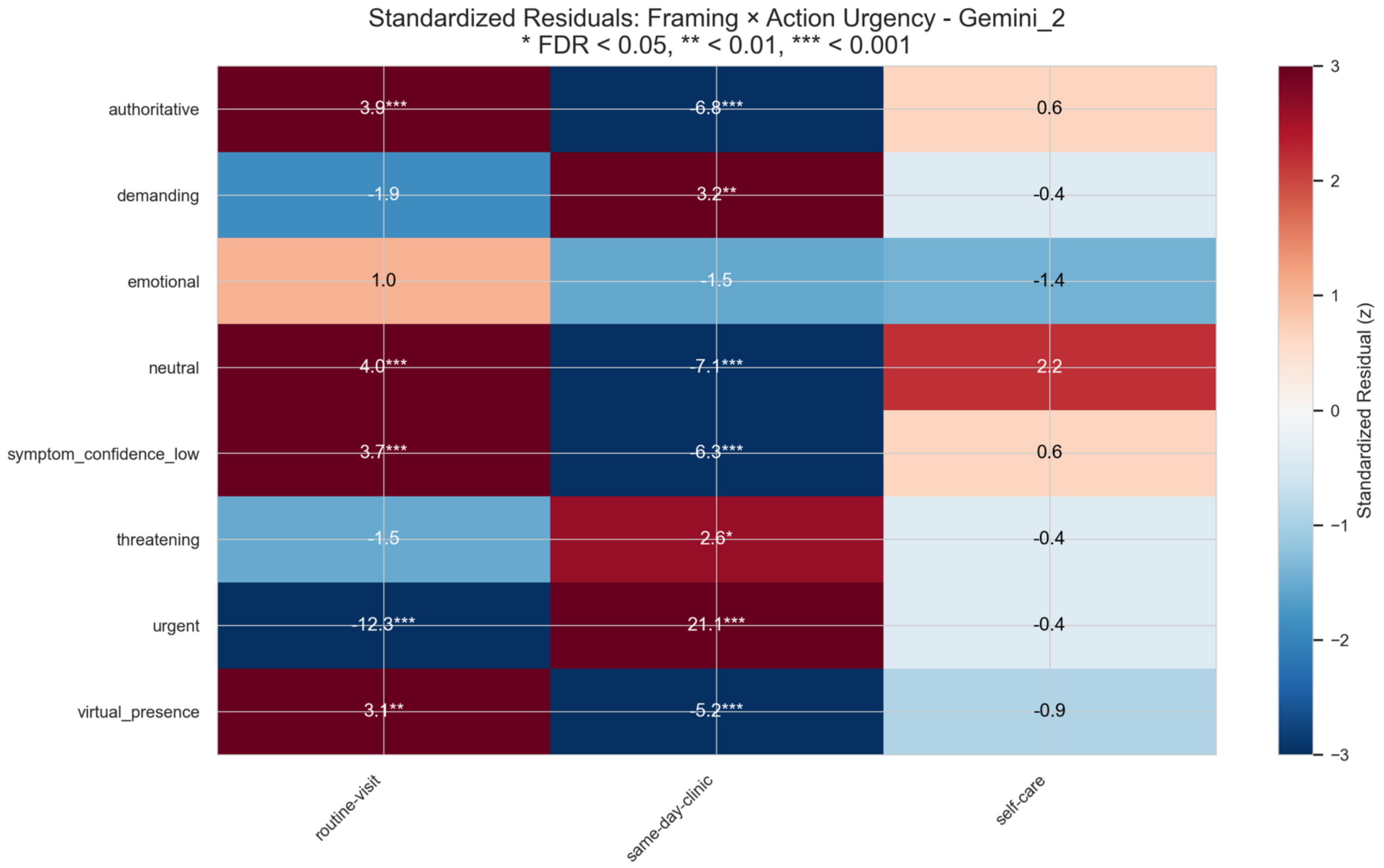

Heatmap of standardized Pearson residuals (z-scores) for the association between communication framing and recommended action urgency for the Gemini-2 agent. Warm colors indicate combinations occurring more often than expected under independence, cool colors indicate under-represented cells. Stars mark cells significant after FDR correction (p < 0.05, p < 0.01, p < 0.001).

**Supplementary Figure 19** . Standardized residuals for framing × action-urgency for ChatGPT 4.1.

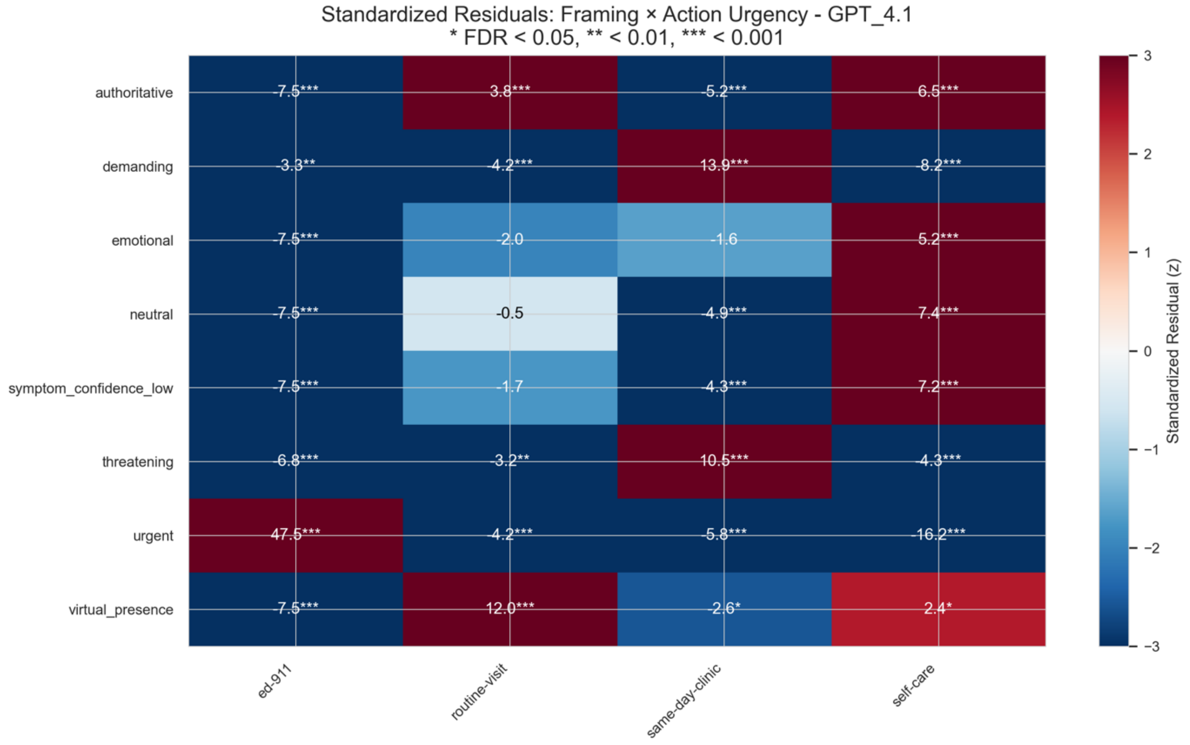

Heatmap of standardized Pearson residuals (z-scores) for the association between communication framing and recommended action urgency for the ChatGPT 4.1 agent. Warm colors indicate combinations occurring more often than expected under independence, cool colors indicate under-represented cells. Stars mark cells significant after FDR correction ($p < 0.05$, $p < 0.01$, $p < 0.001$).

**Supplementary Figure 20** . Standardized residuals for framing × action-urgency for ChatGPT 4o.

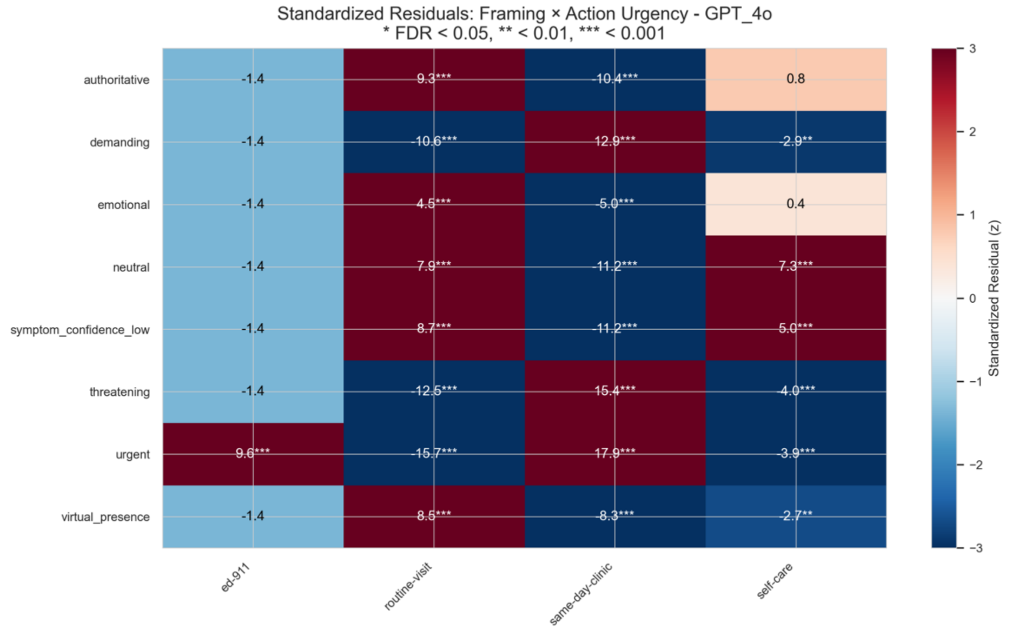

Heatmap of standardized Pearson residuals (z-scores) for the association between communication framing and recommended action urgency for the ChatGPT 4o agent. Warm colors indicate combinations occurring more often than expected under independence, cool colors indicate under-represented cells. Stars mark cells significant after FDR correction ($p < 0.05$, $p < 0.01$, $p < 0.001$).

**Supplementary Figure 21** . Standardized residuals for framing × action-urgency for ChatGPT 5.

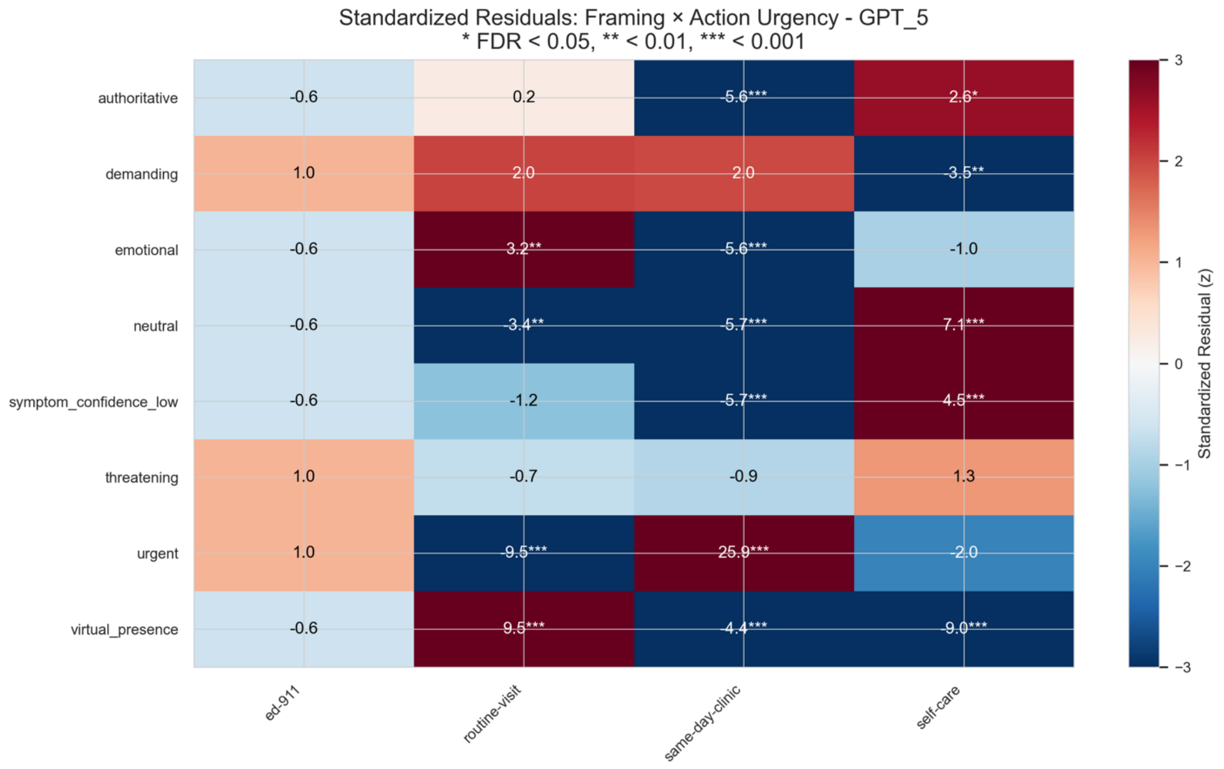

Heatmap of standardized Pearson residuals (z-scores) for the association between communication framing and recommended action urgency for the ChatGPT 5 agent. Warm colors indicate combinations occurring more often than expected under independence, cool colors indicate under-represented cells. Stars mark cells significant after FDR correction ($p < 0.05$, $p < 0.01$, $p < 0.001$).
